# Phenome-Wide Association Study of Actigraphy in the UK Biobank

**DOI:** 10.1101/2021.12.09.21267558

**Authors:** Thomas G. Brooks, Nicholas F. Lahens, Gregory R. Grant, Yvette I. Sheline, Garret A. FitzGerald, Carsten Skarke

## Abstract

Wrist-worn accelerometer actigraphy devices present the opportunity for large-scale data collection from people during their daily lives. Using data from approximately 100,000 participants in the UK Biobank, actigraphy-derived measures of physical activity, sleep, and diurnal rhythms were associated in exploration and validation cohorts with a full phenome-wide set of diagnoses, biomarkers and metadata. Rhythmicity was captured by two independent models based on accelerometer and skin temperature harnessing behavioral (diurnal) and molecular (circadian) components. We found that robust rhythms significantly with biomarkers, survival, and phenotypes including diabetes, hypertension, mood disorders, and chronic airway obstruction; these associations were comparable to those with physical activity and sleep. Surprisingly, associations were mostly consistent between the sexes, while modulation by age was significant. More importantly, rhythms were found to be powerful predictors of future diseases: a two standard deviation difference in wrist temperature rhythms corresponded to increases in rate of diagnosis of 61% in diabetes, 38% in chronic airway obstruction, 27% in anxiety disorders, and 22% in hypertension. Our PheWAS of actigraphy data in the UK Biobank establishes that rhythmicity is fundamental to modeling disease trajectories, as are physical activity and sleep. Integration of long-term remote biosensing into patient care could thus afford an individualized approach to risk management.

## Introduction

Genome-wide association studies (GWAS) [1] offer a systematic approach to identify genetic variants for their association with specific phenotypes of interest. Phenome-wide association studies (PheWAS) [2] go further and screen the multitude of phenotypes available from electronic health records (EHR) for their association with specific genetic variants of interest. These power discoveries in large-scale biobanks annotated with rich EHRs and have elucidated disease mechanisms, so that they can be exploited for treatment and prevention. A recent example illustrates how these approaches can be leveraged to discern new genetic markers of lipid levels in the Million Veteran Program [3]. The success of PheWAS on genetic variation and the strong association of accelerometry-based behavioral phenotyping and mortality [4, 5] and its potential use in disease mapping [6, 7] highlight the need to develop phenome-wide approaches for associating behavior with health outcomes.

Here, we develop this phenome-wide approach for remotely sensed behavioral health measures, quantified per wrist actigraphy in the UK Biobank (UKB). We associated these metrics with phenome-wide disease phenotypes and quantitative traits, and accounted for medication exposure extracted from the UKB. To distinguish this from traditional PheWAS approaches that associate phenotypes with genetic variants, we coined the term *<u>Phe</u>nome-<u>W</u>ide <u>A</u>ssociation <u>S</u>tudies <u>Sens</u>ed <u>R</u>emotely* or *PheWAS-SensR*.

Our PheWAS-SensR analysis underscores the relevance of physical activity and sleep for human health and extends this to consolidated diurnal variability as an equivalent behavioral health dimension; all three categories of measures were significantly associated with disease phenotypes. Associations between these behavioral health measures and disease phenotypes or quantitative traits were mostly consistent between the sexes. Age, in contrast, significantly modulated these relationships. We established robustness to determine a phenotype’s diurnal variability by both accelerometer-(capturing diurnal rhythmicity) and temperature-based (capturing circadian rhythmicity) modelling.

Importantly, we found that wrist temperature rhythms are powerful predictors of future diseases. Phenotypes including diabetes, hypertension, chronic airway obstruction, anxiety and pneumonia exhibited large increases in diagnosis rates in individuals who showed reduced circadian rhythms. Concretely, our PheWAS-SensR quantified physical activity, sleep, and rhythms from remote sensors to model disease trajectories.

## Methods and Results

### Architecture of the PheWAS-SensR Cohort

The UKB project collected 7 days of actigraphy data from 103,688 participants roughly from June 2013 to December 2015 (Figure S 1 top left) [8]. Participants were excluded from further analyses based on failed calibration (n=2,826), insufficient data (n=4,258) or crossing over daylight savings time (n=4,279), following prior experiments[8-10]. A total of 92,325 participants passed to enter the PheWAS-SensR. We divided the participants into two cohorts by random sampling: 25,000 participants formed an exploratory cohort, and the remaining 67,325 participants formed an independent validation cohort (Figure S 1 bottom left). All results are reported from the validation cohort and are consistent in both cohorts, see Supplemental File 1.

From the week-long actigraphy readings, 214 objective behavioral health measures were derived for each participant as detailed in the Supplemental Methods. In brief, the data were calibrated [8] and classified by a machine learning pipeline [11], giving each 30 second epoch one of six actigraphy type labels (such as “sleep” or “walking”). Actigraphy health variables were derived via cosinor fits, circadian parameters, Gaussian sliding windows identifying daily peaks, and summary statistics.

Behavioral health measures were assessed for stability within an individual across different measurements, using the repeated data collections performed on a subset of individuals (n=2,339 with five week-long measurements). After discarding health metrics displaying more variability between repeated measurements on the same individual than between individuals, a total of 116 actigraphy-derived behavioral health measures remained.

Measures were furthermore generated from temperature level sensors included on the actigraphy device. Skin temperature reflects circadian rhythmicity [12], and thus offered an orthogonal assessment to diurnal variability derived from physical activity.

Eight self-reported health variables physical activity, sleep and chronotype were included. This created a final catalog of 124 behavioral health measures (Table S 3) in the PheWAS-SensR cohort.

We annotated by expert opinion (T.B., C.S.) each behavioral health measure as being reflective of physical activity (60 metrics), sleep (24 metrics) or diurnal rhythms (40 metrics). A typical example of a physical activity metric is acceleration_overall (mean acceleration), typical examples of sleep metrics are main_sleep_duration_mean (sleep duration of longest sleep period) and main_sleep_ratio_mean (mean fraction of longest sleep period spent asleep, commonly referred to as sleep efficiency), and a typical example of a diurnal rhythm metric is acceleration_RA (normalized difference in day and night activity levels) (Table S 3).

### Phenotypes

We derived the lifetime history of diagnoses for the PheWAS-SensR participants from medical record ICD-10 and ICD-9 codes as well as from self-reported conditions during the initial assessment dated 2007-2010 (Figure S 1 top left). ICD-10 and ICD-9 codes were grouped into phenotypes by mapping to PheCODES, an algorithm developed for the Vanderbilt University Medical Center (VUMC) and UKB datasets [13, 14] using Phecode Map v1.2b1. Self-reported diagnoses were mapped by expert opinion (C.S.) to the PheCODES (Table S 3).

Previous works have found a “healthy volunteer” effect in the UKB where participants have greater health than the overall population [15]. This bias increased among participants who enrolled in actigraphy, who had overall fewer diagnoses at study end (mean±SD 9.3±11.5 per person) than those who did not enroll in actigraphy (11.7±14 per person) (Figure S 1 top right).. Compared to the UKB overall, individuals with actigraphy tended to be slightly more white, more female, younger and had a lower BMI (Table S 1).

As expected, medical record diagnoses accelerated over time, culminating in a total of 478,697 distinct diagnoses to date (Figure S 1 top). Furthermore, a total of 1,516 deaths were recorded among PheWAS-SensR participants (2.25%, per data download on November 20, 2020, Figure S 1 bottom).

To complement diagnosis PheCODEs, 83 quantitative phenotypes were derived from measures, taken at initial assessment, of blood assays, urine assays, and physical measurements.

### Phenome-Wide Association Study Sensed Remotely through wearable devices (PheWAS-SensR)

For diagnoses with at least 50 cases, a phenome-wide association of actigraphy health variables and diagnoses was performed (Figure 1, Table S 4, Figure S 2 top). To achieve this, a linear model was computed for each actigraphy health measure and PheCODE pairing. The model included covariates for sex, ethnicity (white/other), self-reported overall health, high household income (≥ 52,000 pounds sterling/year), age at the time of actigraphy, BMI, college education and if they’ve ever smoked, all measured at the initial assessment. This resulted in a total of 55,676 statistical tests performed with 12,081 of them achieving FDR significance of *q*<0.05 and 3,357 achieving Bonferroni-corrected significance of *p*<0.05.

**Figure 1.**
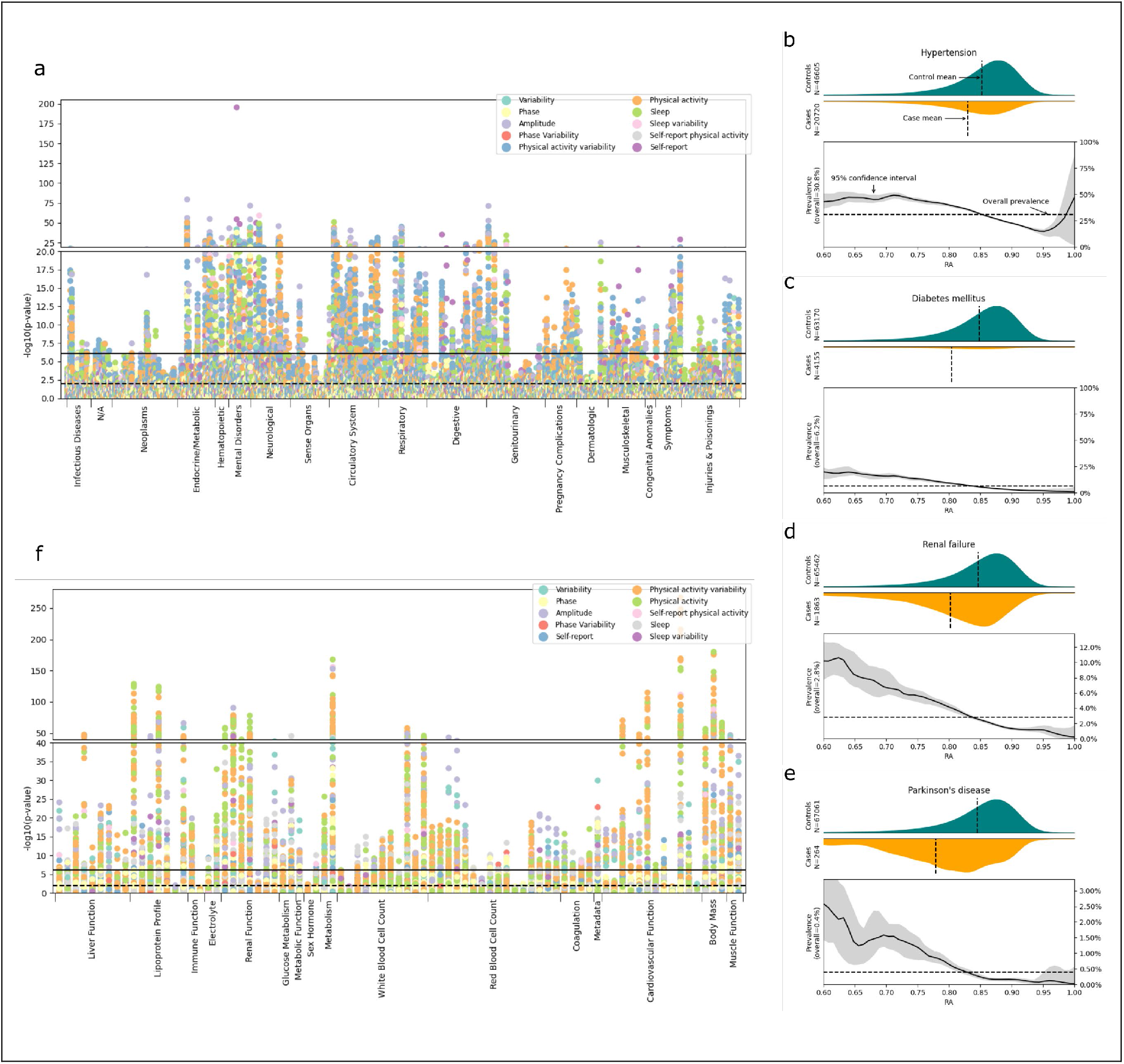
Associations between Disease Phenotypes and Behavioral Measures. (a) Manhattan plot of -log_10_ *p*-values of associations between behavioral measures and PheCODE. The Benjamini-Hochberg FDR cutoff of *q* < 0.05 is shown in the horizontal dashed line and a Bonferoni cutoff for *p* < 0.05 in horizontal solid line. To enhance visualization, the vertical axis is broken. (b-e) Example associations of the most common phenotypes, (b) hypertension, and the RA score as well as other representative disease phenotypes, (c) diabetes mellitus, (d) renal failure, and (e) Parkinson’s disease. The top graph in each panel shows the distributions of RA scores in cases (orange) and controls (torquise). The bottom graphs display the prevalence of the diagnosis by RA score. As compared to the overall population, those with lower acceleration_RA than the mean (0.845) had an increased prevalence of the disease, while those with higher RA scores had decreased prevalence. Note that due to low case counts in renal failure and Parkinson’s, axes have been rescaled. (f) Manhattan plot of -log_10_ *p*-values of associations between behavioral measures and quantitative phenotypes.

Disease phenotypes across all major organ systems associated significantly with physical activity, sleep and diurnal variability (Figure 1 a).

Overall, in healthy people we observed greater daytime activity paired with low nighttime activity. This difference is summarized using the relative amplitude (RA) score, in the measure “acceleration_RA” [16]. This is computed as 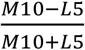 where *M10* is the average activity level during the 10 most active hours of the individual’s day, and *L5* is the average activity during the 5 least active hours. Values of acceleration_RA near 1 reflect strong daily activity rhythms while lower values indicate inconsistent or low-amplitude rhythms. Few people reached an acceleration RA lower than 0.65 to 0.7, suggesting adherence to the common day-night activity-inactivity rhythm. Only a few participants reached an acceleration RA higher than 0.95, suggesting that the summary stats past 0.95 should not be considered reliable (Figure 1 b).

We found that average acceleration_RA was lower in many disease conditions including diabetes mellitus, renal failure, Parkinson’s disease, chronic airway obstruction, hypertension, and pneumonia. In the case of hypertension, participants with an acceleration RA of ≈0.75 had a 50% prevalence of comorbid elevated blood pressure, while participants displaying an acceleration RA close to 0.9 barely registered with high blood pressure (≈15% prevalence). This was reflected in the overall lower mean of 0.83 acceleration RA in cases (*n*=20,720) compared to 0.85 acceleration RA in controls (*n*=46,605) (*q*=9.2×10^−43^). We observed similar associations for other diseases, shown in Figure 1 c-e for diabetes mellitus, renal failure, and Parkinson’s disease. Patients with Parkinson’s disease showed, as expected [17], a low mean acceleration RA of 0.78.

To examine physical activity-related PheCODEs, we selected the acceleration_overall measure, which is the mean activity acceleration level. The most significant associations with it were with hypertension (*q* = 9.5×10^−42^, Cohen’s d = -0.11) and tobacco use disorder (*q* = 9.3×10^−40^, d = -0.27). Next are Parkinson’s disease (*q* = 5.3×10^−33^, d = -0.72), diabetes mellitus (*q* = 1.7×10^−27^, d = -0.18), and renal failure (*q* = 4.8×10^−27^, d = -0.25).

For sleep quality, we examined the main_sleep_ratio_mean (i.e., sleep efficiency [18]). Its top associations were also with hypertension (*q* = 4.6×10^−48^, d=-0.13) and Parkinson’s disease (*q* = 2.7×10^−46^, d=-0.88). In addition, it associated with diabetes mellitus (*q* = 4.7×10^−39^, d=-0.22), renal failure (*q* = 7.6×10^−36^, d=-0.30), and with mood disorder (q=2.6×10^−32^, d = -0.21).

In summary, these findings illustrate how well actigraphy-derived metrics of physical activity, sleep, and diurnal rhythmicity track with disease phenotypes.

We further examined behavioral health associations with the quantitative phenotypes (Figure 1 f, Figure S 2 bottom, and Supplemental Results) where we found congruent results with the diagnosis associations.

### Wrist Temperatures Mark Circadian Rhythms and Disease State

Wrist temperature rhythms have been used to mark circadian rhythms [12] and are comparable to melatonin and core body temperature in estimating circadian phase [19]. Wrist temperature from the UKB study may reflect a combination of environmental ambient temperature and skin temperature and was collected originally to calibrate the accelerometer [20]. The traces, however, accurately captured the diurnal variability in distal skin temperature as previously reported. One prior report [21] had, in *n*=103 healthy volunteers, the delta between maximum (36.1±0.5°C) and minimum (30.4±1.7°C) rhythmic temperature fluctuations was around 6°C, which corresponds exactly to our observations (Figure 2). Furthermore, we hypothesized that chronotype (i.e., evening or morning preference) shifts the skin temperature rhythm curve [22]. Indeed, morningness associated with a larger amplitude in the skin temperature rhythm than eveningness (*p*=3×10^−58^ with an effect size of 0.23°C) and as expected the mid-morning nadir was phase advanced (Figure 2 a). In contrast, self-reported nappers had reduced temperature rhythms [23], see Figure S 3 (*p*=2×10^−67^ and effect size of 0.3°C change in amplitude comparing never-nappers to usual-nappers).

**Figure 2.**
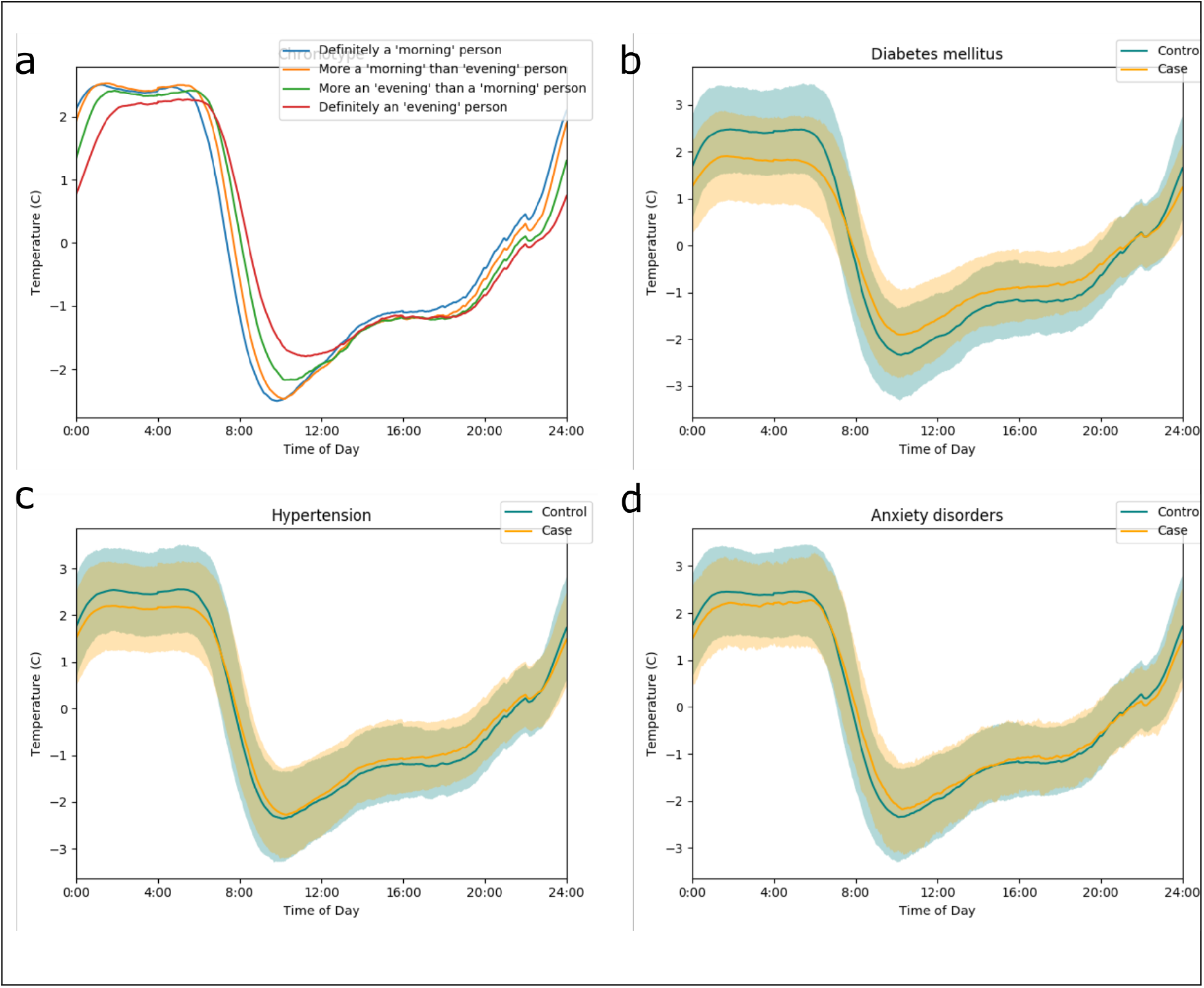
Wrist temperature traces. Wrist temperature traces comparing cases and controls for differences by (a) morning/eveningness, (b) diabetes mellitus, (c) hypertension, and (d) anxiety disorders. 25^th^ to 75^th^ percentiles of the population are displayed in shaded regions (controls in blue, cases in yellow and overlap in grayish green). Temperature is plotted relative to an individual’s overall mean.

Participants with a diagnosis of diabetes mellitus, hypertension, or anxiety disorders showed a reduced amplitude of the wrist temperature-over-time curve (Figure 2 b-d). We did not find an interaction of disease with chronotype: the difference between amplitudes of cases and controls in evening chronotypes was similar to the difference in morning chronotypes (Figure S 3). Dysfunction of adipose tissue in individuals with elevated body weight may cause various comorbidities [24]. An increase in BMI was associated with a proportional loss of amplitude in wrist temperature, without modulation by chronotype (Figure S 3).

Taken together, these observations strongly support using skin temperature variance over the day, temp_RA, as marker of circadian rhythm. We define temp_RA as 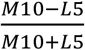 where *M10* is the average temperature during the 10 consecutive hours of highest activity and *L5* is the average temperature during the lowest 5 hours of activity. The *M10* and *L5* reflect the same time periods used in the acceleration_RA, and not necessarily the lowest and highest times for the temperature itself. In the computed associations to quantitative traits and disease phenotypes, significant hits included HDL-C (*q* < 10^−39^), triglycerides (*q* < 10^−38^), hypertension (*q* < 10^−32^), diabetes, mood and anxiety disorders (*q* < 10^−7^) and respiratory conditions (chronic airway obstruction q < 10^−13^, asthma *q* < 10^−9^).

### Deconstructing Disease Phenotypes into the Behavioral Health Components: Physical Activity, Sleep, & Diurnal Rhythmicity

One limitation is that multiple behavioral measures have been derived from a single data source (acceleration values) and therefore these may reflect a combination of behaviors. For example, the acceleration_RA score measures daytime-nighttime differences and was classified as a measure of diurnal rhythmicity. Despite this, it is influenced by physical activity levels (*r*=0.66 Spearman correlation between acceleration_RA and overall acceleration levels) as well as sleep quality. An alternative for this is to use temp_RA, which has lower correlations with activity levels (*r* =-0.25).

To separate the contributions of diurnal rhythmicity independent from other health measures, we first chose behavioral health measures from each of the categories of physical activity, sleep, and circadianness. Specifically, we chose acceleration_overall for physical activity, sleep efficiency (main_sleep_ratio_mean) for sleep, and temp_RA for diurnal rhythms. We performed a logistic regression, modelling each PheCODE phenotype as a function of the three selected behavioral measures, identifying the contributions of activity levels, sleep, and diurnal rhythms (**Figure 3** a, Table S 6). For the quantitative phenotypes, we performed a linear model of the phenotype as a function of the three behavioral measures (**Figure 3** b, Table S 6). From these, we obtained significance of any one variable while controlling for the other two.

**Figure 3.**
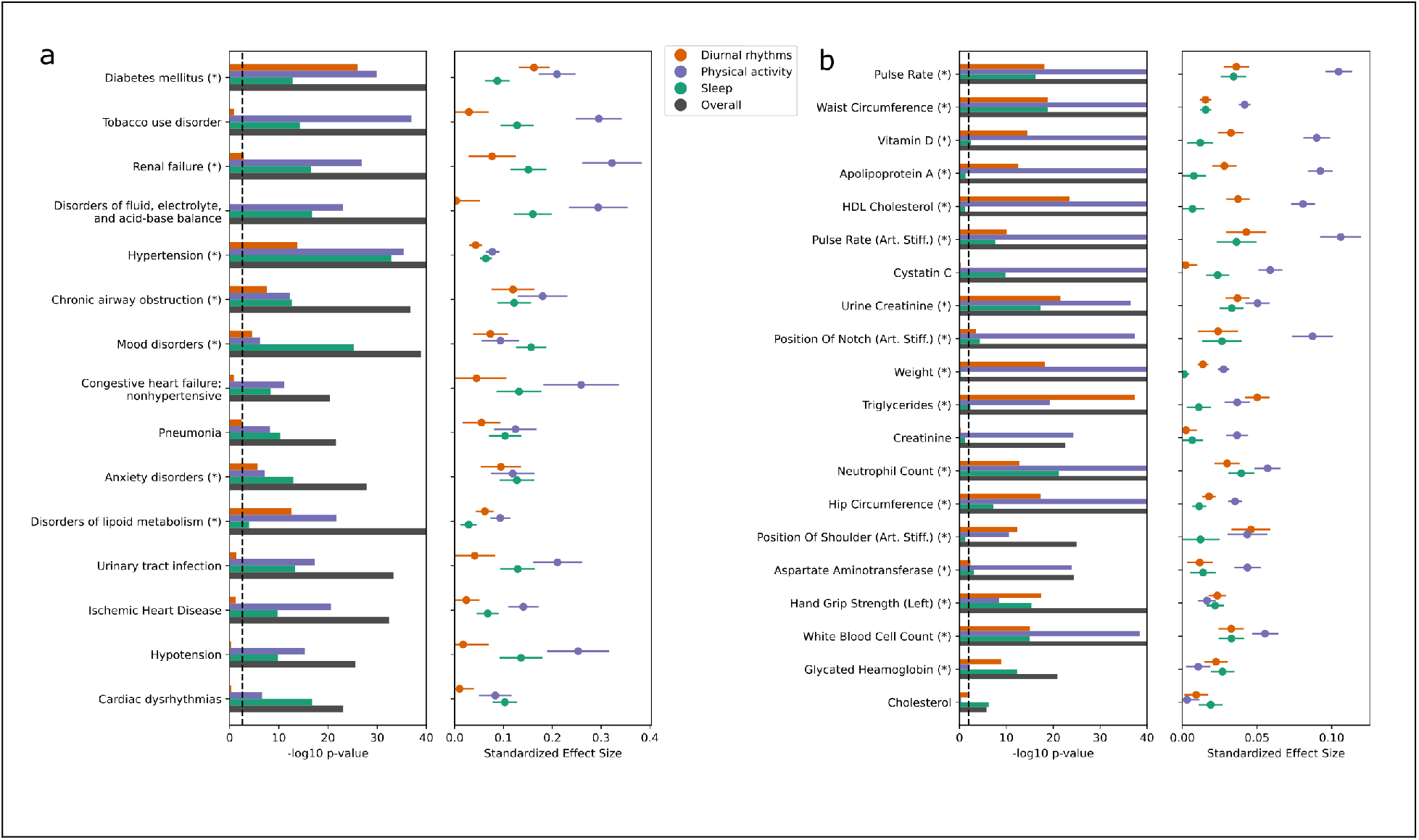
Characterizing Disease Phenotypes and Quantitative Traits by Sleep, Physical Activity, and Diurnal Rhythmicity. Logistic and linear regression were fit for top diagnoses (a) and quantitative phenotypes (b), respectively, as functions of three behavioral health measures: one from each of physical activity (blue), sleep (green), and diurnal rhythmicity (orange) categories. In the left panels, the -log_10_ p-values of the contributions of each of the three variables (capped at 40). The vertical dashed line denotes the FDR q < 0.01 cutoff and phenotypes whose diurnal variability contribution is significant at this level are marked with an asterisk. In right panels, the standardized effect sizes of the model coefficients (absolute population mean marginal effect size per standard deviation of the actigraphy health variable, divided by the phenotype’s overall prevalence) with 95% confidence intervals.

In both cases, significant contributions of diurnal rhythms (as measured by temp_RA) were identified after controlling for both physical activity levels and sleep quality across multiple phenotypes. Prominently associated PheCODES were diabetes mellitus, hypertension, asthma, chronic airway obstruction, lipoid disorders, mood disorders, and anxiety disorders (all with *q*<0.0002); the most significant quantitative traits included triglycerides, HDL-C, and urine creatinine (all with *q*<10^−20^). These indicated a significant contribution of diurnal rhythmicity, quantified by wrist temperature, beyond sleep and physical activity levels towards disease state associations.

### Behavioral Health Measures are Strong Predictors of Survival

Balanced physical activity and stable sleep-wake rhythms are predictors for survival [25, 26]. To address this hypothesis in the UKB we performed a Cox proportional hazards model of all-cause mortality to test for associations between survival and each behavioral measure. The model controlled for sex as well as BMI and smoking status at time of initial assessment in 1,516 deaths. One hundred and eight of the 124 actigraphy variables were significantly associated (FDR *q* < 0.05) with survival in the model (Figure 4, Table S 3).

**Figure 4.**
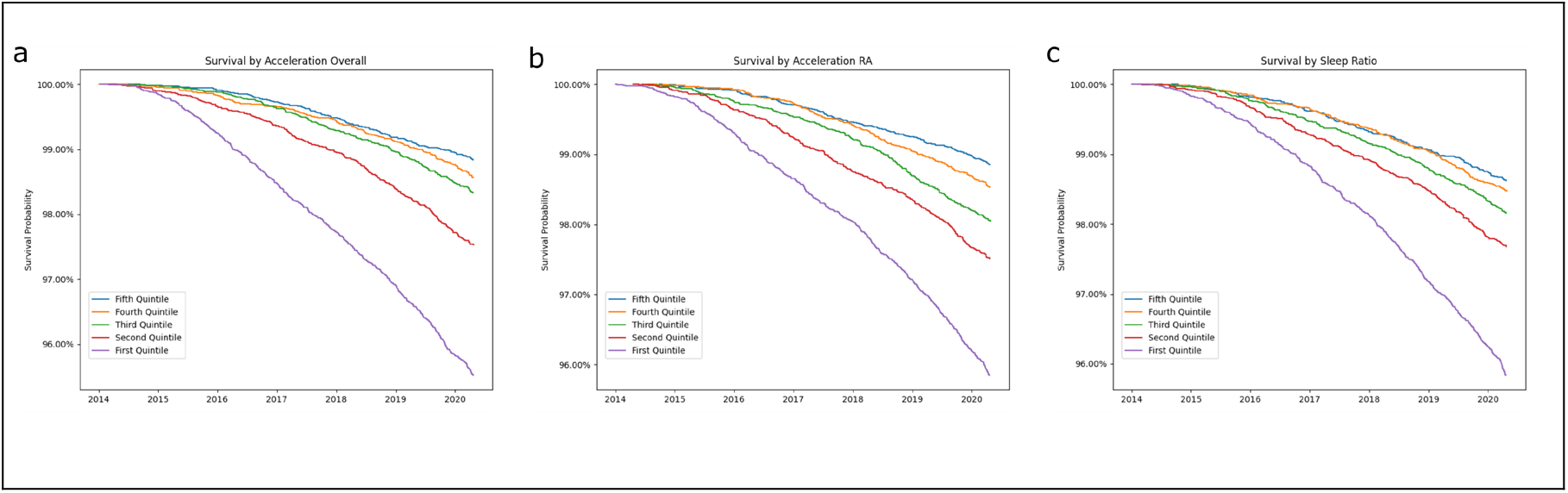
Associations Between Behavioral Health Variables and Survival / Disease Phenotypes. Survival curves by quintiles of behavioral health variables for physical activity (acceleration_overall), diurnal rhythmicity (acceleration_RA) and sleep (main sleep ratio).

Next, we applied this same survival model in males and females separately. Most measures clustered along the line of identity for sex-dependent differences in Figure S 4, suggesting that men and women overall experience similar benefits from physical activity, sleep, and diurnal rhythmicity. A few significant interactions emerged for physical activity (*n*=12) and sleep (n=2) related behavioral health variables (sex_difference_*q*<0.05). These suggested a trend that men have increased mortality risk from low physical inactivity compared to women (for example, moderate_overall physical activity logHR=-8.21 in men compared to logHR=-0.88 in women, *q*=0.001). Similarly, fragmentation of sleep (acceleration_during_main_sleep_mean) associated with higher mortality in men (logHR=0.003 in men, logHR=-0.47 in women, *q*=0.0017).

### Rhythms Predict Future Diagnoses

We hypothesized that diurnal rhythmicity predicts future diagnoses, rather than simply associating with lifetime disease status. To test this, we selected the temp_RA measure of diurnal rhythmicity and performed Cox proportional hazards models with each PheCODE diagnosis as an endpoint. All subjects with the diagnosis occurring prior to actigraphy measures were removed from consideration. Factors were included to control for sex, ethnicity (white/other), self-reported overall health, high household income (≥ 52,000 pounds sterling/year), age at the time of actigraphy, BMI, college education and smoking status, all measured at the initial assessment.

Of 332 PheCODEs with at least 50 cases, 45 PheCODEs reached significance at q<0.05 (Figure 5, Table S 7). The most significant association was in diabetes mellitus, where a two standard deviation increase in temp_RA increased the log hazard ratio by 0.48 corresponding to a 61% (48-76%, 95% CI) increase in rate of diabetes diagnosis. Wrist temperature rhythms also had large predictive power in other diagnoses, see Table 1 and Table S 2.

**Table 1.** Hazard Ratios for select diagnoses

| **Activity Variable** | **Hazard Ratio**<br>**From 1 SD increase**<br>**(95% CI)** | **Hazard Ratio**<br>**From 2 SD increase**<br>**(95% CI)** |
| --- | --- | --- |
| **Diabetes** | 1.27 (1.21 – 1.33) | 1.61 (1.48 – 1.76) |
| **Hypertension** | 1.10 (1.08 – 1.13) | 1.22 (1.16 – 1.27) |
| **Renal Failure** | 1.14 (1.08 – 1.21) | 1.30 (1.16 – 1.46) |
| **Chronic Airway Obstruction** | 1.18 (1.11-1.25) | 1.38 (1.23 – 1.55) |
| **Pneumonia** | 1.14 (1.08 – 1.20) | 1.30 (1.16 – 1.45) |
| **Anxiety Disorders** | 1.13 (1.06 – 1.19) | 1.27 (1.13 – 1.42) |
| **Disorders of Lipoid Metabolism** | 1.09 (1.06 – 1.13) | 1.19 (1.11 – 1.27) |

From the Cox proportional hazards models, wrist temperature rhythms (temp\_RA) were predictive of disease outcomes. Seven of the largest effect sizes are given here, as hazard ratios comparing between mean and 1 SD above average temp\_RA or comparing mean to 2 SD above average. See also Table S 7.

**Figure 5.**
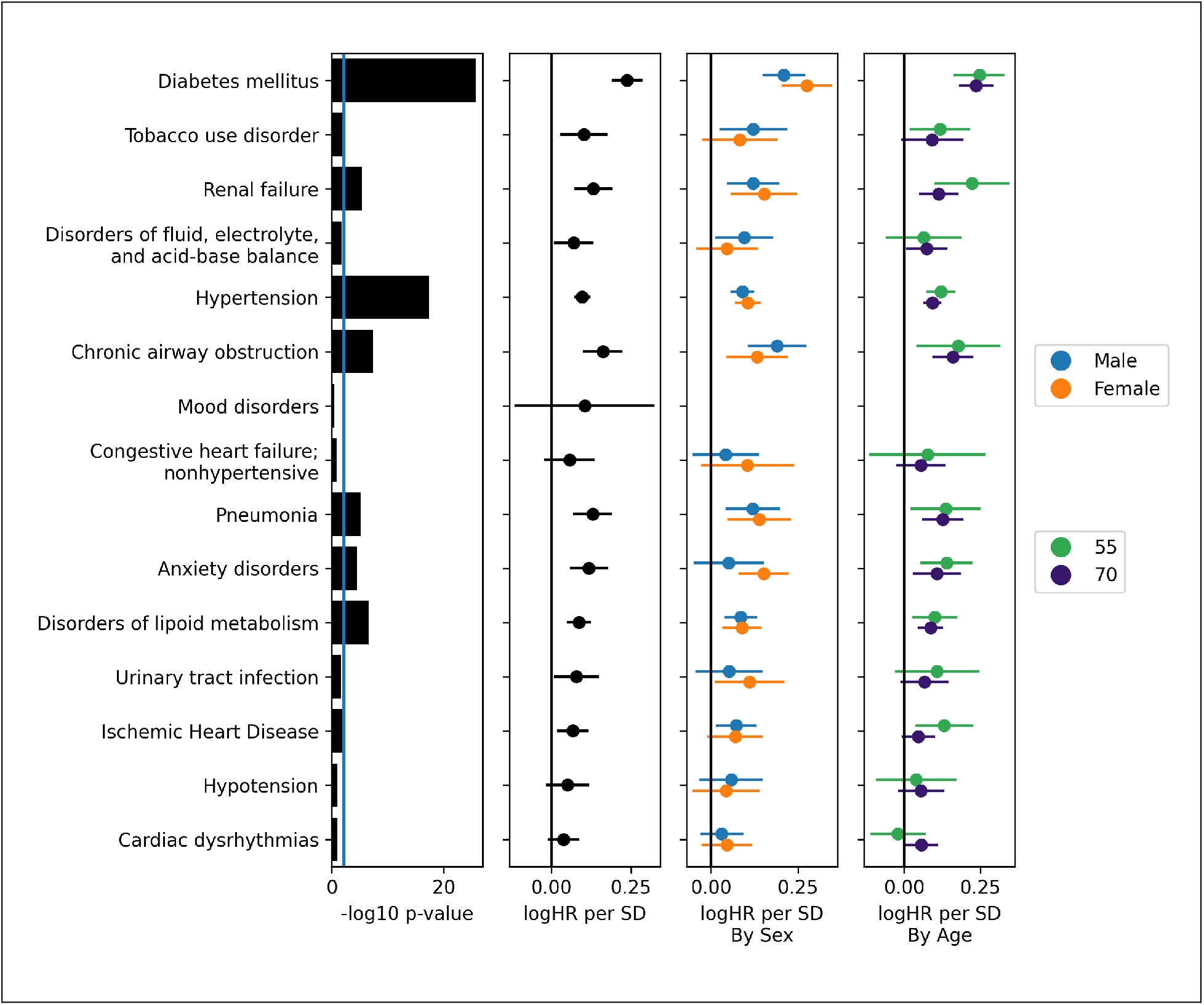
Diurnal Rhythmicity Predict Diagnoses. To test whether diurnal rhythmicity predict future disease, a Cox proportional hazards model was run for each PheCODE testing for effects from the diurnal rhythm measure temp_RA. Individuals with diagnoses prior to the actigraphy measurement were excluded. In many phenotypes, weaker temperature rhythms predict future diagnosis. No phenotypes displayed significant differences by sex or by age in the level of predictive power provided by temp_RA. Left, significance of the overall effect size (not by age or sex). Right three panels, effect sizes by overall model, by sexes, and in younger and older populations. Effect size measured as the log hazard ratio (log HR) per standard deviation (SD) change in the temp_RA measure.

The model was repeated with sex interaction terms to check for differences in males and females and with age interaction terms to check for difference in younger and older populations. No significant sex or age differences were observed (*q* > 0.4 for all PheCODEs), indicating that temperature RA is a robust marker of future disease state in this population.

### Phenome-wide Sex and Age Differences

#### Most PheCODE Associations Consistent Between Sexes

To check for sexual dimorphic effects, associations between PheCODEs and behavioral health measures were computed in each sex for phenotypes with at least 50 cases in both males and females (**Figure 6** a,c, **Figure S 5**). Effect sizes in the two sexes were similar but with significant differences in the digestive and mental disorders. Among 38,316 disease phenotype and behavioral measure pairs, only 54 reached a significant difference between men and women (*q*-diff≤0.05, Table S 4). See also Supplemental Results.

**Figure 6.**
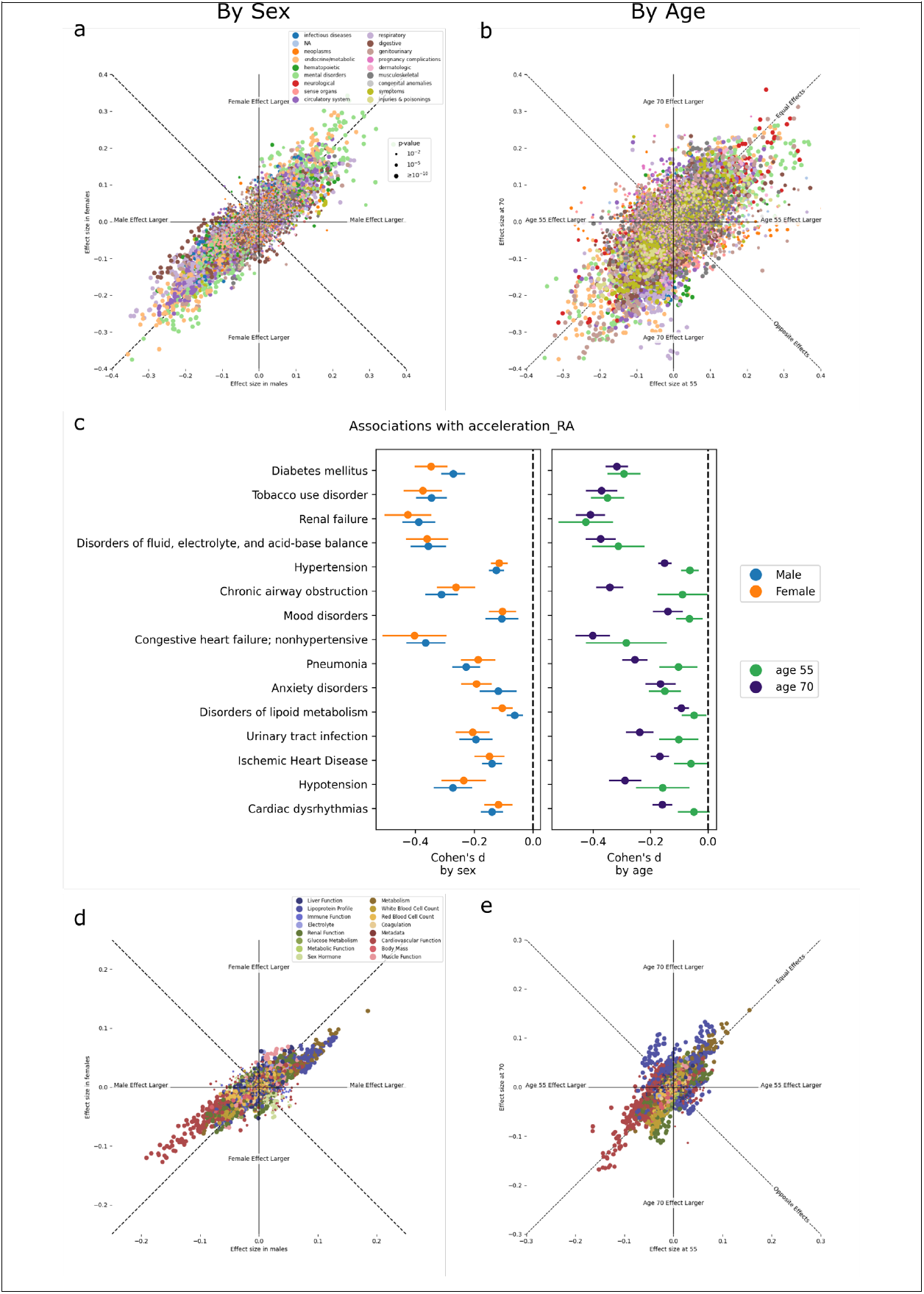
Sex- and Age-Specific Effects. Comparison of associations between medical diagnoses and behavioral health measures by sex and age. (a, b) Comparison of the sex-specific effect sizes for behavioral health measure (difference in behavioral health measure or quantitative trait between cases and controls, normalized by the standard deviation of the behavioral health measure) in (a) males versus females and (b) in the younger (age 55) and older (age 70) populations by phenotype category. Dashed lines represent equal or opposite effect sizes between the two groups. (c) Effect sizes in males versus females and by age in select categories of phenotypes, associating with circadian rhythm robustness (measured as acceleration_RA). Lines denote 95% confidence intervals. (d-e) Comparison of sex-specific effect sizes for quantitative traits (SD difference in quantitative trait per SD change in behavioral measure) in (d) males and females and in (e) younger and older populations, by quantitative trait category.

#### Physical activity shows stronger associations with quantitative phenotypes in males than females

Among the quantitative traits, a strong sex-dependence was observed (Figure **6** d, Table S 5) with 944 associations displaying sexual dimorphism where males have the larger effect sizes in 81% of those associations. Effect sizes were measured as Pearson *r* correlations (using each sex’s individual variations) and so these differential effects were not driven by differences in variability in the sexes. The actigraphy health variables driving these changes are primarily those reflecting physical activity levels (Figure S 6) and are consistent in both the objectively measured activity level and the IPAQ activity group derived from self-report. These sex-dependent effects were seen in a wide range of quantitative traits including vitamin D, physical measures (grip strength, waist circumference), creatinine, cardiovascular measures (pulse rate, apolipoprotein A and triglycerides) (*q*<10^−5^).

#### Age Interacts Significantly with Physical Activity, Sleep, and Diurnal Rhythmicity and PheCODEs

Next, we examined age-dependent differences. This model was performed among phenotypes with at least 200 cases and age (at the time actigraphy was taken) was included as an interaction term with case/control status. Here, 1,397 out of 39,432 pairs of disease phenotype and actigraphy health variable are significant at the *q*≤0.05 level (Table S 4). This demonstrated significant differences between older and younger populations in a subset of diagnoses. To illustrate this, in Figure 6 b,c, effect size estimates at age 55 were plotted against estimates at age 70, which corresponded approximately to the 20^th^ and 80^th^ percentile ages in this population, respectively. Hypertension, osteoarthrosis, chronic airway obstruction, anxiety and mood disorders showed larger effect sizes in the younger populations. Disorders of menstruation and varicose veins had larger effects in the younger populations. See Supplemental Results.

#### Age-Dependent Patterns in Quantitative Traits Associated with Physical Activity

Age dependency in the association between quantitative trait and actigraphy health variables were seen in 20% (2,109 out of 10,292 tests) at a level of *q*≤0.05. Top candidates almost exclusively belonged to the cardiovascular system, lipoid metabolism, body mass and kidney function (Figure **6** e). Results were consistent with or without controlling for medication, except in some lipoprotein profiles while controlling for cholesterol lowering medications (see Supplemental Results).

## Discussion

Many chronic disease conditions are associated with disrupted biological rhythms [27]. By applying our PheWAS-SensR approach to the UKB, we show that actigraphy data strongly associates with PheCODES. Overall exercise and sleep behaviors correlate well with disease, biomarkers of organ function and health, and outcomes. We find that these relationships are modulated little by sex but substantially by age.

In a novel approach for which we quantified diurnal behavior in two distinct ways, actigraphy-based (diurnal rhythmicity) and skin temperature-based (circadian rhythmicity), we demonstrated that these rhythms associate substantially with disease, survival outcomes, and quantitative biomarkers of health and organ function. These effects were similar in size to effects from exercise and slumber, underscoring the relevance of robust rhythms for health. Hypertension, diabetes mellitus, pneumonia and anxiety disorders are exemplary PheCODES for these relationships. Diabetes mellitus had strong correlations with both physical activity and diurnal rhythmicity. Interestingly, the levels of glycated hemoglobin were less associated with exercise but significantly so with rhythmicity. This raises the question of whether the structured lifestyle interventions used to treat diabetes [28] confer part of their benefit on glycemic control through re-establishing rhythmic behavioral patterns. Our analyses show that robustness of diurnal rhythmicity is an important determinant of health status, and that deconsolidation increases disease risk separate from physical activity and sleep. Therefore, it not only matters for our wellbeing that we exercise and sleep well, but also the regularity of timing that we are engaged in these activities.

A key question is which disease onsets are predicted by deconsolidation of rhythmicity. Here, we find that weaker circadian rhythms of skin temperature in healthy participants at the time of actigraphy had a higher risk for a future diagnosis of diabetes mellitus, hypertension, asthma, chronic airway obstruction, lipid disorders, chronic liver disease, renal failure, pneumonia, and anxiety disorders. This suggests that longitudinal monitoring of diurnal rhythmicity has prognostic value for disease progression. These hazard ratios were not modulated by sex or age in our modeling, suggesting this as a robust circadian marker amongst those older than 50. In contrast, our PheWAS-SensR associations demonstrated significant age modulation. For example, temp_RA had a stronger association with hypertension among younger individuals than older, but its predictive power was not significantly affected by age. However, the predictive models have lower sample sizes and lower power than the associative scores, though sample sizes are expected to be higher when re-run in the future.

The UKB population is not a random sample, being older, overwhelmingly white, and in better overall health than the general population. For actigraphy, the large number of retirees is significant and may affect generalizability of results to younger, more diverse populations. However, the older population also enriches the study for many pathological phenotypes of interest.

The increasing ease of wearable technology facilitates incorporation into large, diverse cohorts, such as the *All of Us* research program [29]. While smartphone-based approaches are easily scalable, it is still unclear how much this digital phenotyping, where sleep time data may be limited, can provide insight into diurnal rhythmicity [30].

In conclusion, our PheWAS-SensR quantifies physical activity, sleep, and rhythms from remote sensors to model disease trajectories with strong associations to survival and powerful predictions for future disease onset. Integration of long-term remote biosensing into patient care could thus afford an individualized approach to risk management.

## Supporting information

Table S 3 - Main results

Table S 4 - PheWAS-SensR results

Table S 5 - PheWAS-SensR quantitative results

Table S 7 - Three Components results

Table S 8 - Proportional Hazards Results

## Data Availability

All data produced are provided in supplementary data files. Underlying data is available from the UK Biobank.

## Acknowledgments

TGB, NFL, and GRG received funding from the National Center for Advancing Translational Sciences Grant (5UL1TR000003). TGB received funding from the National Institute of Mental Health (NIMH) T32 Training grant (5T32MH106442-04). CS is the Robert L. McNeil Jr. Fellow in Translational Medicine and Therapeutics. GAF is the recipient of an AHA Merit award and the McNeil Professor of Translational Medicine and Therapeutics. YIS received funding from NIH/NIMH grants (R01MH098260 and U01MH109991). This research has been conducted using the UK Biobank Resource under Application Number 50398. The authors would like to acknowledge Dr. Harry R. Kennard for providing us advice and code for processing temperature values from the UKB.

GAF is an advisor to Calico Laboratories. CS is an advisor to Antibe Therapeutics Inc. The authors declare no other conflicts of interest.

This paper contains results first presented in posters (titled “Phenome-Wide Association Study of Actigraphy in the UK Biobank”) at the Cold Spring Harbor Laboratory Symposium on Quantitative Biology, Biological Time Keeping conference on June 1 - 5, 2021 and at the Mid-Atlantic Bioinformatics and the MidAtlantic Bioinformatics Conference on Nov 8, 2021.

## Notes

### Author Declarations

Data for this study was provided by the UK Biobank under Application Number 50398 and is available at ukbiobank.ac.uk.

## References

1. Gallagher, M.D. and A.S. Chen-Plotkin, The Post-GWAS Era: From Association to Function. Am J Hum Genet, 2018. 102(5): p. 717–730.

2. Denny, J.C., et al., PheWAS: demonstrating the feasibility of a phenome-wide scan to discover gene-disease associations. Bioinformatics, 2010. 26(9): p. 1205–10.

3. Klarin, D., et al., Genetics of blood lipids among ∼300,000 multi-ethnic participants of the Million Veteran Program. Nat Genet, 2018. 50(11): p. 1514–1523.

4. Ekelund, U., et al., Dose-response associations between accelerometry measured physical activity and sedentary time and all cause mortality: systematic review and harmonised meta-analysis. BMJ, 2019. 366: p. l4570.

5. Casiraghi, L., et al., Moonstruck sleep: Synchronization of human sleep with the moon cycle under field conditions. Sci Adv, 2021. 7(5).

6. Zinkhan, M. and J.W. Kantelhardt, Sleep Assessment in Large Cohort Studies with High-Resolution Accelerometers. Sleep Med Clin, 2016. 11(4): p. 469–488.

7. Burton, C., et al., Activity monitoring in patients with depression: a systematic review. J Affect Disord, 2013. 145(1): p. 21–8.

8. Doherty, A., et al., Large Scale Population Assessment of Physical Activity Using Wrist Worn Accelerometers: The UK Biobank Study. PLoS One, 2017. 12(2): p. e0169649.

9. Lyall, L.M., et al., Association of disrupted circadian rhythmicity with mood disorders, subjective wellbeing, and cognitive function: a cross-sectional study of 91 105 participants from the UK Biobank. Lancet Psychiatry, 2018. 5(6): p. 507–514.

10. Ferguson, A., et al., Genome-Wide Association Study of Circadian Rhythmicity in 71,500 UK Biobank Participants and Polygenic Association with Mood Instability. EBioMedicine, 2018. 35: p. 279–287.

11. Willetts, M., et al., Statistical machine learning of sleep and physical activity phenotypes from sensor data in 96,220 UK Biobank participants. Sci Rep, 2018. 8(1): p. 7961.

12. Sarabia, J.A., et al., Circadian rhythm of wrist temperature in normal-living subjects A candidate of new index of the circadian system. Physiol Behav, 2008. 95(4): p. 570–80.

13. Wu, P., et al., Mapping ICD-10 and ICD-10-CM Codes to Phecodes: Workflow Development and Initial Evaluation. JMIR Med Inform, 2019. 7(4): p. e14325.

14. Wei, W.Q., et al., Evaluating phecodes, clinical classification software, and ICD-9-CM codes for phenome-wide association studies in the electronic health record. PLoS One, 2017. 12(7): p. e0175508.

15. Fry, A., et al., Comparison of Sociodemographic and Health-Related Characteristics of UK Biobank Participants With Those of the General Population. Am J Epidemiol, 2017. 186(9): p. 1026–1034.

16. Witting, W., et al., Alterations in the circadian rest-activity rhythm in aging and Alzheimer’s disease. Biol Psychiatry, 1990. 27(6): p. 563–72.

17. Mantri, S., et al., Physical Activity in Early Parkinson Disease. J Parkinsons Dis, 2018. 8(1): p. 107–111.

18. Schutte-Rodin, S., et al., Clinical guideline for the evaluation and management of chronic insomnia in adults. J Clin Sleep Med, 2008. 4(5): p. 487–504.

19. Cuesta, M., et al., Skin Temperature Rhythms in Humans Respond to Changes in the Timing of Sleep and Light. J Biol Rhythms, 2017. 32(3): p. 257–273.

20. Kennard, H.R., et al., The associations between thermal variety and health: Implications for space heating energy use. PLoS One, 2020. 15(7): p. e0236116.

21. Capella, M.D.M., A. Martinez-Nicolas, and A. Adan, Circadian Rhythmic Characteristics in Men With Substance Use Disorder Under Treatment. Influence of Age of Onset of Substance Use and Duration of Abstinence. Front Psychiatry, 2018. 9: p. 373.

22. Lack, L., et al., Chronotype differences in circadian rhythms of temperature, melatonin, and sleepiness as measured in a modified constant routine protocol. Nat Sci Sleep, 2009. 1: p. 1–8.

23. Monk, T.H., et al., Effects of afternoon “siesta” naps on sleep, alertness, performance, and circadian rhythms in the elderly. Sleep, 2001. 24(6): p. 680–7.

24. Bluher, M., Adipose tissue dysfunction in obesity. Exp Clin Endocrinol Diabetes, 2009. 117(6): p. 241–50.

25. Merghani, A., A. Malhotra, and S. Sharma, The U-shaped relationship between exercise and cardiac morbidity. Trends Cardiovasc Med, 2016. 26(3): p. 232–40.

26. Hou, C., et al., Association of sleep duration with risk of all-cause mortality and poor quality of dying in oldest-old people: a community-based longitudinal study. BMC Geriatr, 2020. 20(1): p. 357.

27. Fitzgerald, G.A., et al., Molecular clocks and the human condition: approaching their characterization in human physiology and disease. Diabetes Obes Metab, 2015. 17 Suppl 1: p. 139–42.

28. Colberg, S.R., et al., Physical Activity/Exercise and Diabetes: A Position Statement of the American Diabetes Association. Diabetes Care, 2016. 39(11): p. 2065–2079.

29. All of Us Research Program, I., et al., The “All of Us” Research Program. N Engl J Med, 2019. 381(7): p. 668–676.

30. Onnela, J.P., Opportunities and challenges in the collection and analysis of digital phenotyping data. Neuropsychopharmacology, 2021. 46(1): p. 45–54.

