## Supplementary figures and images for "Phenome-Wide Association Study of Actigraphy in the UK Biobank"

### age_effects.2x2.png

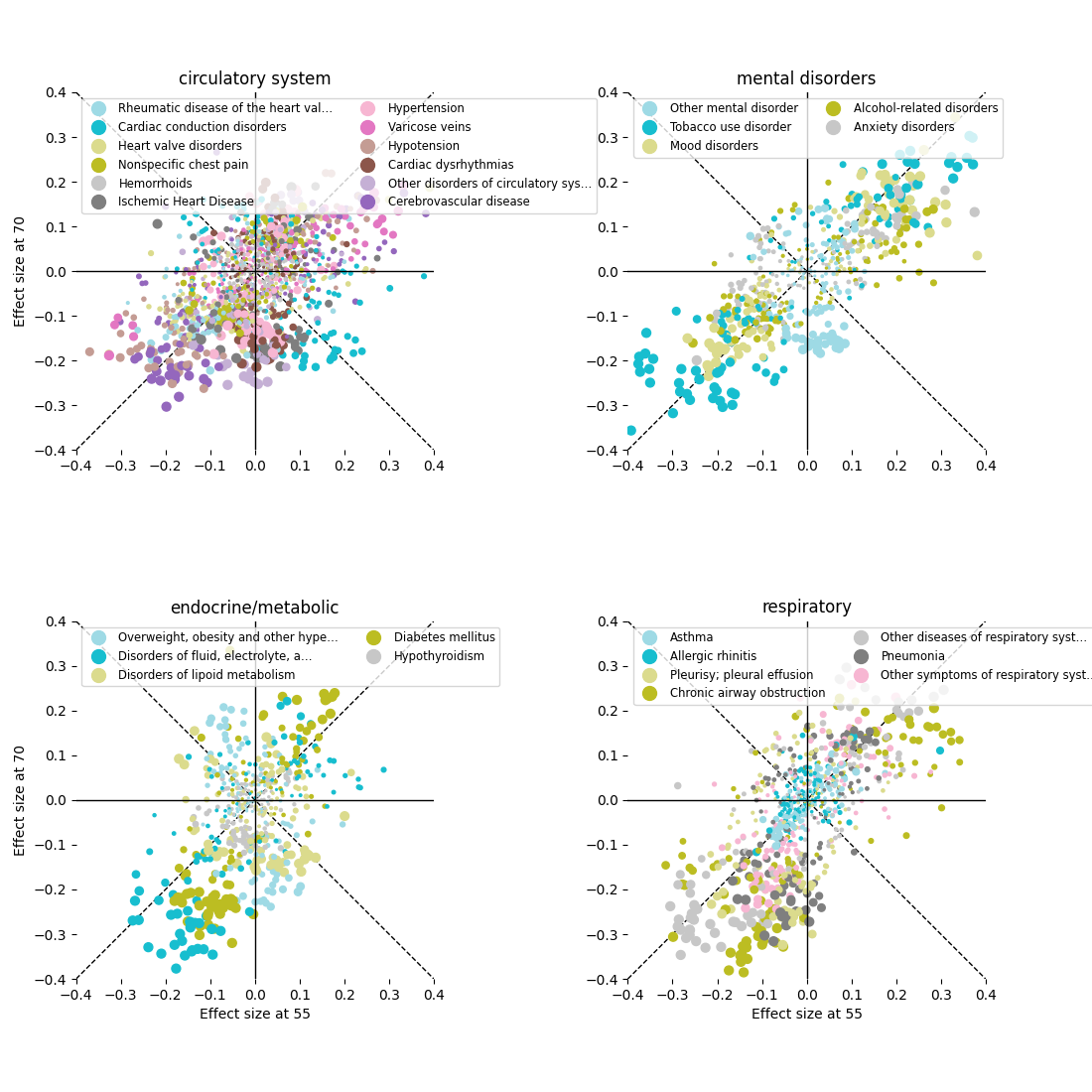

### age_effects.png

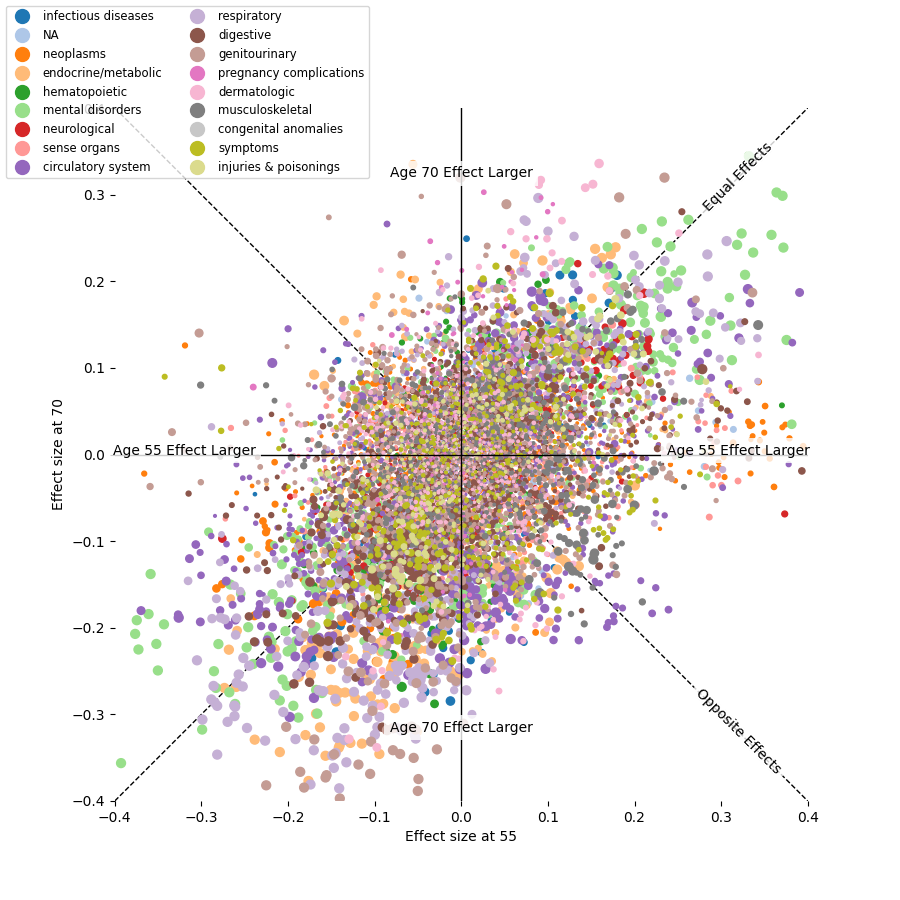

### age_effects.quantitative.2x2.png

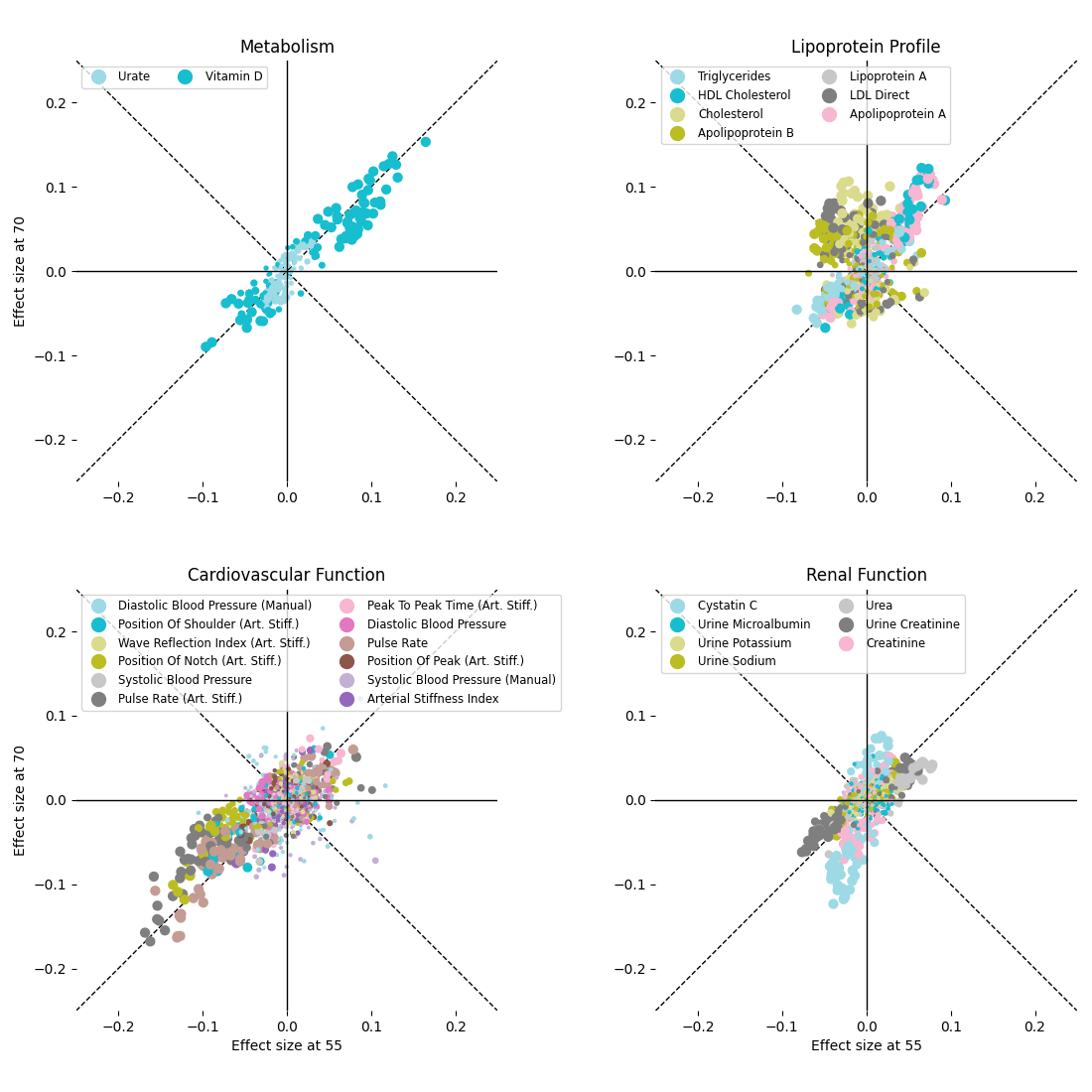

### age_effects.quantitative.png

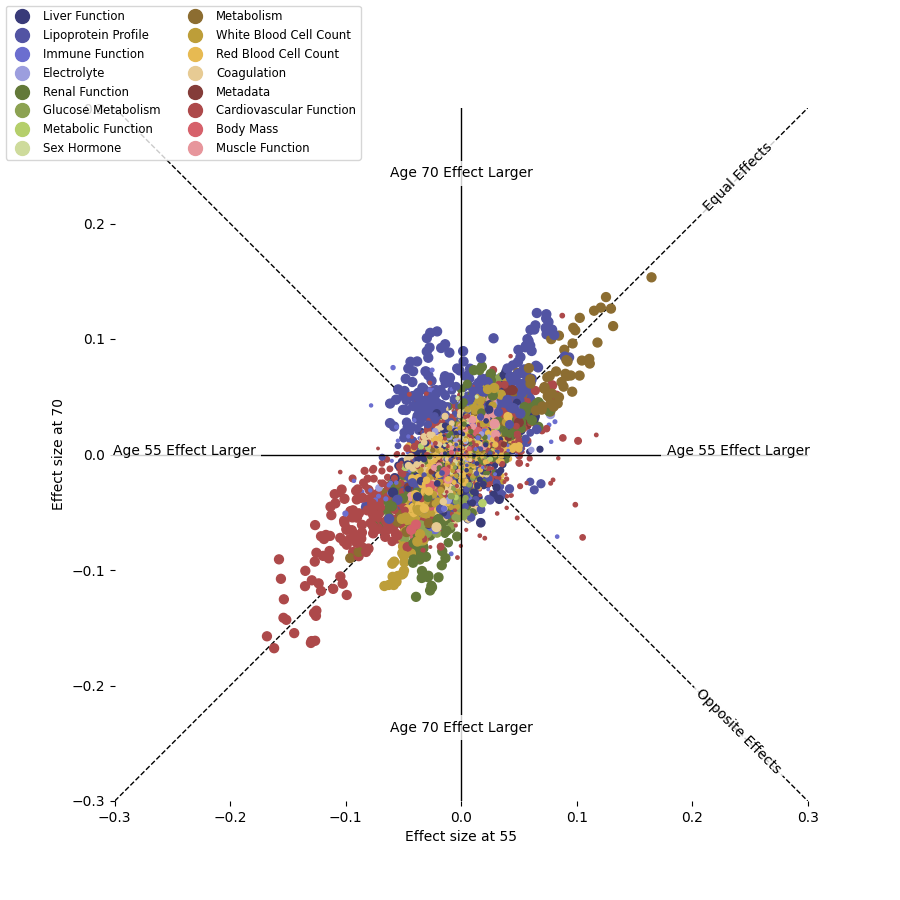

### age_effects.with_meds.quantitative.lipoproteins.png

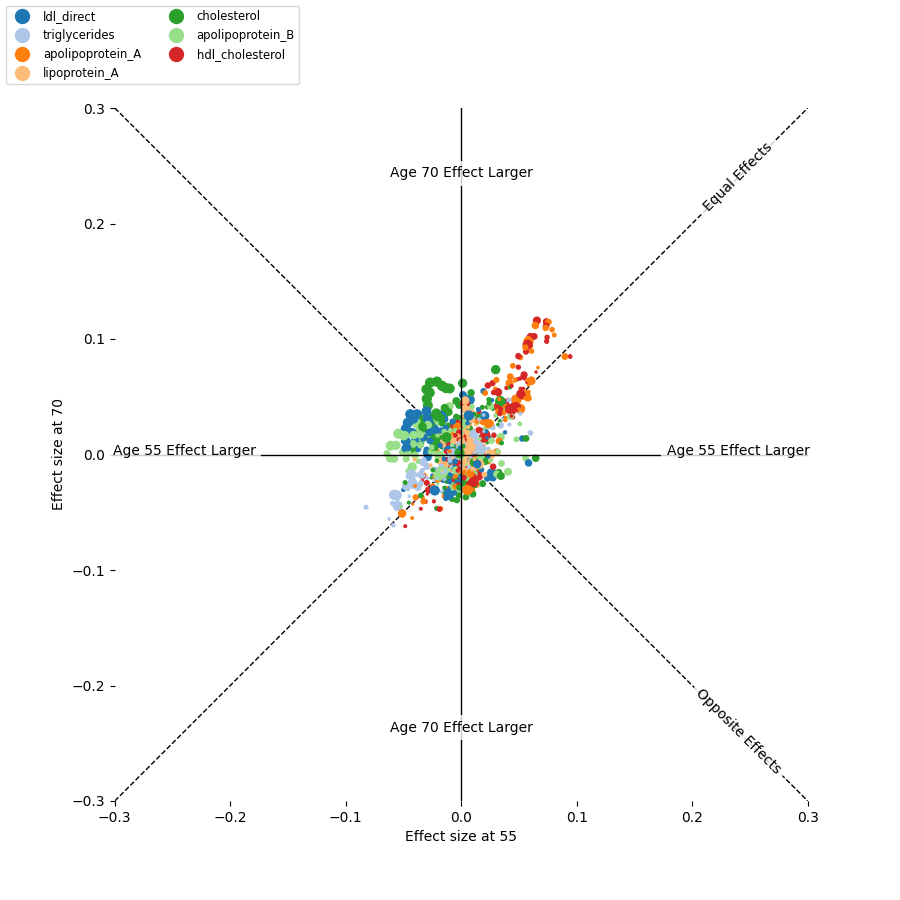

### age_effects.with_meds.quantitative.png

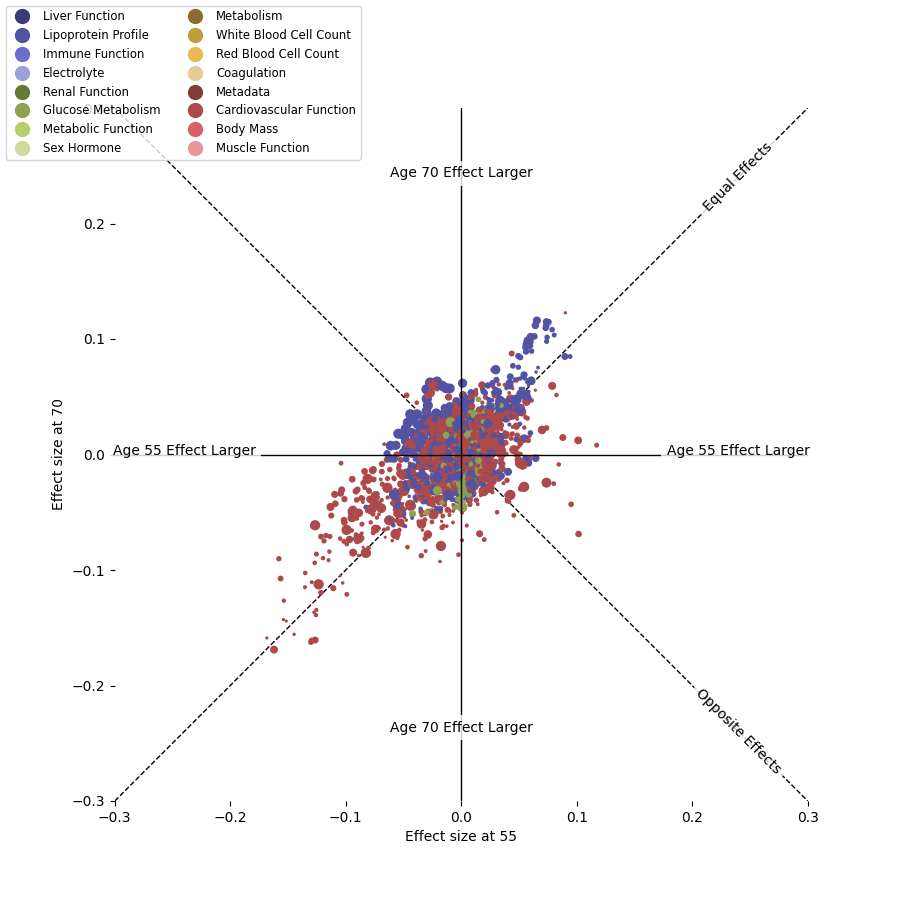

### by_age.by_sex.ldl_direct.combined.all.png

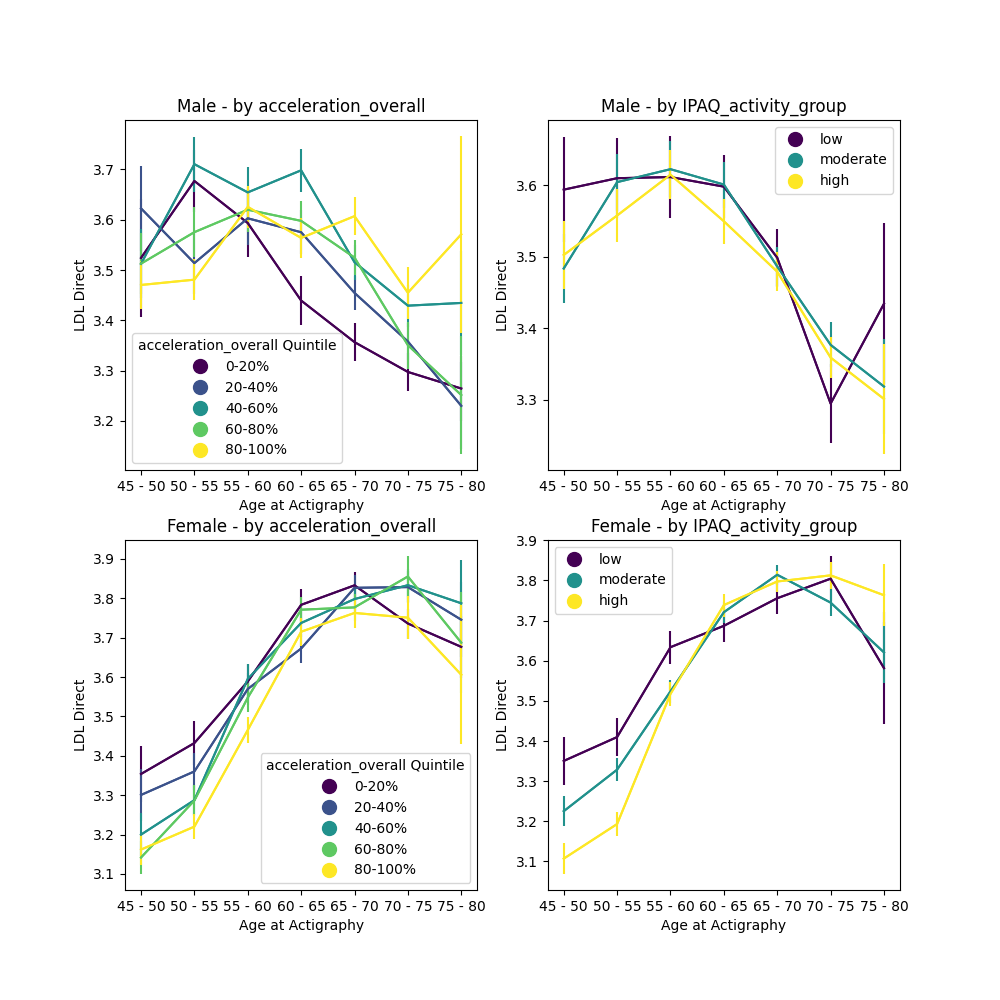

### by_age.by_sex.ldl_direct.combined.no_meds.png

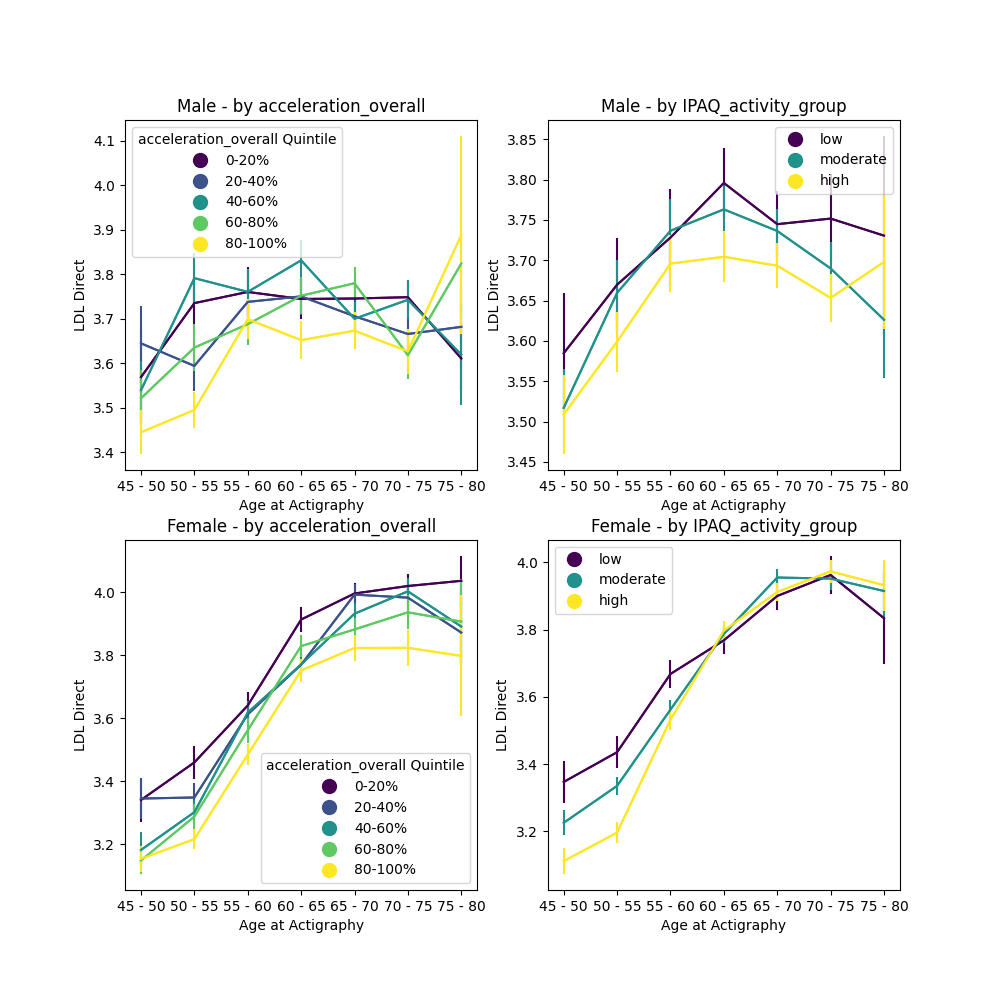

### by_age.by_sex.ldl_direct.combined.with_meds.png

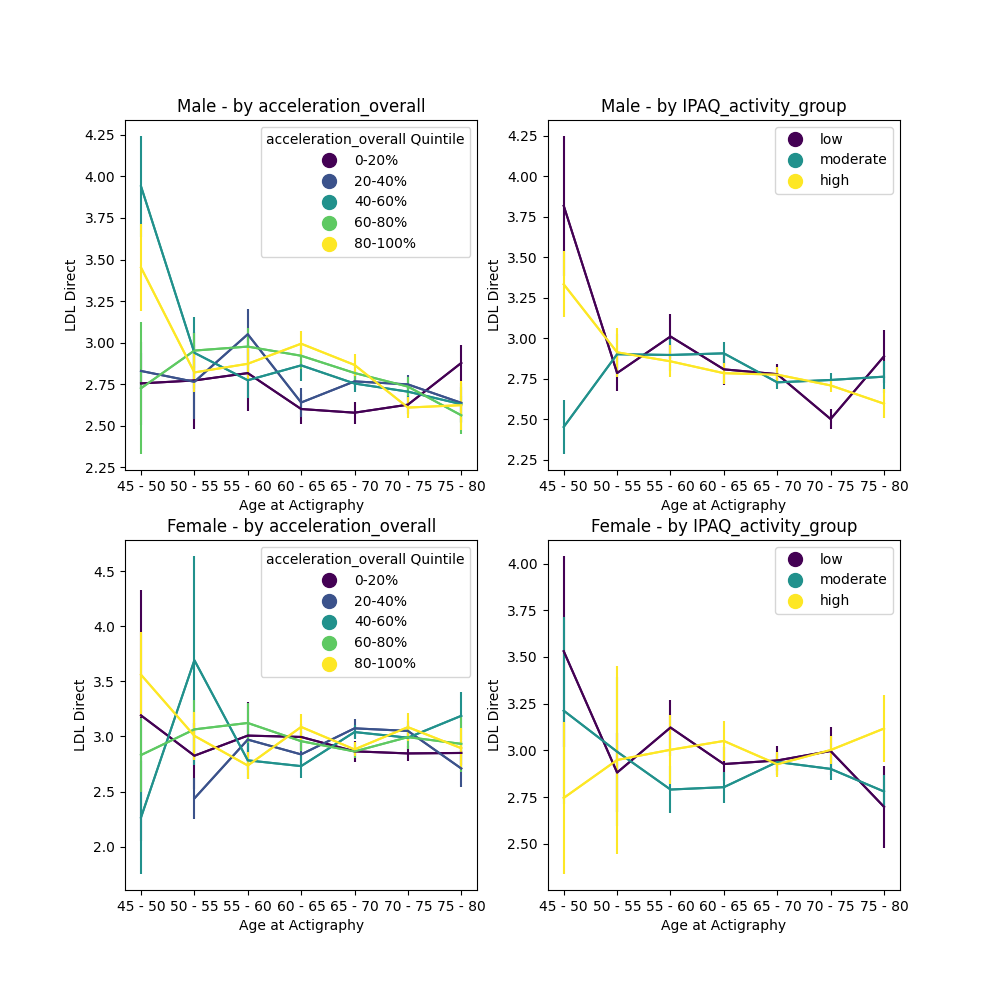

### by_age.creatinine.vs.acceleration_overall.png

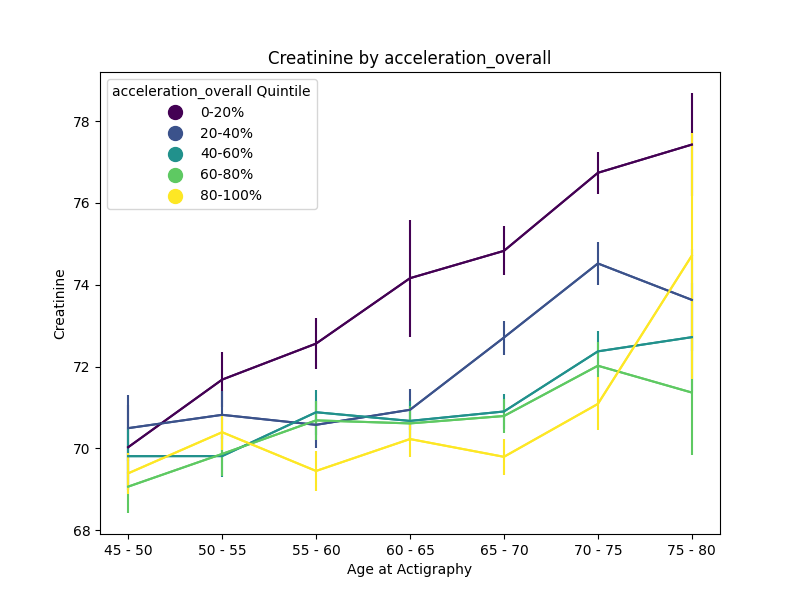

### by_age.cystatin_C.vs.acceleration_overall.png

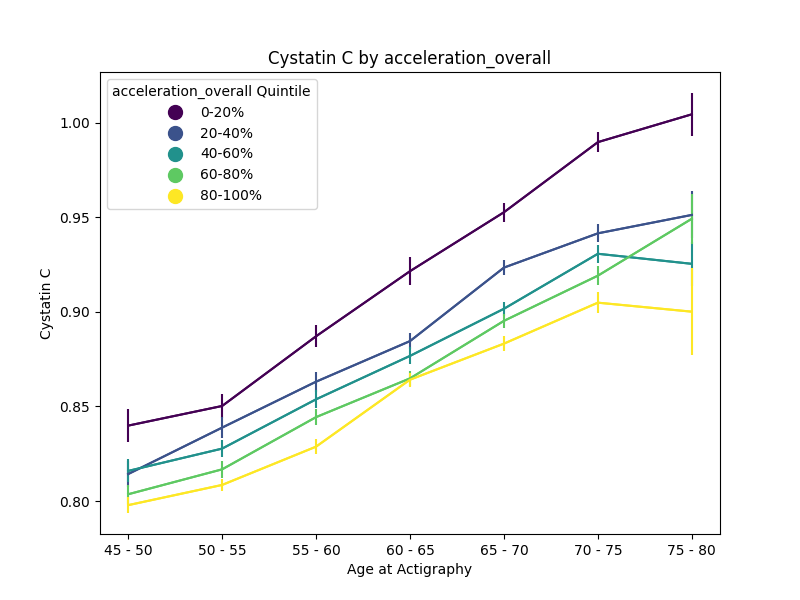

### by_age.urine_creatinine.vs.acceleration_overall.png

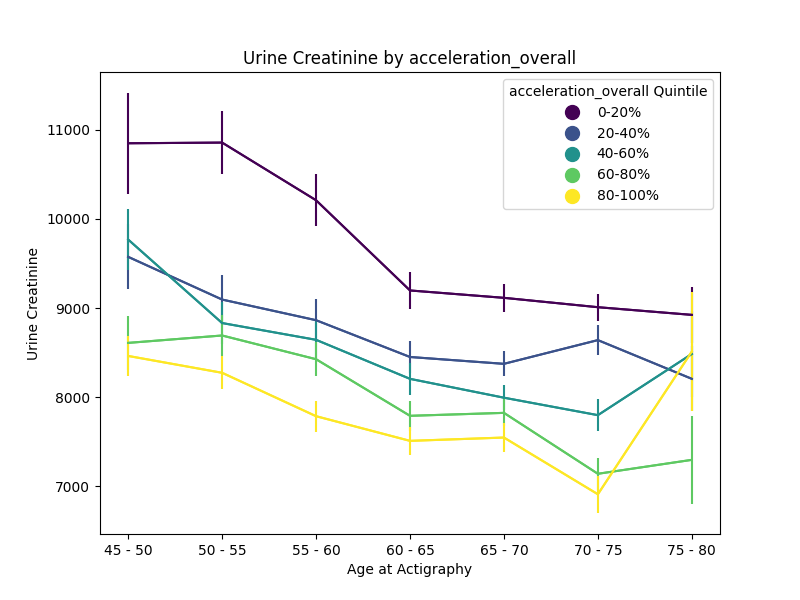

### cholesterol.by_age.hdl_cholesterol.vs.acceleration_overall.png

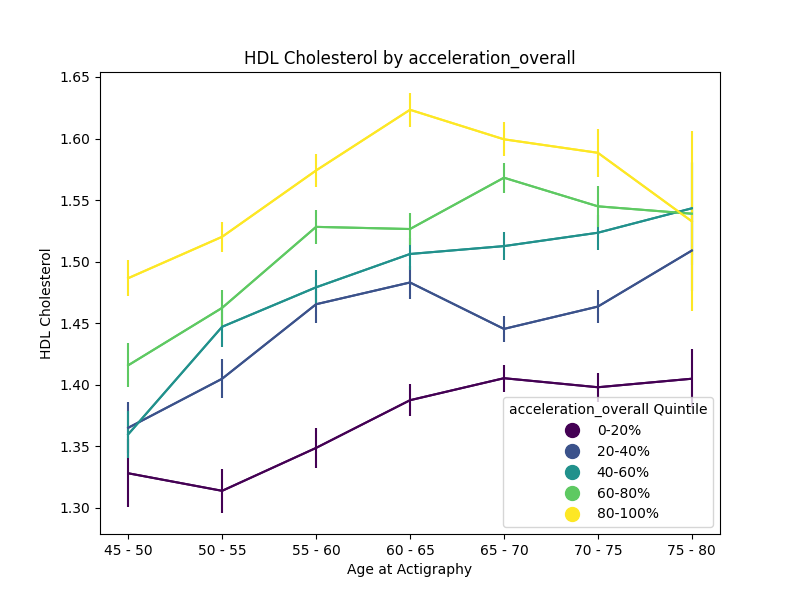

### cholesterol.by_age.lipoprotein_A.vs.acceleration_overall.png

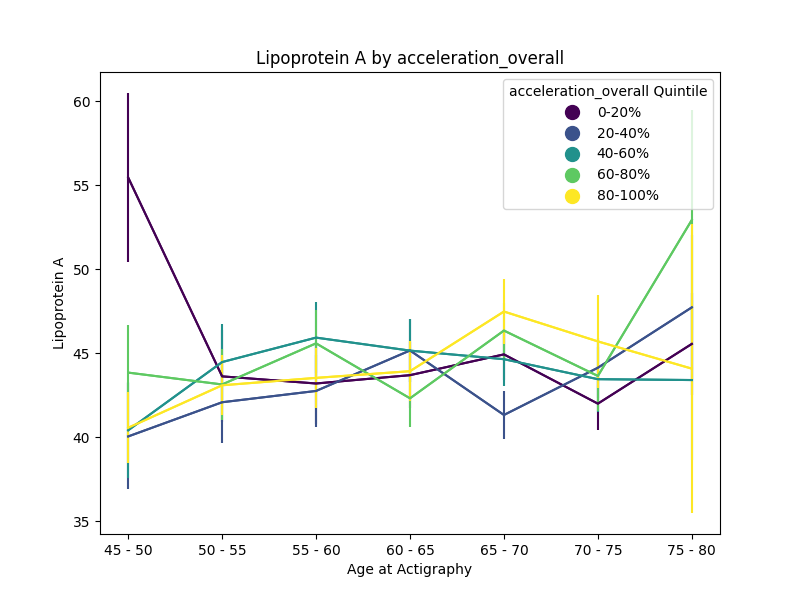

### cholesterol.by_age.triglycerides.vs.acceleration_overall.png

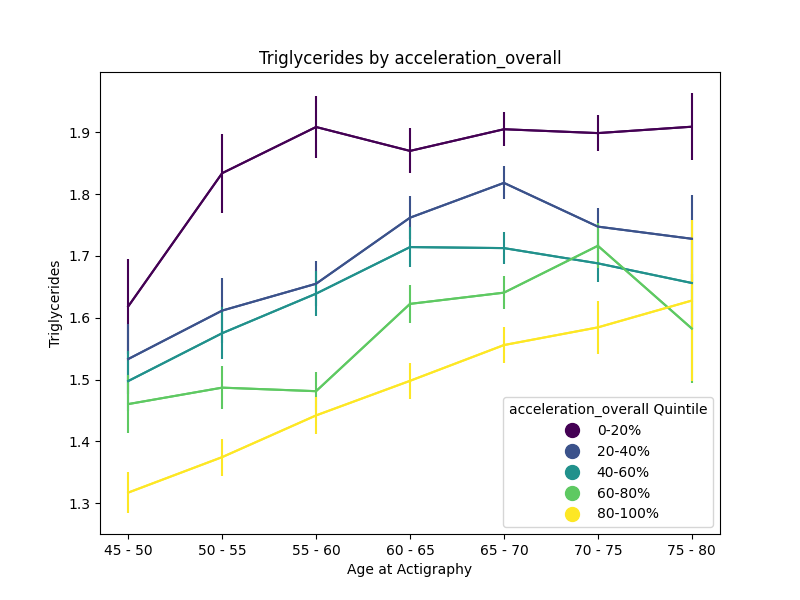

### FIG1.manhattan_plot.png

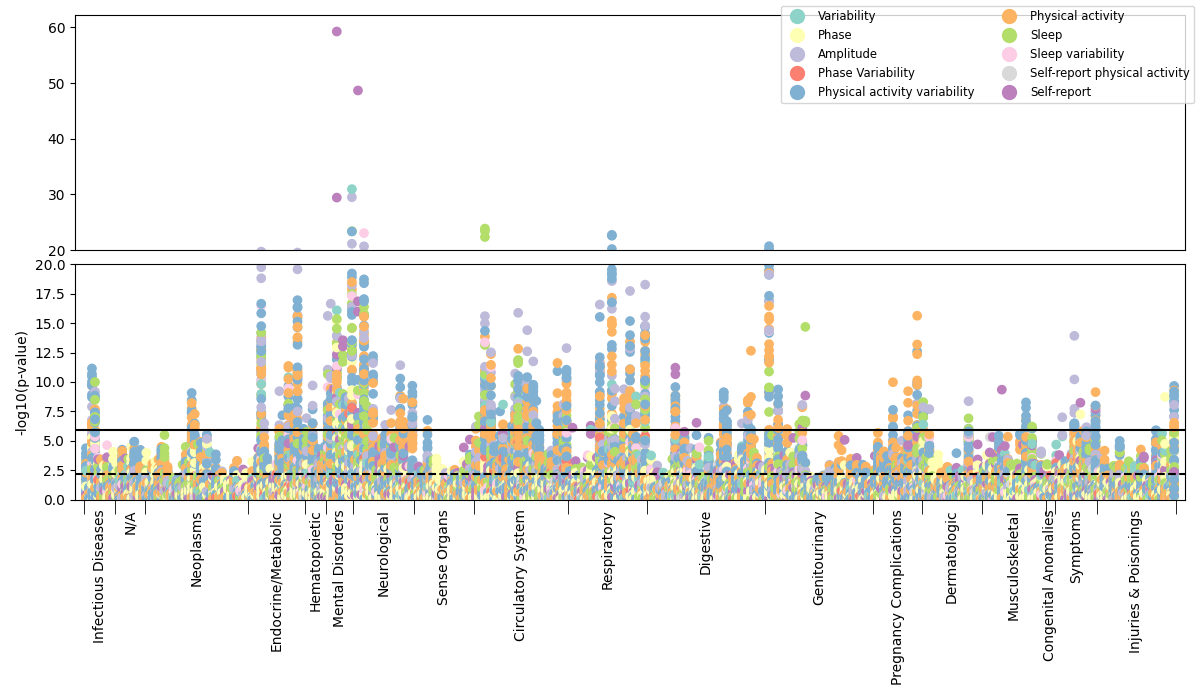

### FIG1.manhattan_plot.quantitative.png

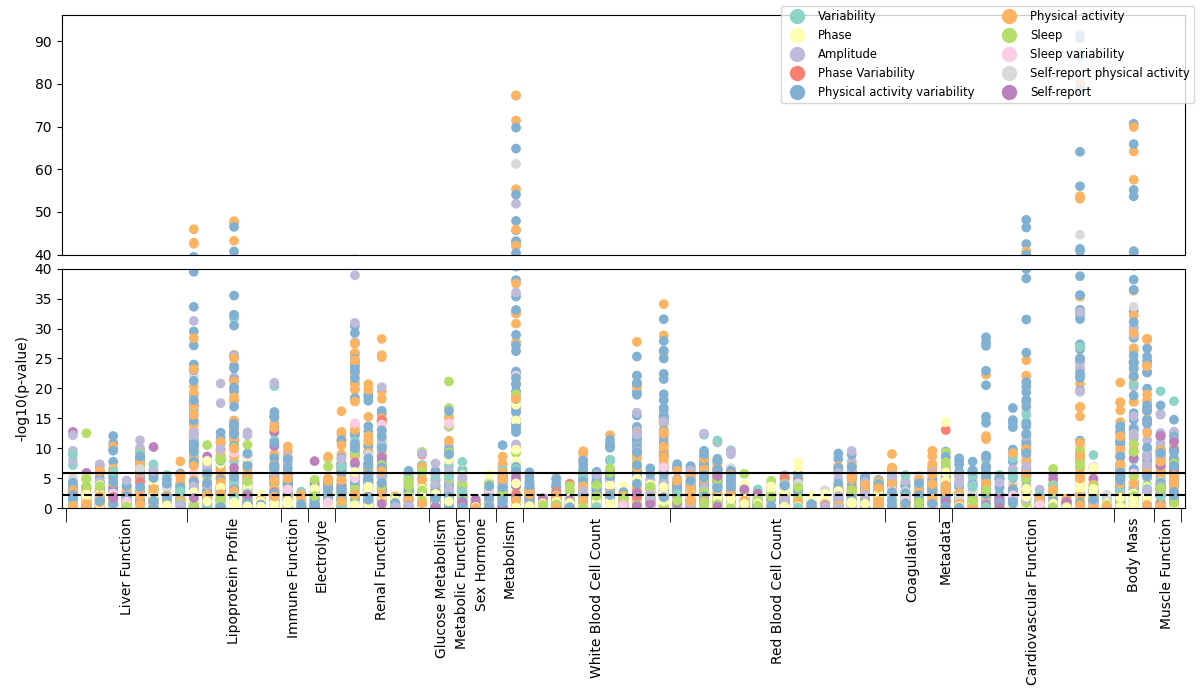

### FIG1.phenotypes.diabetes.png

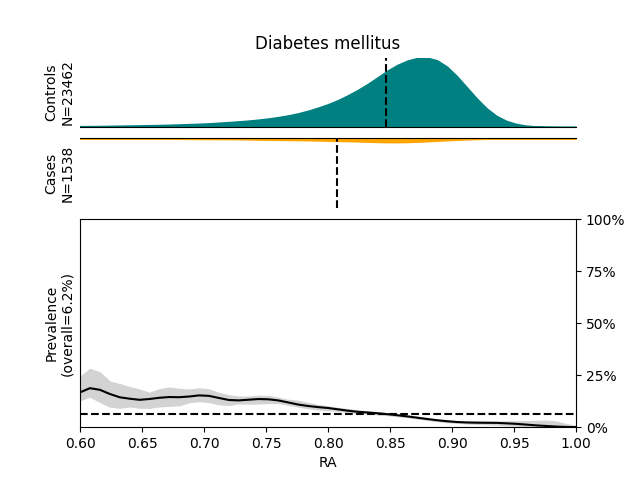

### FIG1.phenotypes.hypertension.png

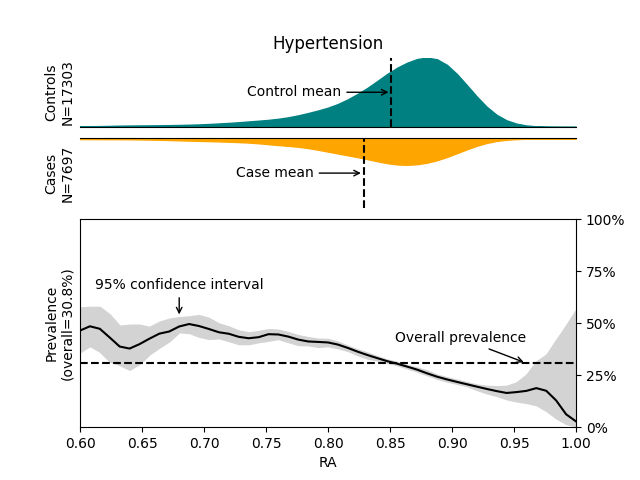

### FIG1.phenotypes.parkinsons.png

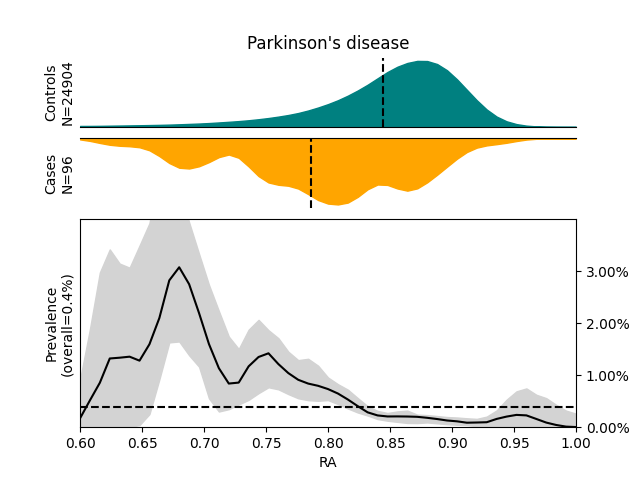

### FIG1.phenotypes.renal_failure.png

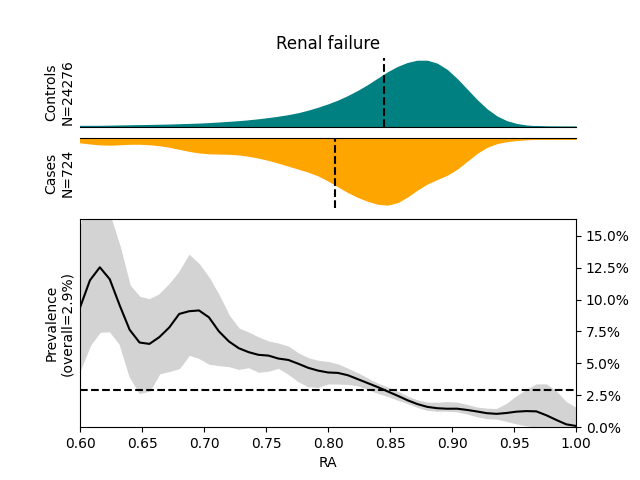

### FIG3.circadian_vs_other_vars.png

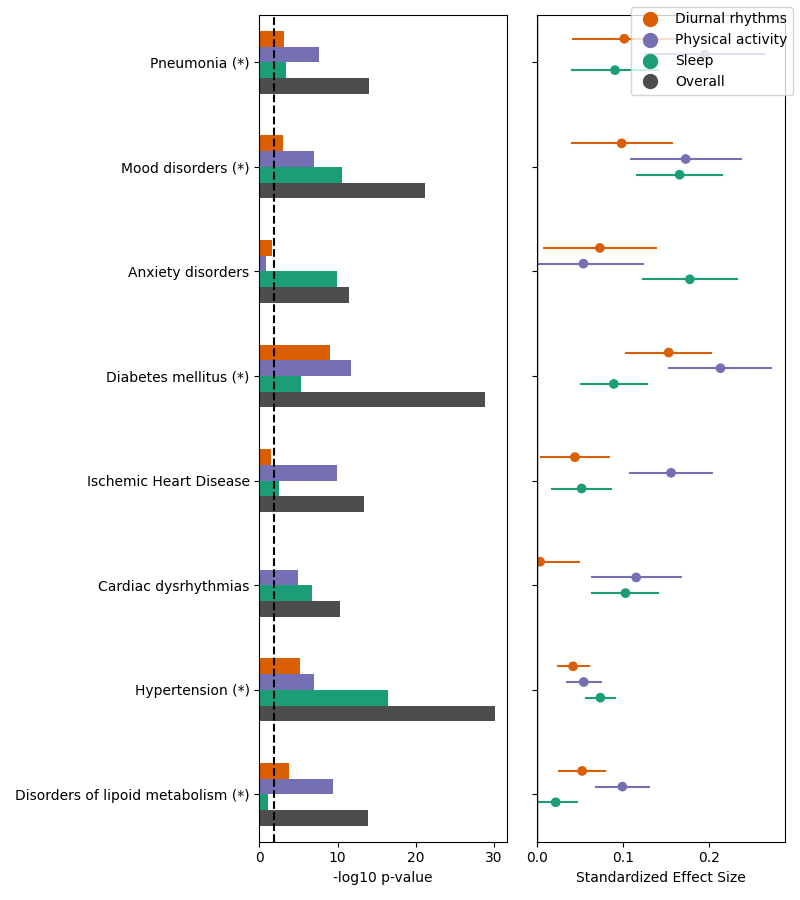

### FIG3.circadian_vs_other_vars.quantitative.temp_amplitude.png

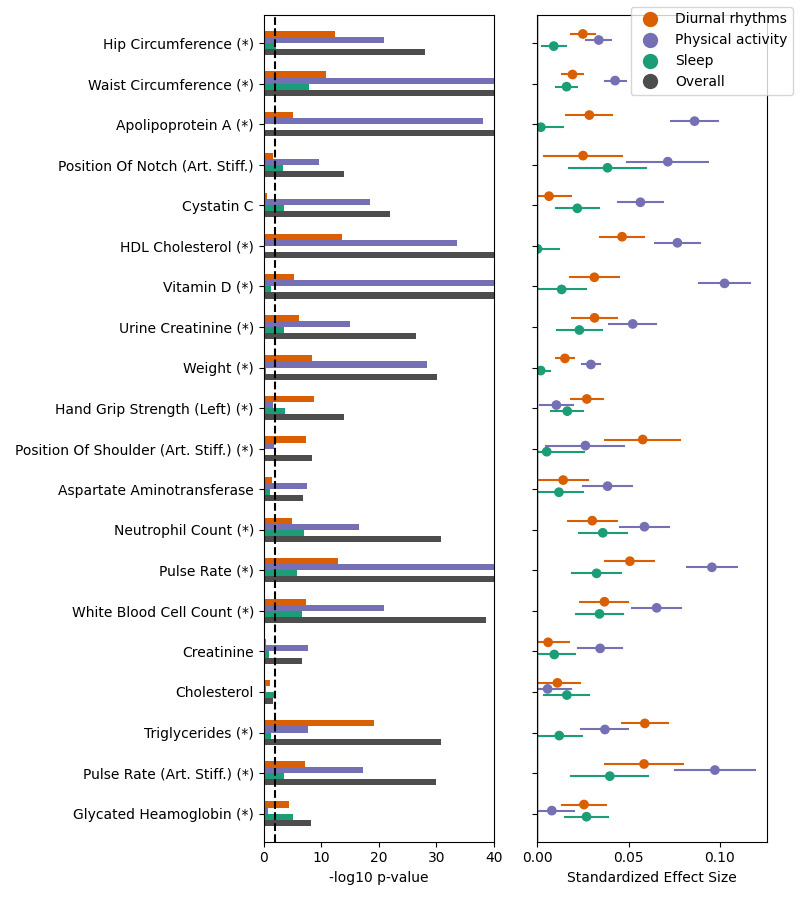

### FIG4.survival.acceleration_overall.png

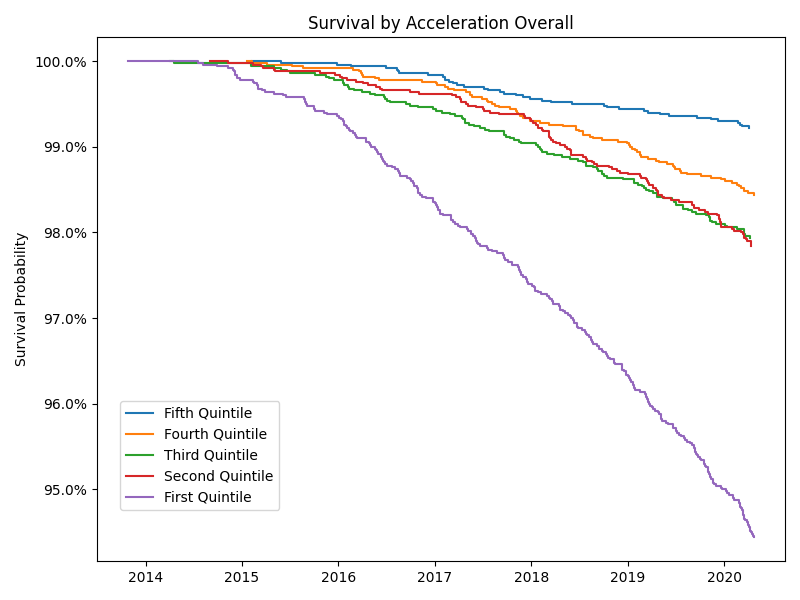

### FIG4.survival.main_sleep_ratio_mean.png

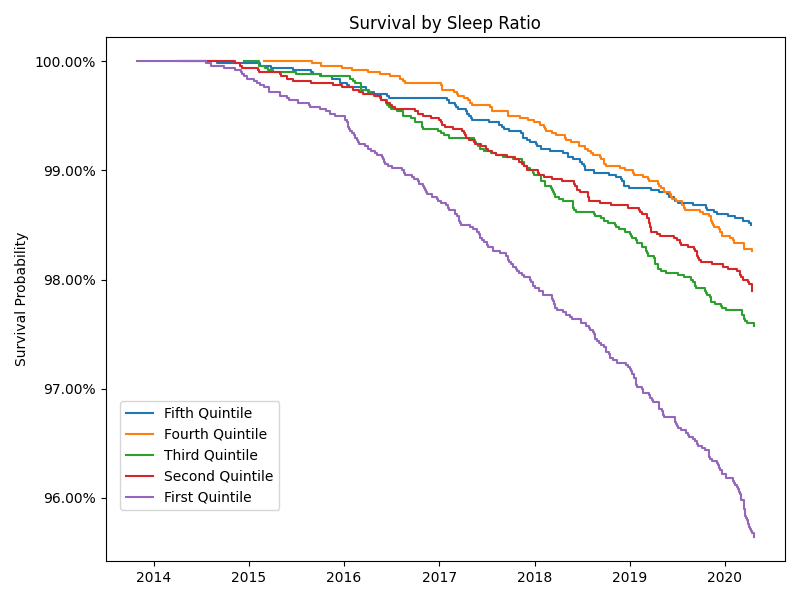

### FIG4.survival.RA.png

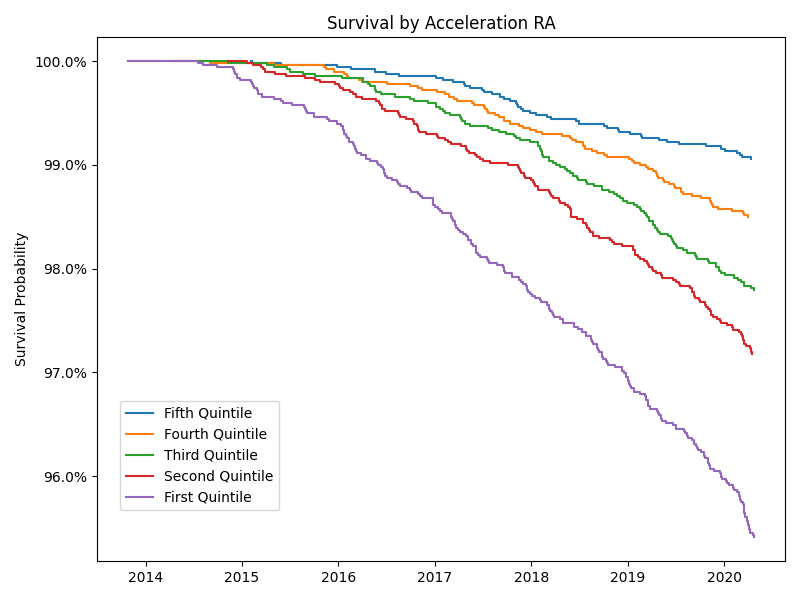

### FIG5.predictive_tests.png

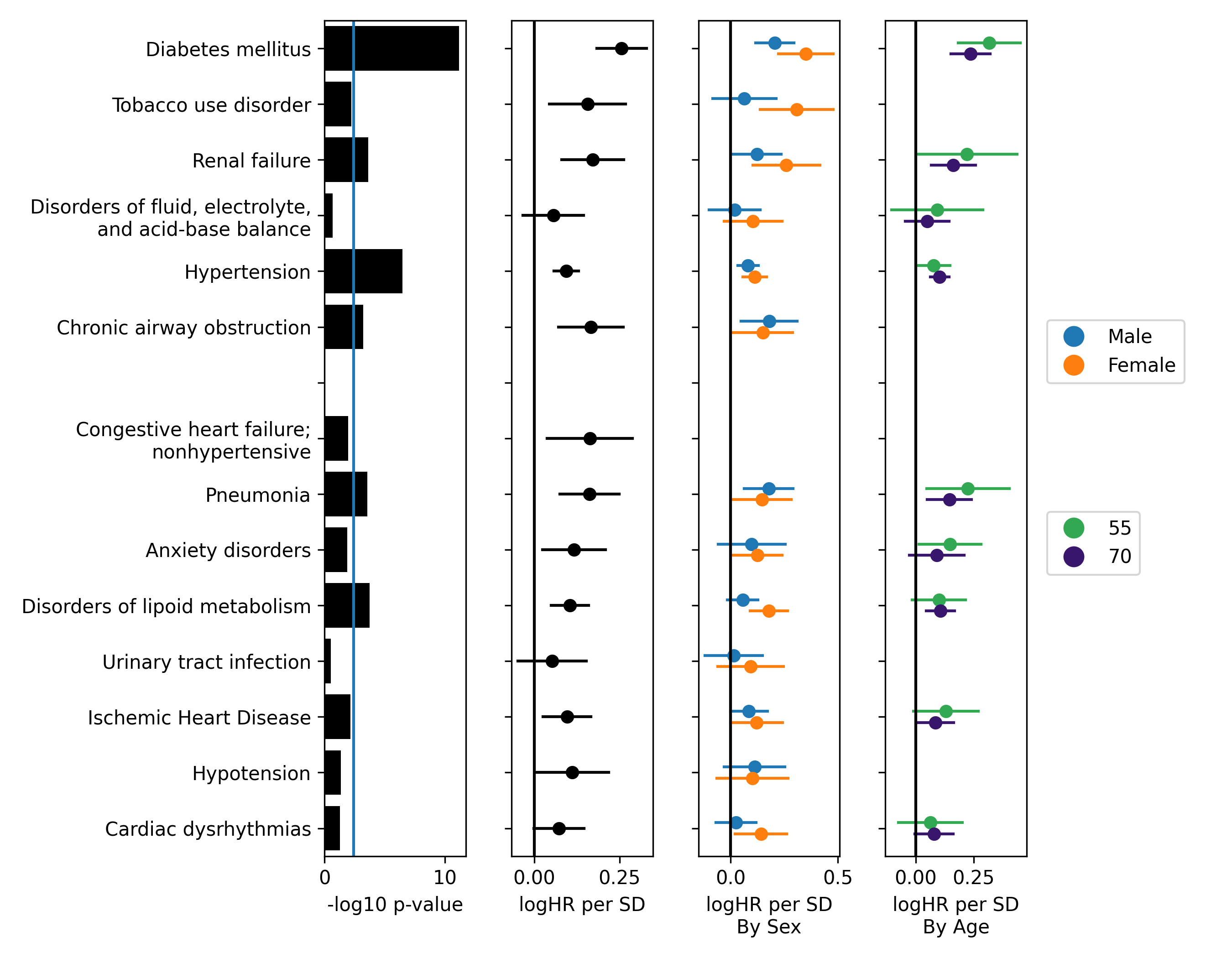

### FIG6.age_effects.png

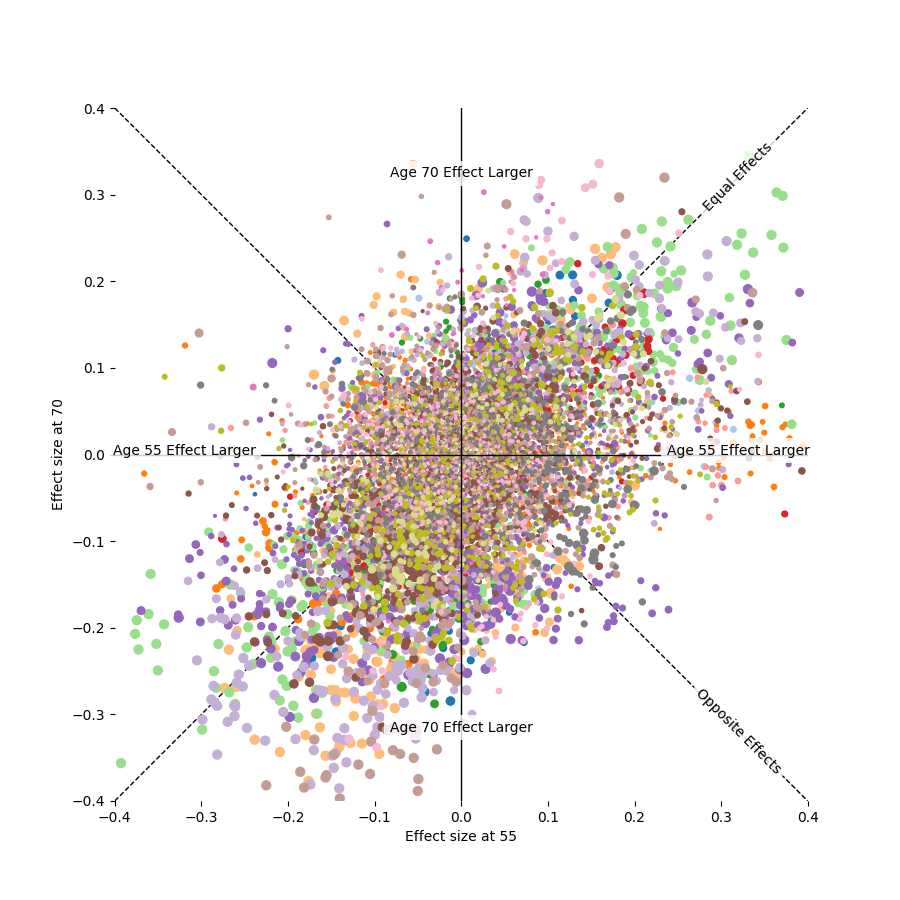

### FIG6.age_effects.quantitative.png

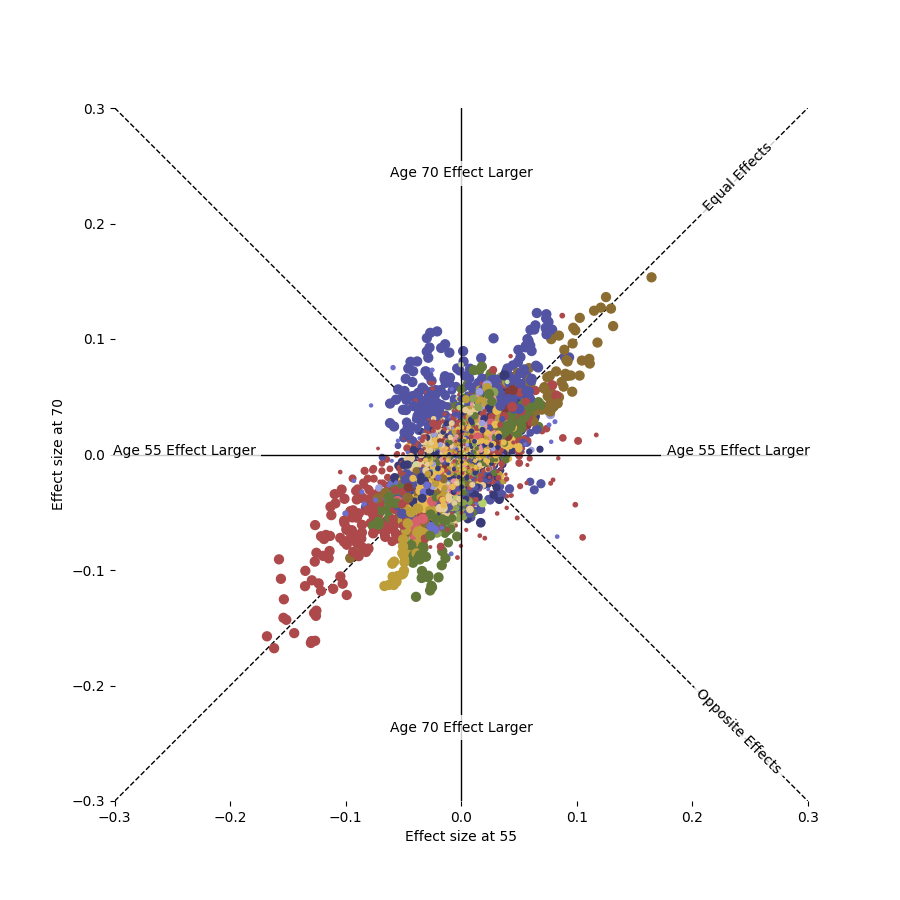

### FIG6.sex_differences.all_phenotypes.png

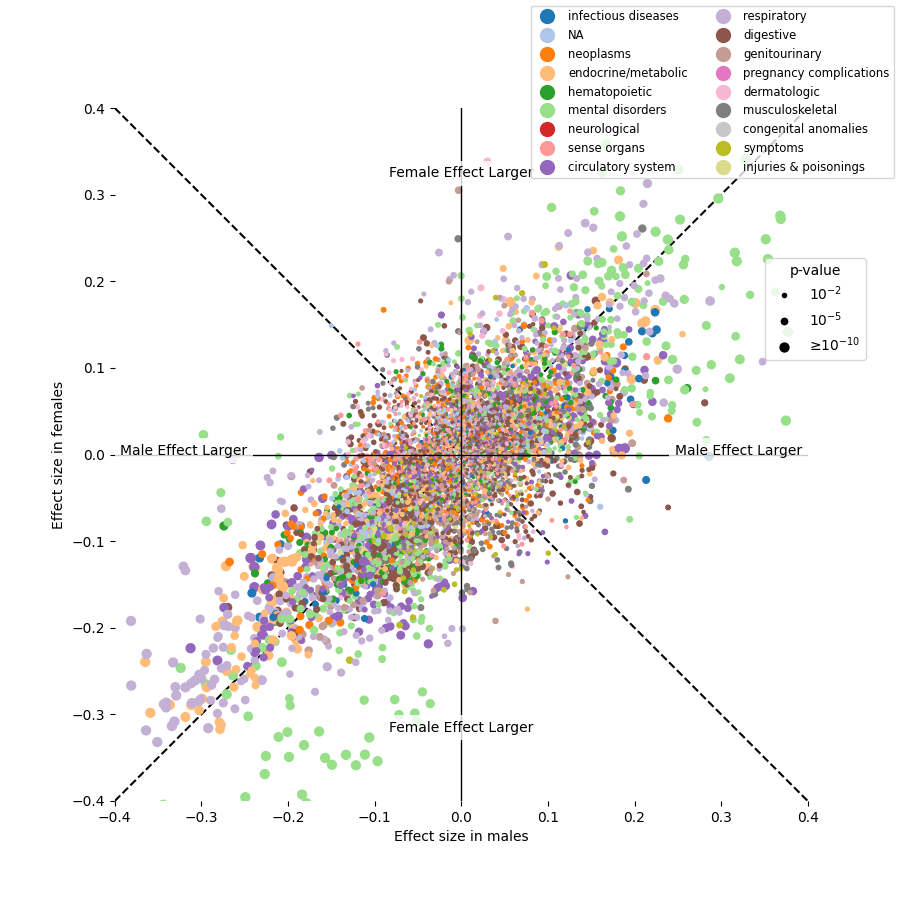

### FIG6.sex_differences.quantitative.png

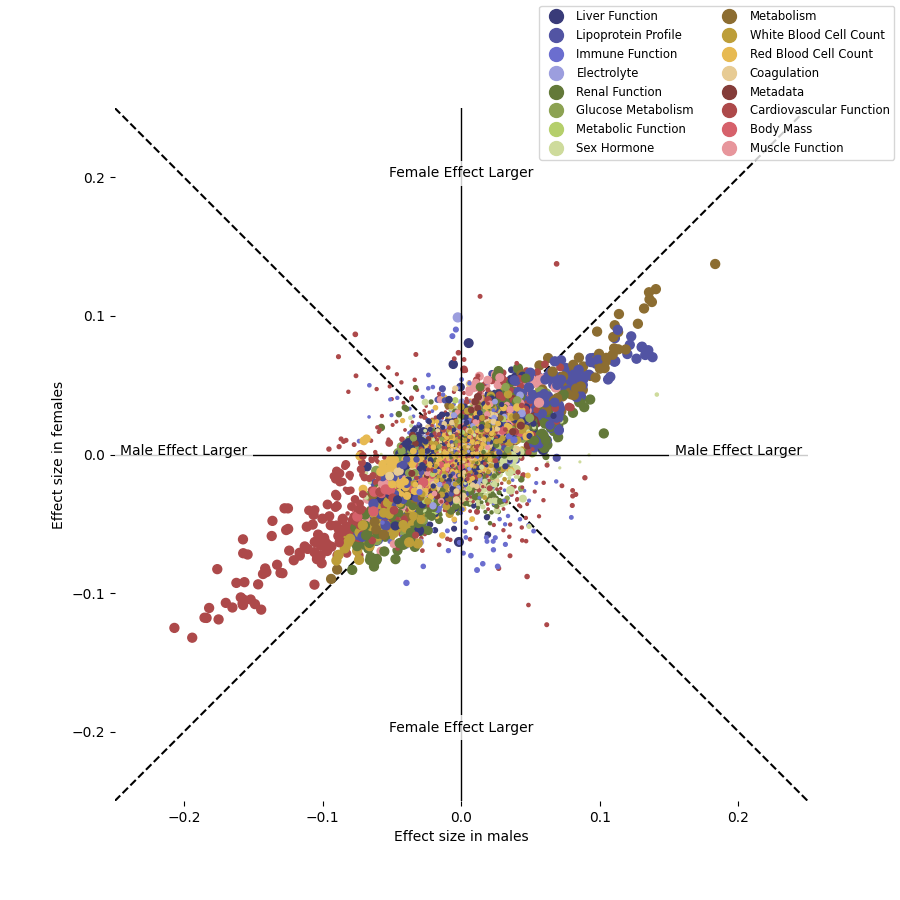

### manhattan_plot.by_activity_var.png

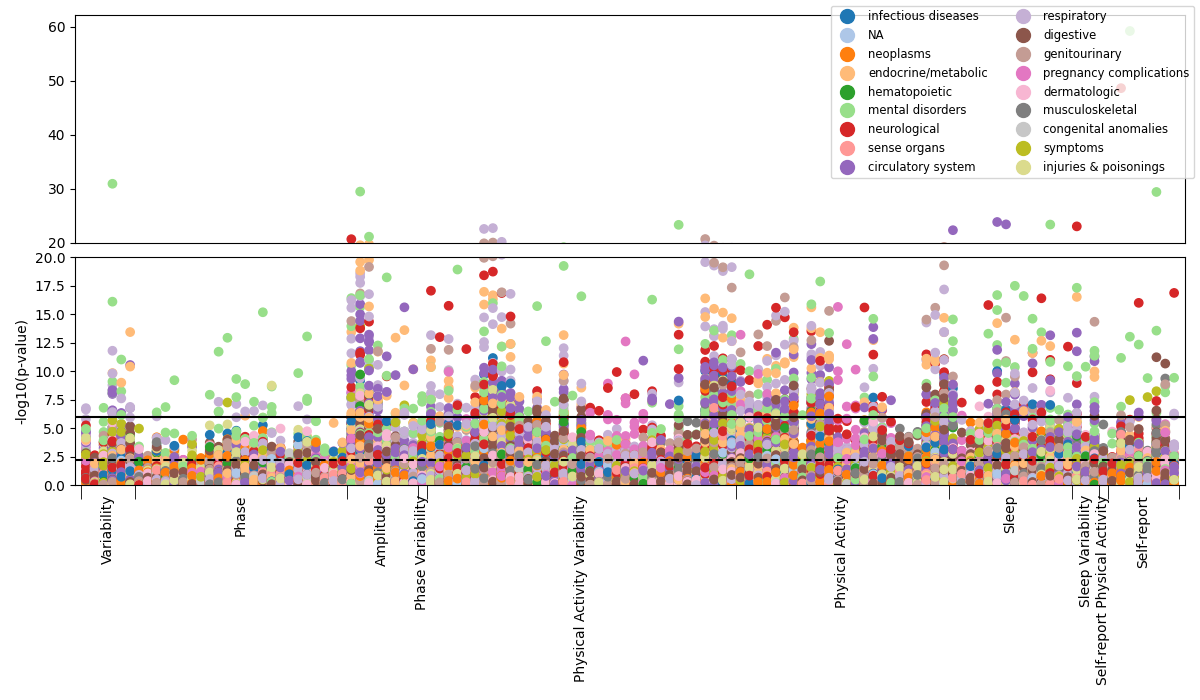

### manhattan_plot.quantitative.by_activity_var.png

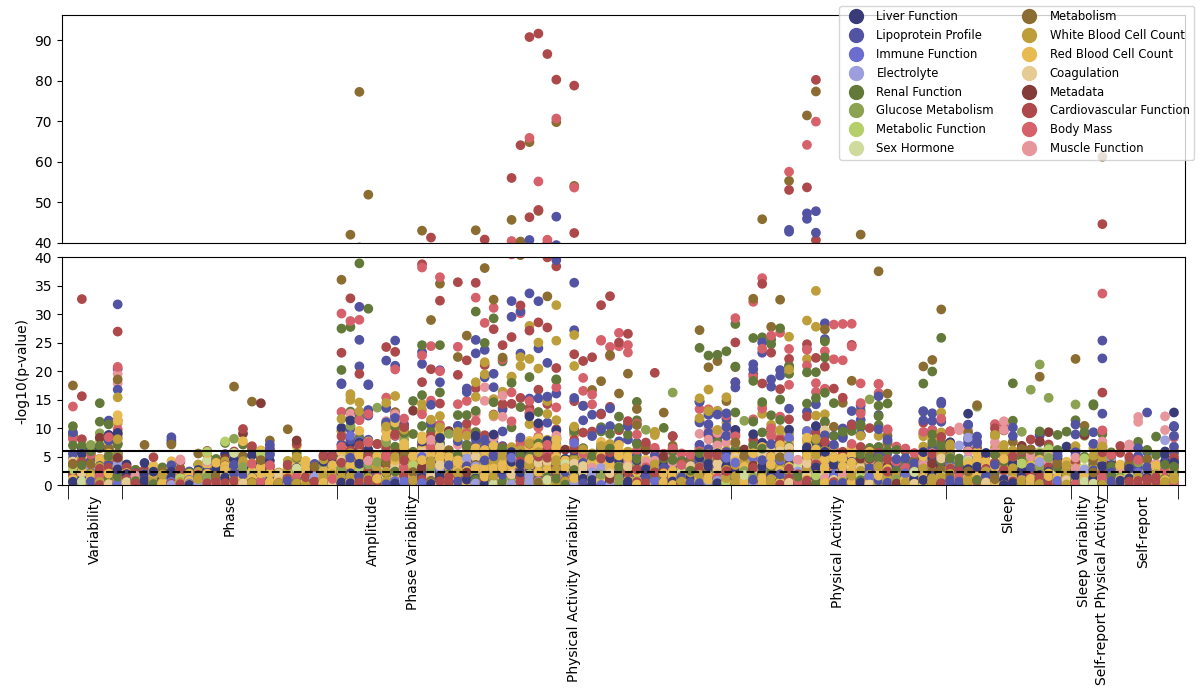

### sex_differences.2x2.png

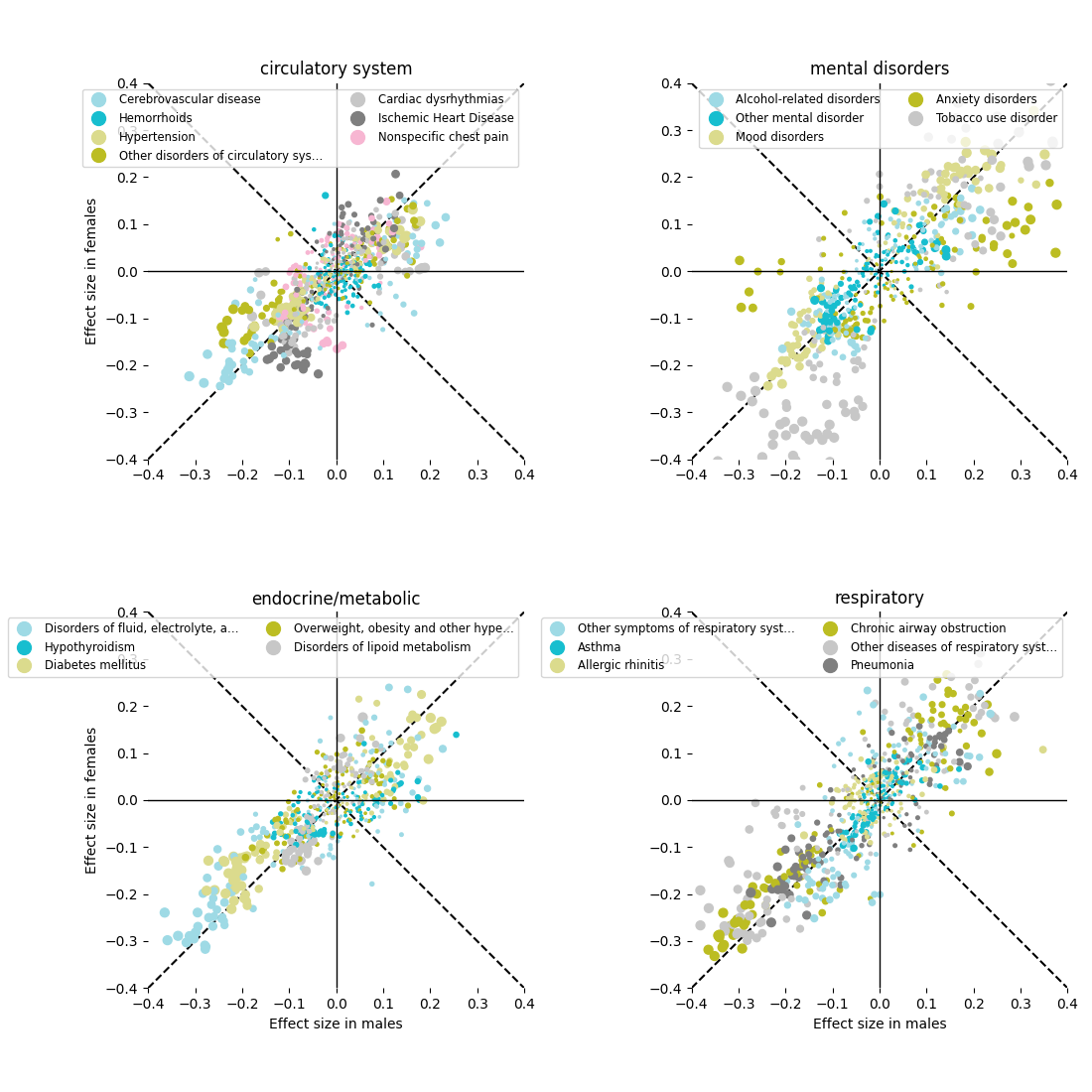

### sex_differences.quantitative.by_activity_var.png

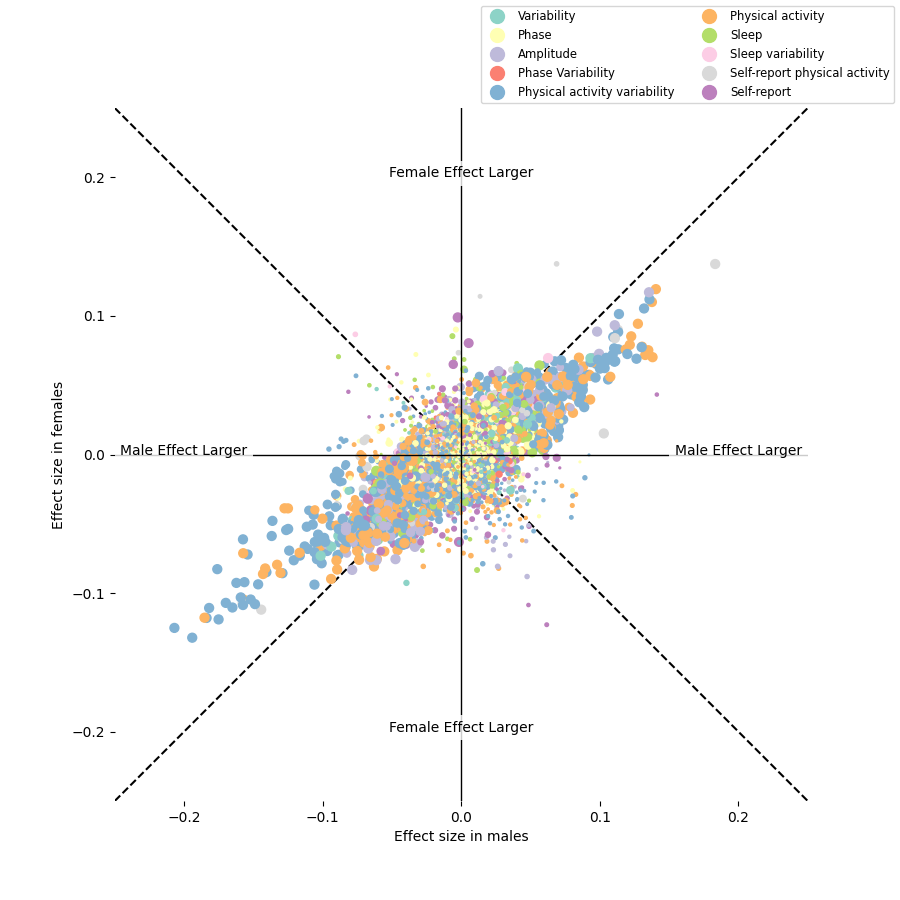

### sex_differences.with_meds.quantitative.lipoproteins.png

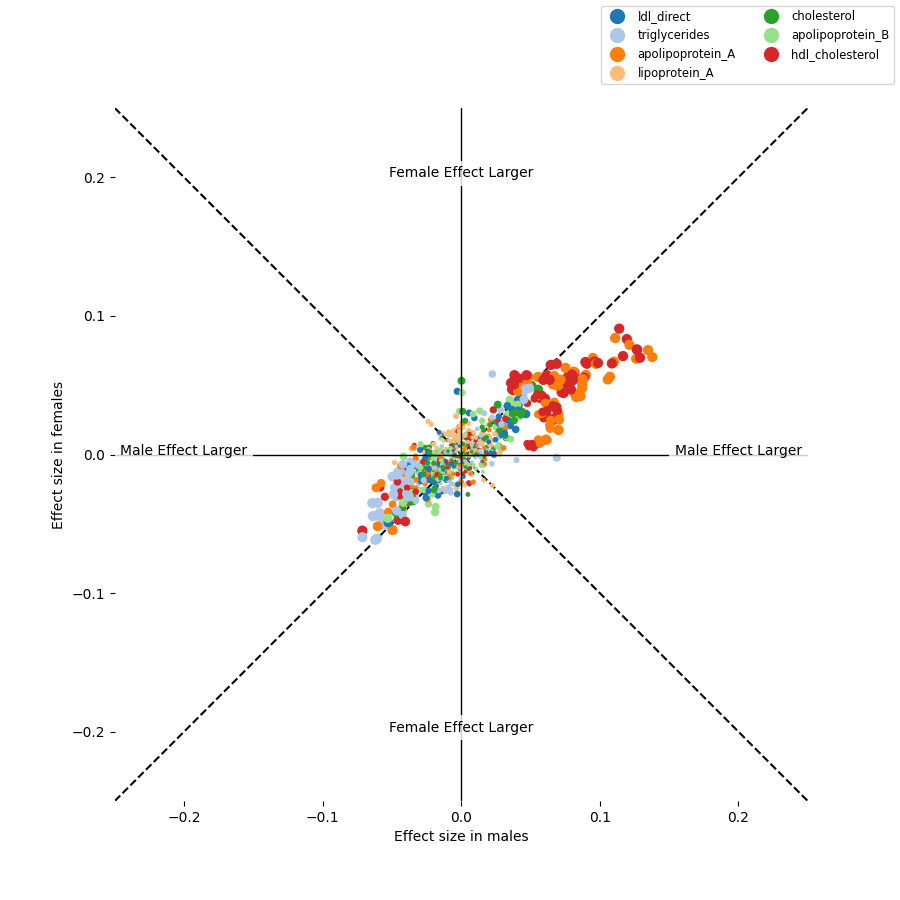

### sex_differences.with_meds.quantitative.png

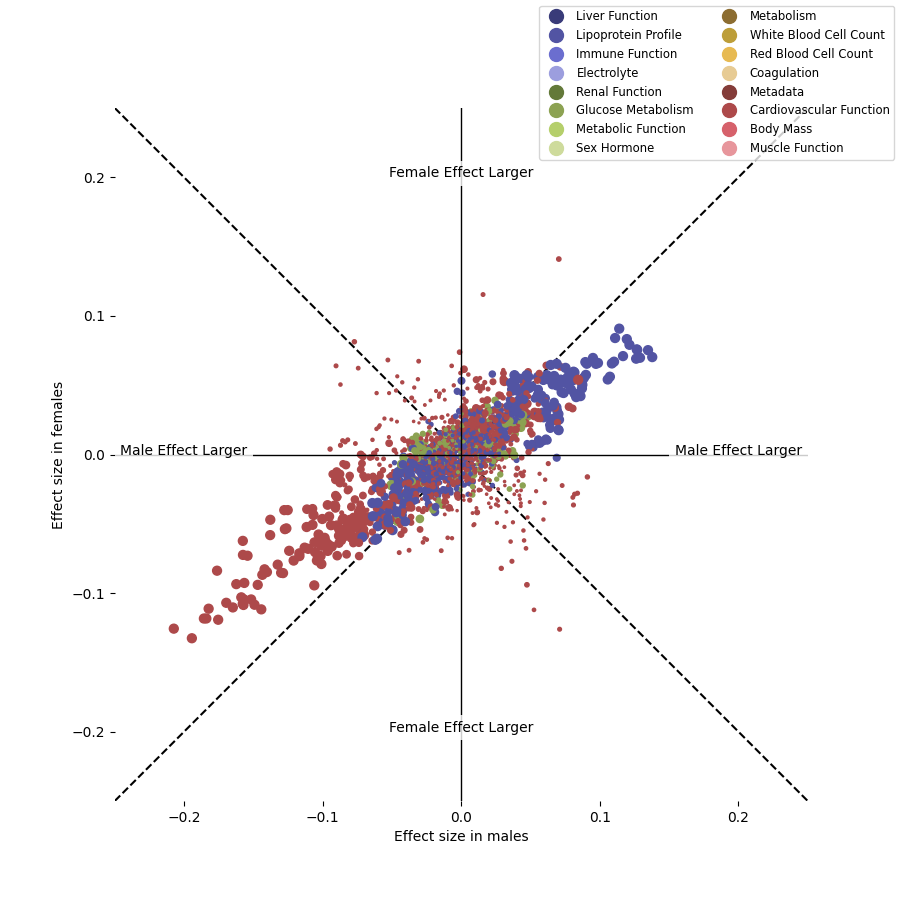

### summary_disease_burden.png

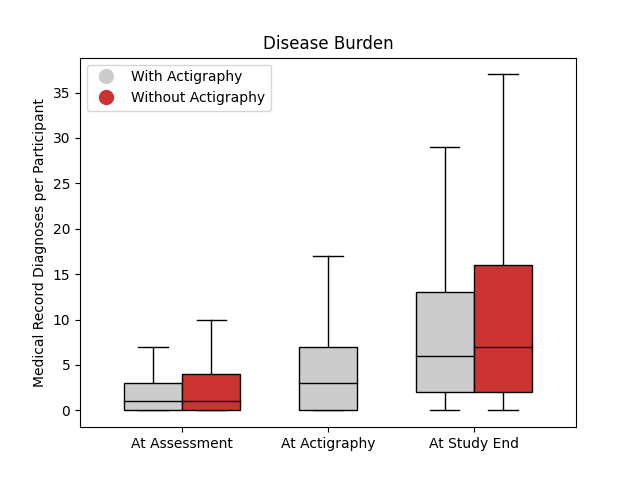

### summary_timeline.png

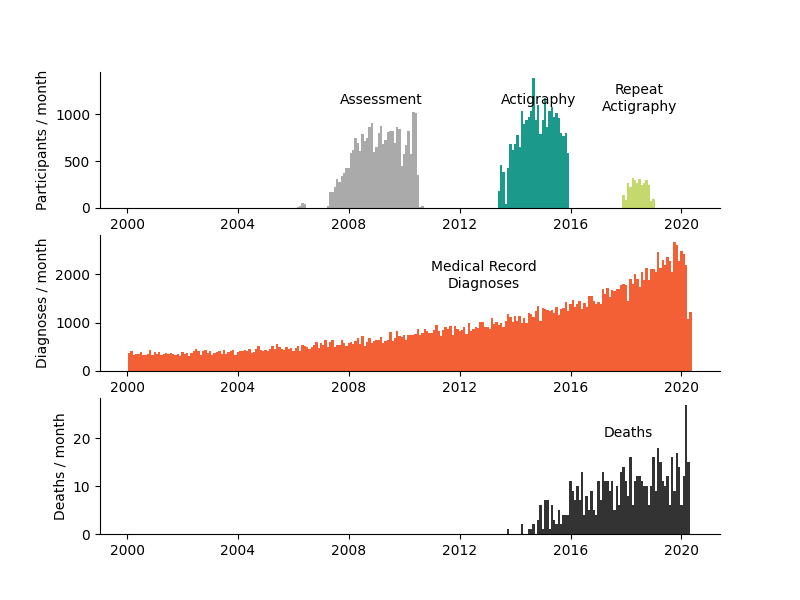

### survival.acceleration_hourly_SD.by_sex.png

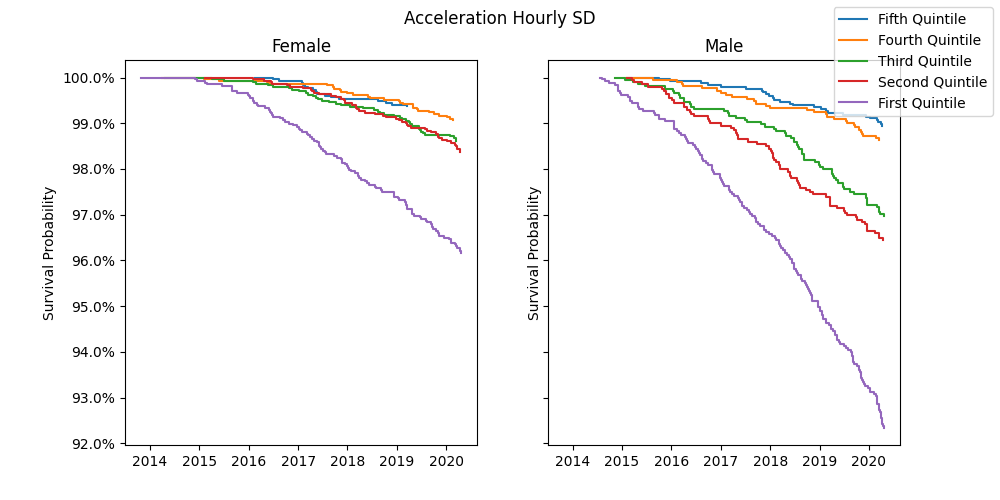

### survival.by_sex.png

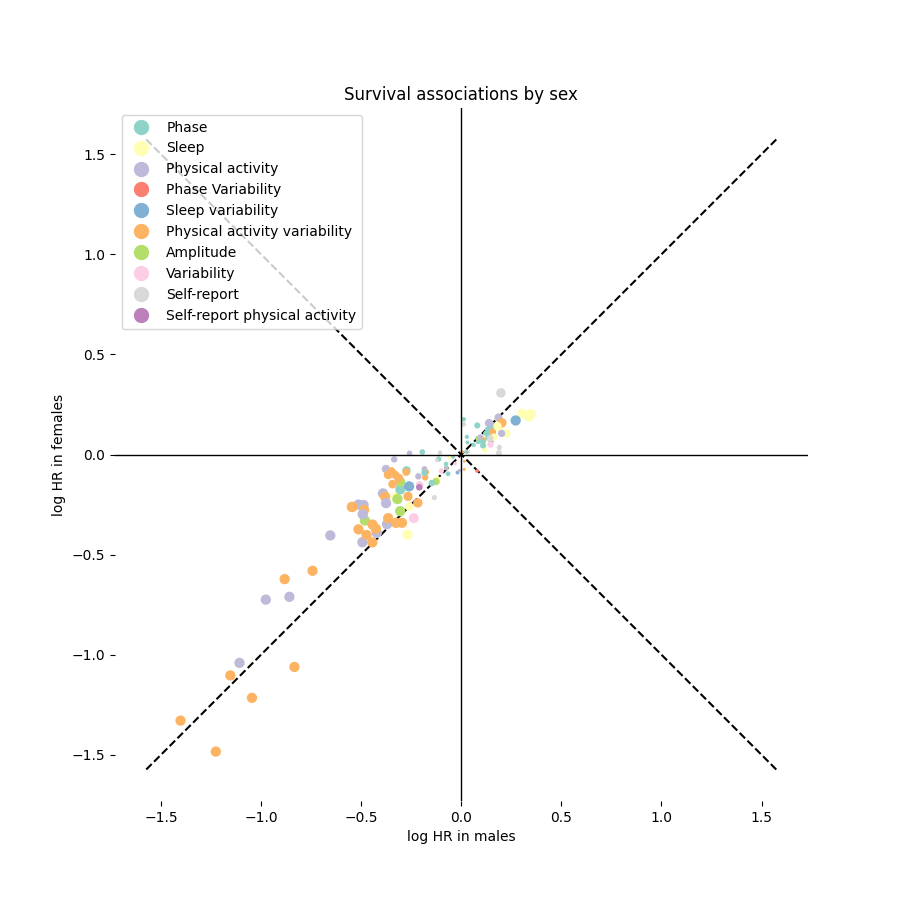

### survival.main_sleep_ratio.by_sex.png

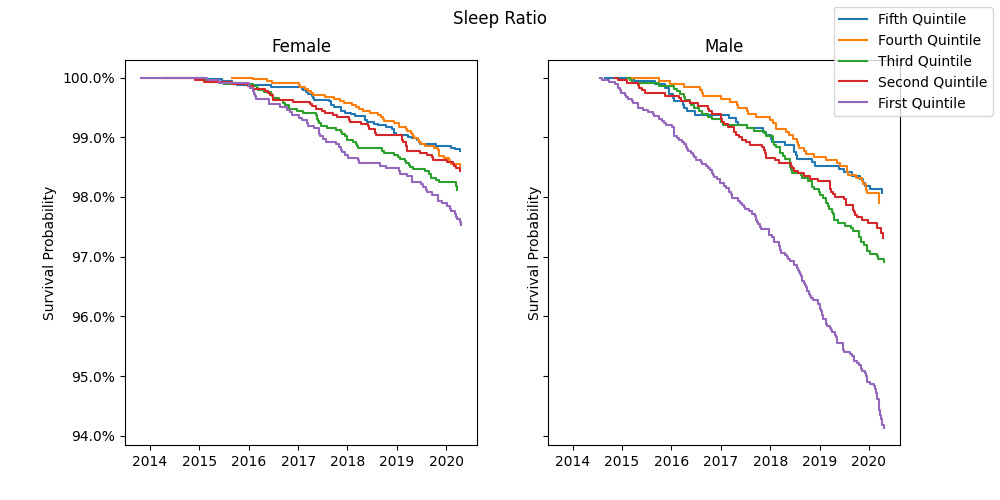

### survival.moderate_between_day_SD.by_sex.png

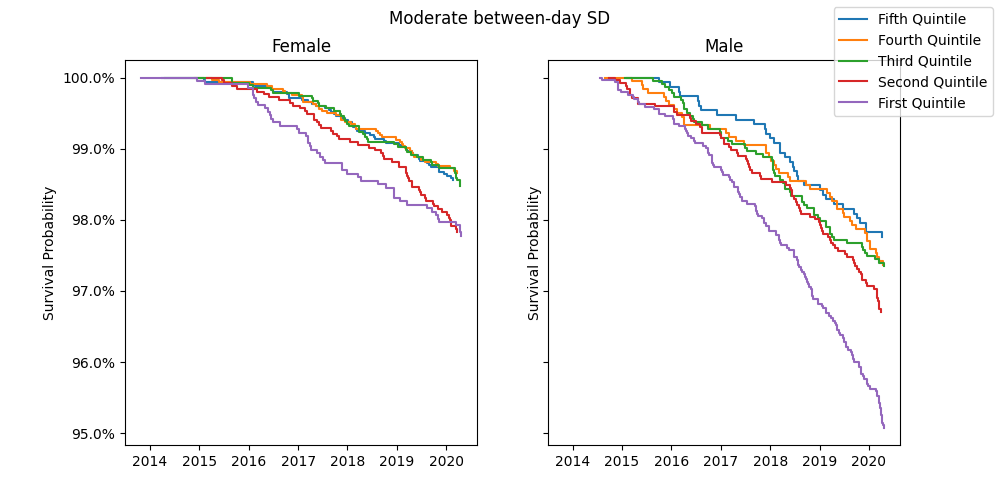

### survival.RA.by_sex.png

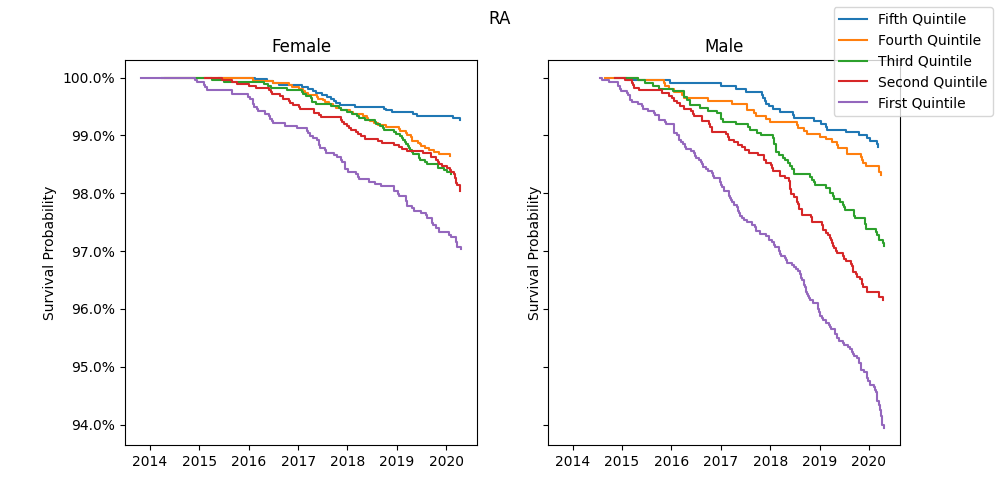
