## Supplemental Text for "Phenome-Wide Association Study of Actigraphy in the UK Biobank"

**Supplemental Results**

Exploratory Cohort

Findings were consistent across smaller exploration and larger validation cohorts. Supplementary File 1 contains corresponding analyses for the exploration cohort.

Ambient Light Level Sensors

In addition to accelerometer- and temperature-derived measures, measures were also generated from ambient light level sensors included on the actigraphy device. However, all light-level derived measures failed to meet the stability condition under repeat measurements and were excluded.

PheWAS-SensR

Mental and neurological disorders have prominent effect sizes between cases and controls. Patients suffering from schizophrenia and other psychotic disorders (n=97) slept 1.034 standard deviations more or less than controls ($q$=4.2x10$^{-22}$). This was judged by the variable “total_sleep_mean_abs_dev” which is the absolute deviation from the population mean of the individual’s average amount of time spent sleeping per day. This is in line with self-reported difficulties in getting out of bed in the morning (self_report_getting_up_in_morning, Cohen’s $d$=1.06, $q$=5x10$^{-09}$) and the widely reported sleep abnormalities in these disease phenotypes [1]. Moreover, these patients experienced high intradaily variability in their sleep (“sleep_IV”) compared to controls (Cohen’s $d$=0.9, $q$=2.1x10$^{-16}$), reflecting diurnal rhythms deconsolidated by daytime napping and nighttime arousals. For physical activity, schizophrenic patients or individuals with other psychotic disorders showed less switching between active and rest periods than controls as described by “MVPA_hourly_SD_M10”, the variance in moderately vigorous physical activity during the 10 most active hours or *M10* (Cohen’s $d$= -0.6, $q$=9.2x10$^{-8}$). Parkinson’s disease patients (n=256) were, compared to controls, less active (MVPA_overall, -0.8 SDs, $q$=2.9x10$^{-33}$), showed less consolidated diurnal rhythms (acceleration_RA, -0.9 SDs, $q$=1.3x10$^{-43}$), and displayed less restorative sleep (sleep efficiency, -0.9 SDs, $q$=3.3x10$^{-46}$; sleep_peak_quality_mean, -0.8 SDs, $q$=2.4x10$^{-39}$). These observations are consistent with the reported disease phenotype [2, 3].

**PheWAS-SensR in Quantitative Traits**

Subsequently, we incorporated quantitative phenotypes available from the UKB assessment through physical measures and blood and urine samples. In these tests, 5,542 out of 10,292 pairs, tested significantly at a level of $q \leq 0.05$ (Figure 1 f, Table S 5). Nearly all participants were assessed for quantitative traits so that sample sizes usually reached ~60,000 (see supplements for exact numbers).

As expected, quantitative traits characterizing cardiovascular function, body mass, metabolism, and lipoproteins were the most significant to associate with physical activity. For example, heart rate was lower among those with higher maximum physical activity (acceleration_peak_value_mean, $r$=-0.14, $q$=5.2x10$^{-241}$) and those people also had smaller waist circumference (acceleration_peak_value_mean, $r$=-0.05, $q$= 1.1x10$^{-177}$), both illustrating the known health effects of exercise [4].

Associated metabolic changes underscore the physiological impact of exercise, where higher levels of physical activity associated with higher levels of vitamin D (acceleration_peak_value_mean, $r$=0.12, $q$=1.5x10$^{-165}$), and apolipoprotein A (acceleration_overall_M10, $r$=0.09, $q$=5.3x10$^{-127}$), the primary protein component of HDL and of HDL cholesterol (acceleration_overall_M10, $r$=0.09, $q$=2.7x10$^{-120}$). Improved organ function was implied in exercised participants. Lower cystatin C, a cellular breakdown product excreted by the kidneys, was associated with more exercise (acceleration_overall_M10, $r$=-0.07, $q$=2.6x10$^{-66}$), as was creatinine measured in urine (MET_overall, $r$=-0.07, $q$=1.6x10$^{-76}$) and in blood (MET_overall, $r$=-0.06, $q$=2.4x10$^{-57}$). Modulation of the immune system was suggested by lower white blood cell and neutrophil counts in those with overall higher levels of exercise (acceleration_overall, $r$=-0.06, $q$=6.7x10$^{-45}$ and $r$=-0.06, q=2.2x10$^{-46}$, respectively).

The relationships described above were also found in the diurnal variability of quantitative traits (1,524 out of 5,542 pairs at a level of $q \leq 0.05$). Higher heart rate (acceleration_RA, $r$=-0.08, $q$=1.4x$10^{-82}$) and higher levels of vitamin D (acceleration_RA, $r$=-0.09, $q$=4x$10^{-97}$), for example, were associated with a high relative amplitude of movements. These directional associations could similarly be observed for sleep (988 out of 5,542 pairs at a level of $q \leq 0.05$)

Here, our findings underscore the congruence between associations for actigraphy-derived metrics of physical activity, sleep, and diurnal rhythmicity with disease phenotypes and quantitative traits.

Survival

Physical activity quantified by acceleration_overall associates substantially to survival (log HR=-0.69 per SD, q=2.8x$10^{-95}$). This variable calculates the mean acceleration vector magnitude from the actigraphy device's accelerometers. Similar strong survival associations held for related variables such as acceleration_within_day_SD (the mean across days of the standard deviation of the hourly binned acceleration vector magnitudes) and acceleration_hourly_SD (the mean across days of standard deviation across hourly binned acceleration vector magnitudes). Circadianness represented by acceleration_RA similarly showed a strong association with survival (log HR=-0.028 per SD, $q$=1x$10^{-61}$) and without sex differences ($q$ = 0.69).

For sleep, the top association to survival was with sleep efficiency (q = 1.8x$10^{-88}$, log HR -0.37 per SD), which aims to measure the average fraction of in-bed time spent asleep. Variables reflective of fragmented sleep such as num_wakings_mean and WASO_mean associate strongly with all-cause mortality ($q < 10^{-77}$, log HR ≥ 0.35 per SD) without affecting sexes differently (sex_difference_q >0.6).

Survival associations were consistent across sexes, **Figure S 4**.

These associations are visualized in **Figure S 9** where mortality tracks dose-dependently for each quintile of participants for each domain of physical activity, sleep and circadianness. Comparing the extremes, the first versus the fifth quintile, the survival trajectory unfolds substantially different. For circadianness (defined by acceleration_RA), the difference in mortality after 5 years amounts to 156 cases (1$^{st}$ quintile) compared to 560 cases (5$^{th}$ quintile), a 3.6-fold difference. For physical activity (acceleration_hourly_SD), this difference is 4.2-fold (146 cases in the 1$^{st}$ quintile compared to 608 cases in the 5$^{th}$ quintile).

Other sleep associations include total_sleep_mean_abs_dev ($q < 10^{-9}$, log HR 0.25 per SD), which means the difference of total sleep time from the population mean and was more significant than total_sleep_mean ($q < 10^{-6}$, log HR = 0.20 per SD) indicating that likely both low and high sleep durations are associated with all-cause mortality. Among self-reported sleep metrics, daytime napping ($q < 10^{-7}$, log HR = 0.91 “Usually” versus “Never” or “Rarely”) showed the strongest associations with all others (including sleep duration, sleeplessness, and difficulty getting up in the morning) being substantially weaker associations. Meta-analyses to date have found that self-reported sleep duration surveys do not have strong evidence of association with survival [5], concordant with our findings that objective sleep duration measures strongly out-perform self-reported duration. Similarly, subjective sleep quality measures may not associate with all-cause mortality in Europeans [6], but we find the objective sleep quality measures perform well. Moreover, we replicate [7] which found that rhythmicity and WASO were the most important of seven sleep-related measure for predicting mortality.

**Sex-behavior Interaction in Diagnoses**

While activity-phecode relations were largely stable between sexes, we highlight certain differences, see **Figure S 5 top row**. The abdominal hernia phecode had significant sexual dimorphism in its associations with 16 actigraphy health variables related to physical activity, for example, walking_overall (sex interaction $q = 1.4 \times 10^{-5}$), and its variability (walking_within_day_SD, $q = 1.4 \times 10^{-4}$). The risk for hernias is skewed towards men [8, 9], however our comparison of cases-to-controls means that differences in case counts between sexes should not bias the results towards significance. We observed that male abdominal hernia cases had larger walking overall time than male controls (*n*=5,928, Cohen's *d*=0.05 in males), while female cases have less walking overall time than female controls (*n*=4,092, Cohen's *d*=-0.09 in females). Compared to male cases, the higher risk to relapse in female cases [10] and to experience more pain postoperatively [11], may have had an inhibitory effect on establishing higher levels of physical activity among female cases compare to female controls. This raises the question of whether improved postoperative pain management in women allows the achievement of a higher degree of physical activity which is then protective against relapse.

Sleep disturbances are commonly associated with mental illnesses [12] and in this context are more frequently reported by women [13]. This is evident in the case numbers of the present cohort, with about double the number of women (*n*=2,344) indicating daytime dozing compared to men (*n*=1,130). The measure "other_sleep_mean", a variable to quantify average daily napping duration, supports this observation (Cohen's *d*=0.26 in males versus 0.11 in females, sex interaction *q* =0.003), and this agrees with self-reported higher daytime dozing sex interaction (*q*=0.0003). This is provocative as evidence to date indicates women experiencing a more pronounced interaction between disturbed sleep and mood swings [14].

#### Age-behavior Interaction in Diagnoses

Age-dependent associations between behavior and diagnoses were more common than sex-specific differences (1397 age interactions versus 54 sex interactions were significant at $q < 0.05$), see **Figure S 5 third row**. Hypertension emerged as one of the top PheCODEs to show substantial interactions between behavioral health measures and age (51 associations significant at $q \leq 0.05$, Table S 4). For walking during the most active time (walking_overall_M10) there was a strong age-dependent lower level of physical activity in hypertensive cases compared to normotensive controls (*n*=20,720, age interaction $\Delta d$=-0.01/year, $q=5.2\text{x}10^{-17}$).

To illustrate these interactions, we calculated effect sizes and significances for 55- and 70-year-old cases and controls. Here, hypertensive septuagenarians walked much less during their most active time than their age-matched controls (walking_overall_M10, age_70_std_effect=-0.14, $q$= 2.2x$10^{-35}$), while hypertensive 55-year old's did not differ from their age matched controls (walking_overall_M10, age_55_std_effect= 0.007, $q$=0.84). This corresponds to 70-year-old hypertensives walking 6.8 minutes less per day, on average, during their M10 than their normotensive counterparts. For 55-year-old cases versus controls, the difference in walking time was negligible, only 0.29 minutes per day, on average. However, this difference is significant ($q$=1.9x$10^{-13}$ with the null hypothesis that M10 walking difference is non-zero). What appears to be a little extra effort per day, amounts to roughly 2.2-3.7 million additional steps over the course of 15 years when converting minutes to steps [15].

A diagnosis of hypertension often tracks with sleep disturbances [16]. Time asleep during the main sleep period (sleep efficiency) decreased age-dependently and degree of sleep disruption was much more pronounced in 70-year-old hypertensives compared to their controls than for the 55-year-old case-control comparison. Circadian rhythms were also implicated. Amplitude and $r$-squared of cosinor fits decline more rapidly in aging hypertensives compared to their controls ($\Delta d$=-0.001/year, $q$=1.8x$10^{-17}$ and $\Delta d$=-0.001/year, $q$=3.8x$10^{-15}$, respectively).

In contrast to hypertension, mood disorders showed only an age interaction for a few sleep variables with age. There was a narrowing difference between cases with mood disorder and controls over age in sleep duration (main_sleep_duration_mean, std_age_effect=-0.012/year, $q$=5x$10^{-05}$). Here, the significant interaction at age 55 in the case-control comparison (age_55_std_effect=0.176, $q$=5.5x$10^{-12}$) has disappeared at age 70 year where cases with mood disorder sleep similar amounts as their controls (age_70_std_effect=-0.001, q=0.98). Similar effects were also seen with the self-reported difficulty getting up. For anxiety patients, results were similar for self-reported difficulty getting up but had less of an aging effect for quantitative sleep duration (main_sleep_duration_mean, Δd=-0.012/year, $q$=0.0004).

Two conditions, ageing and mood disorders, known to interfere with diurnal rhythms, surprisingly do not produce a significant interaction in the UKB population (acceleration_RA, Δd=-0.005/year, $q$=0.21; amplitude, Δd=-0.003/year, $q$=0.535; phase, Δd =-0.0007/year, $q$=0.927).

In other diseases, diabetic patients also demonstrated age dependence on behavioral differences in sleep and activity levels. Chronic airway obstruction patients showed age-dependent interaction primarily with measures of physical activity ($n$=7), its variability ($n$=8) as well as diurnal rhythmicity ($n$=2).

#### Age-behavior Interaction in Quantitative Phenotypes

Age interactions with the phewas were investigated in quantitative phenotypes, **Figure S 5** second row and Table S 5. The known age-related decline in renal clearance of both creatinine and cystatin c [17] was significantly blunted in individuals with higher levels of physical activity (acceleration_overall, $n$=63,364, std_age_effect=-0.004/year, $q=8.1\times10^{-22}$ and $n$=63,380, std_age_effect=-0.005/year, $q=4.4\times10^{-32}$, respectively) as well as higher degrees of variance in their exercise behavior. Here, cystatin c, unlike creatinine, allows assessment of renal function independent of skeletal muscle mass. The trajectories of these relationships suggest that physical activity-related dampening of the age-dependent retention of these metabolites became more pronounced with age (age_55_std_effect=-0.01, $q$=0.06 compared to age_70_std_effect=-0.07, $q=7.1\times10^{-44}$ for creatinine and age_55_std_effect=-0.03, $q=5.1\times10^{-9}$ compared to age_70_std_effect=-0.11, $q=5.9\times10^{-88}$ for cystatin c). In contrast to studies with small sample sizes [18], our results suggest that cystatin c levels are modifiable by physical activity over the entire age spectrum of 45-80 years which are represented by meaningful sample sizes in the UKB (Figure S 8). Exercise may confer a protective effect on kidney function under pathophysiological duress as postulated, for example, for patients with acute myocardial infarction [19]. Urinary creatinine excretion declined as expected [20] over age, dependent on the level of physical activity but without a meaningful interaction (acceleration overall, std_age_effect=-0.0004/year, age interaction $q$=0.6).

Distinctive directional associations surface for analytes of the lipoprotein metabolism. HDL cholesterol blood levels follow stringent exercise-dependent patterns (Figure S 8), where the slopes of the quintile curves suggest an interaction with age (acceleration_overall, n=58,060, std_age_effect=0.003/year, $q$=6.1x10$^{-10}$), meaning that, for example, the 0-20% quintile in physical activity did not experience an exercise-induced increase in HDL-C past the age of 60 years. Triglyceride levels increased age-dependently and were modified by physical activity (acceleration_overall, $n$=63,348, age_55_std_effect=-0.05, $q$=9.7x10$^{-26}$ versus age_70_std_effect=-0.04, $q$=8.9x10$^{-9}$) but without significant interaction (std_age_effect=0.001/year, $q$=0.04). Age- and exercise-specific interactions are, as expected, absent in levels of lipoprotein A (Figure S 8), which are largely genetically determined [21].

Both cholesterol and LDL-cholesterol increased age-dependently between 45 and 65 years of age. High levels of physical activity associated on average with 0.2 mmol/L lower LDL cholesterol levels than lack of exercise. These relationships change dramatically, however, for individuals aged 65 to 80 years. Consequently, this is flagged as a highly significant interaction (cholesterol and acceleration_overall, $n$=63,382, std_age_effect=0.001/year, $q$=7.5x10$^{-67}$) as shown in Figure S 8.

### Most sex- and age-specific associations are stable despite prescription medicine intake

Since medication use has large, direct effects on certain measured quantitative phenotypes, we controlled for the reported use of medications affecting cholesterol, insulin, or blood pressure among phenotypes classified as lipoprotein profile, glucose metabolism, or cardiovascular function, respectively. Medication use was reported at assessment time, which coincided in time with the measured quantitative phenotypes. Compared to the model without controlling for medication use, sex-specific and age-specific effects largely replicated among highly significant associations in medicated individuals as shown in Figure S 10. However, the age-specific dispersion in measures of the lipoprotein profile was decreased in the cohort medicated with cholesterol-lowering prescription drugs (Figure S 10 center left) compared to the unmedicated reference cohort (Figure S 10 center right), suggesting that the interaction between age and lipoproteins is altered by medication.

However, significant age interactions were found for LDL-C between medicated and unmedicated. To address this, we examined exercise relations with LDL-C separated by age, sex, and use of cholesterol-lowering prescription medicine (Figure S 11). We note that LDL-C in males peaks in 50–55-year-old individuals followed by a steady decline in LDL-C levels at older age. In women, however, LDL-C steadily rises until 60-65 years, then plateaus at older age. For both sexes, these relationships were not affected by physical activity by either the quantitative (acceleration_overall) or qualitative (IPAQ score) measures (Figure S 11). Similar sex-specific patterns emerged for unmedicated individuals, but here higher levels of physical activity over age were associated with lower LDL-C levels. This relationship was reproducible across objective and subjective measures of physical activity and more pronounced in males (Figure S 11 center). Lastly, in medicated males LDL-C levels steadily decline over age without additional benefit from elevated physical activity, possibly linked to statin-induced impairment of exercise capacity [22] albeit this is debated [23]. In medicated females, LDL-C levels remain steady across age groups, unaffected by different levels of physical activity (Figure S 11 right). Therefore, the observed age interactions of LDL-C and physical exercise was driven by cholesterol-lowering medication use, since observed effect sizes are consistent across the age range, in both medicated and unmedicated men and women.

These data underscore the differential approach for cardiovascular risk assessment by sex and age as recently proposed by Agarwala and colleagues [24]. Specifically, our observation that statin-naive women do not benefit from increased physical exercise (i.e. lower LDL-C levels) might suggest that this lifestyle modification is less effective than in men.

**Supplemental Methods**

Activity Variables

The UK Biobank project collected 1 week of actigraphy using custom accelerometer devices on 103,688 participants using the Aximetry AX3 wrist-worn device on dominant wrists [25]. The data were calibrated and processed using an existing pipeline [26] modified slightly to better accommodate varying time zones and to output light and temperature readings (https://github.com/tgbrooks/biobankAccelerometerAnalysis). In short, this pipeline performs calibrations of the accelerometer readings per recording, then runs a balanced random-forests classifier trained from almost 160,000 minutes of labelled data on 132 individuals. The classifier labels each 30 second epoch as one of six activity types (sleep, sedentary, walking, light-tasks, moderate activity, or non-wear) and computes a Metabolic Equivalent of Task (MET) score indicating relative energy expenditure. It then runs a hidden Markov model was then run on the resulting activity type labels to perform smoothing.

After the classification pipeline completed, we then performed further analysis of the sleep time in order to extract sleep measures. Sleep was divided into up to one “main sleep” period per day, to approximate the typical sleeping pattern of one overnight sleep. All sleep outside the main period was then reclassified as “other sleep,” to be interpreted as napping. To determine the main sleep period, sleep periods were joined with adjacent sleep periods if several conditions were met. First, the total amount of non-sleep time between all joined periods was less than 2 hours. Second, the later sleep period is at least twice as long as the gap between it and the previous sleep period. Thirdly, the earlier sleep period is at least 2/3rds of the length of the gap between it and the next sleep period. The main sleep of a day is then taken to be the longest of all joined sleep regions that overlap the given day. These rules were determined by manual inspection of example subjects and were tailored to the results of the actigraphy classification pipeline.

From the main sleep period, several descriptive statistics were generated: main sleep onset (i.e. start time), main sleep offset (i.e. stop time), main sleep duration, wake after sleep onset (WASO) [27, 28], main sleep ratio [29] (duration – WASO / duration), number of awakenings during main sleep [27], and total acceleration during the main sleep period. For each classified activity type, daily summary statistics were computed for peak values and times, determined by a sliding Gaussian-weighted average with standard deviation 1 hour. For temperature and light, the mean, 10th and 90th percentiles were computed per day, to capture both typical and extreme values.

All daily values were then summarized at the participant level by taking the average over all days and the standard deviation across days.

To examine within-person variability specifically, we constructed additional variables. For each identified actigraphy health variable (acceleration, light, temperature, and the classified actigraphy types), the values were binned by hour and the mean was taken. Then the variables hourly_SD (standard deviation of these binned values), within_day_SD (mean across days of the standard deviation of the bins within a day), and between_day_SD (standard deviation across the days' mean of bins). This was repeated twice, restricting first to hourly bins within the M10 period (ten hours of the day with highest average acceleration levels) and then restricting to the L5 period (five hours of the day with lowest average acceleration levels). See Figure S 1d. Moreover, the actigraphy health variables were summarized by three rhythmicity measures: the relative amplitude (RA), interdaily variability (IV) and intradaily stability (IS) measures. See [30] for the IV and IS formulas. The RA measure is computed as (M10 – L5) / (M10 + L5) where M10 is the average value of the variable during the M10 period and L5 is the average during the L5 period. Note that M10 and L5 period are all determined based off the acceleration variable to make them comparable across different variables: e.g., the M10 value of the temperature variable is the average temperature during the ten hours of highest average activity. Finally, the midpoints of the M10 and L5 periods are reported as the M10 time and L5 time.

Cosinor fit values [31] for acceleration were computed to derive phase, amplitude, mesor, and r-squared values.

Since it was expected that extreme phase or sleep duration could be related to poor health, for each phase or sleep duration variable, an absolute deviation variable was derived, giving the absolute difference of the individual's measure from the population mean. Therefore, linear models involving those will capture effects consistent across both extreme low and extreme high values in the population.

Outlier data, identified as those whose value was a least 7 standard deviations from the population mean, were excluded. This excluded 5390 (0.05%) data points.

<u>Stability assessment</u>

Repeat assessments of the 7-day actigraphy were performed on a subset of individuals (n = 3197; n=2339 with five total assessments, 727 with four, 93 with three and 38 with two). To assess the stability and repeatability of the variables, the actigraphy health variables were computed on these measurements. For each actigraphy health variable, we computed the population mean of the variance across repeats, followed by determining the population variance between each participant's mean value. The variability of each actigraphy health measure was assessed as the ratio between these values. Only those with smaller within-person variance than between-person variance (i.e. with ratio less than 1) were included from the study as being too unstable, and 96 out of 206 activity variables passed this criterion.

Since each of the repeated measurements corresponds to a different time of year, we accounted for seasonality of an actigraphy health measure with the cosinor fit as a function of time-of-year. Recomputing the within-person to between-person variance ratio on the seasonally corrected data, most variables' ratio remained unchanged as seasonal effects were much smaller than either within- or between-person variance. However, as expected large differences were seen in temperature and light-level derived variables. Despite this limitation, two additional variables passed the variance cut-off (temp_within_day_SD, temp_RA). We included these two additional variables in the study and corrected all for seasonality.

### Validation and Multiple Hypothesis Testing

Due to the exploratory nature of the study, participants with actigraphy passing quality control were randomly separated into two subgroups, one of 25000 as an exploratory cohort and one of the remaining 67325 participants as a validation cohort. The exploratory cohort was used to determine the scope of the project, write and debug statistical tests, and plan what was to be presented. The larger validation cohort was used to compute all final *p*-values and effect sizes and all results reported in this manuscript correspond to this group unless otherwise noted. All results presented in this paper were consistent between cohorts unless otherwise mentioned. All result tables and figures were also generated in this manuscript were also generated from the exploratory cohort and are provided in Supplemental File 1. The pipeline source code run for both cohorts is available at GitHub https://github.com/tgbrooks/ukbb.

To control for the large number of statistical tests performed, *p*-values have been adjusted where appropriate as Benjamini-Hochberg *q*-values to control the false discovery rate.

### Self-reported Variables

The UK Biobank project includes extensive questionnaires performed at initial assessment time. We collected two sets of questions regarding chronotype (field 1180), sleep and sleep quality (fields 1160, 1170, 1190, 1200, 1210, 1220), and one for physical activity levels (IPAQ activity group, field 22032) derived from recalled activity history. Except for sleep duration, these were binarized by comparing the extreme values (Table S 3), and then were included along with the objectively derived actigraphy health measures for all associations.

Survival Analysis

Actigraphy health variables were assessed for their association with all-cause mortality via a Cox proportional hazards model of all-cause mortality as a function of each actigraphy health variable. The model included covariates for sex, BMI and lifetime smoking history. BMI and smoking status were assessed at initial study assessment. There were 1,516 deaths out of the 67,325 individuals assessed. To assess sex-specific effects, a second model was performed including sex as an interaction effect.

PheWAS

A linear model was performed for each actigraphy health measure and each PheCODE pair, modelling the actigraphy health variable as a function of the binary phenotype variable. Covariates were included for sex (male/female), ethnicity (white/other, since 97% are white as in [32]), self-reported overall health (binarized as Good or Excellent versus lower), high household income (binarized as >52,000 pounds/year versus lower), lifetime smoking history (never/ever), age at the time of actigraphy (continuous), BMI (continuous) and college education (college/no college). These covariates (other than age at actigraphy) were measured at the initial assessment and so predate actigraphy by a median of 5 years.

Two further linear models were computed to investigate interactions. First, a linear model containing sex-interaction terms with the phenotype variable as well as the covariate, to account for differences in males and females was performed for each PheCODE with at least 50 cases in each of males and females. Secondly, a linear model containing an age (at actigraphy) interaction with the phenotype variable was performed for each phenotype with at least 200 cases. In order to interpret the age-dependency, the fit effect sizes at age 50 and age 75 were computed, which corresponds to roughly the $20^{th}$ and $80^{th}$ percentiles of the age distribution of the study population.

Circadian Components

A logistic regression was performed modelling each selected PheCODE as a function of three actigraphy health variables (acceleration_overall, sleep efficiency, temp_RA) while controlling for sex, ethnicity, self-reported overall health, high household income, lifetime smoking history, age at actigraphy, BMI and college education. The significance p-values of the three actigraphy health variables was computed (compared to the model with just the other two variables). In order to compare across phenotypes, the effect size of each variable's contribution was determined as the fitted coefficients of each actigraphy health variable and then standardized by the standard deviation of the actigraphy health variable and the overall prevalence of the phenotype, i.e., by Cohen's *d*. Variables with a significant circadian component were identified as those with $q < 0.05$ significance of temp_RA. This process was then repeated on the quantitative phenotypes with an ordinary least-squares model rather than a logistic model. Effect sizes were standardized by both actigraphy and phenotype standard deviations.

Wrist Temperature

Wrist temperatures were determined via a temperature sensor on the Aximetry AX3 device which records temperature as well as acceleration and light levels. While the temperature sensor was originally included for the purposes of calibration of the accelerometer, it provides a useful measure which has components both of ambient environmental temperature and skin temperature [33]. To convert into degrees Celsius, the values reported by our actigraphy pipeline, we corrected them as $T = (500 X - 2550)/256$, where X is the original value reported. This confirms our values to those used in [33].

Since actigraphy devices were used by multiple individuals, we examined the device-effect to check for calibration problems. Indeed, we identified three "clusters" of device ids with distinctly different values reported, Figure S 7 top. The clusters were separated according to device IDs below 7500, between 7501 and 12500, or above 12501. The temperature-derived values were corrected by a multiplicative scaling of each cluster such that the median of each cluster is equal to the overall median.

After this correction, we assessed calibration quality, by comparing the mean values among all individuals who used the same device to the means of a random permutation of the device ids. We expect measures without calibration problems to have a comparable device-level distribution to that generated by the random permutation of device IDs. The mean temperature shows strong device-specific bias compared, Figure S 7 bottom left, indicating a lack of temperature-sensor calibration. However, the temperature-derived RA and cosinor amplitude of temperature measures are relatively robust to poor calibration and so are suitable to use, Figure S 7 bottom center.

### Effect Sizes

Effect sizes for the PheCODE associations were determined as Cohen's *d*, where the standard deviation was estimated from the sample standard deviation of the entire population rather than the pooled estimate. This slightly increases the standard deviation estimate and decreases the magnitude of effect sizes but makes it so that effect sizes are consistent across different diagnoses (which each have different case/control subsets and so different pooled standard deviation estimates). Reported effect sizes for the quantitative phenotype associations were obtained by standardizing the regression coefficient of the behavioral health variable by the standard deviations of the behavior health variable and the quantitative phenotype, obtaining a value between -1 and 1. For age interactions, effect sizes are reported as a difference per year in the standard effect size (i.e., the age-interaction effect size, written $\Delta d$, is the change per year in the Cohen's *d* giving case-control effect sizes).

### Supplemental Figures

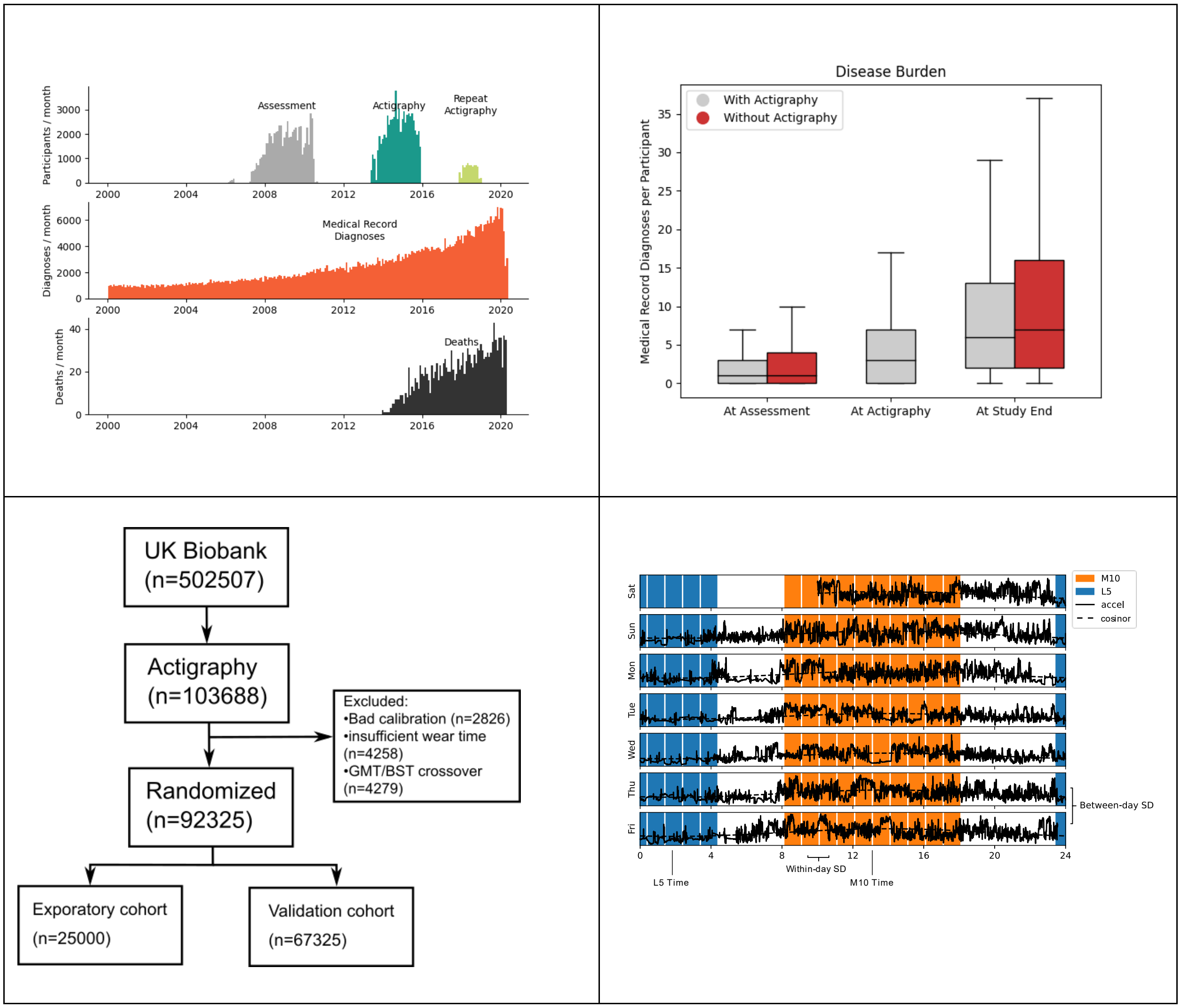


Figure S 1 – UKB data structure

(Top left) Dates of data collection among 67,325 individuals selected to have high-quality actigraphy health measurements. A total of 1,516 deaths and 478,697 distinct diagnoses were recorded. (Top right) ICD-10 medical record diagnosis counts in the selected population at time of assessment, at time of actigraphy, and at the study end compared to the UK Biobank participants without actigraphy. (Bottom left) Study flow diagram. Exploratory cohort was used to perform initial analyses and generate hypotheses. All results presented here are from validation cohort. (Bottom right) Example activity trace from one participant, with M10 period (ten hours of highest average activity) and L5 period (5 hours of lowest average activity) highlighted. Terms for generated actigraphy health variables are highlighted.

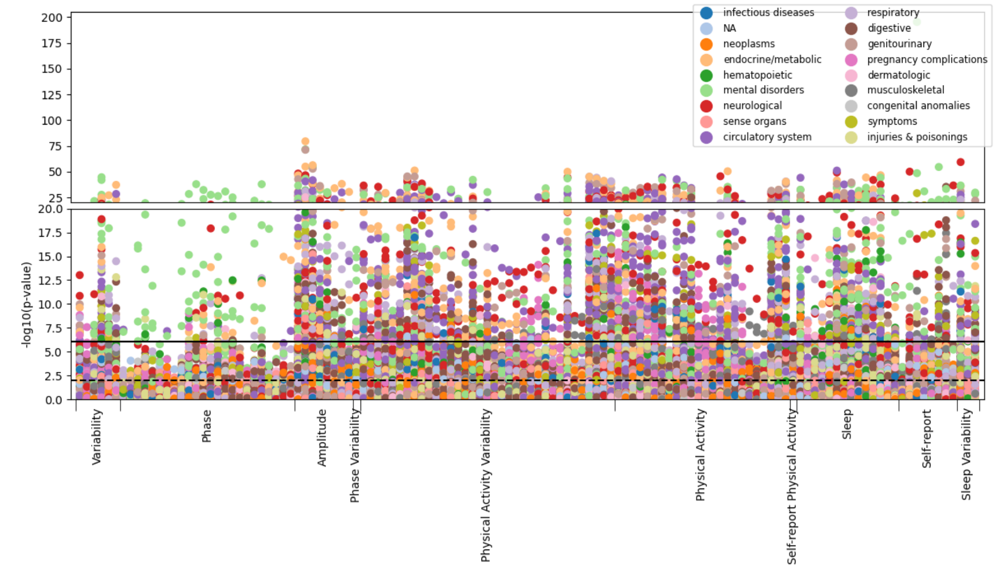


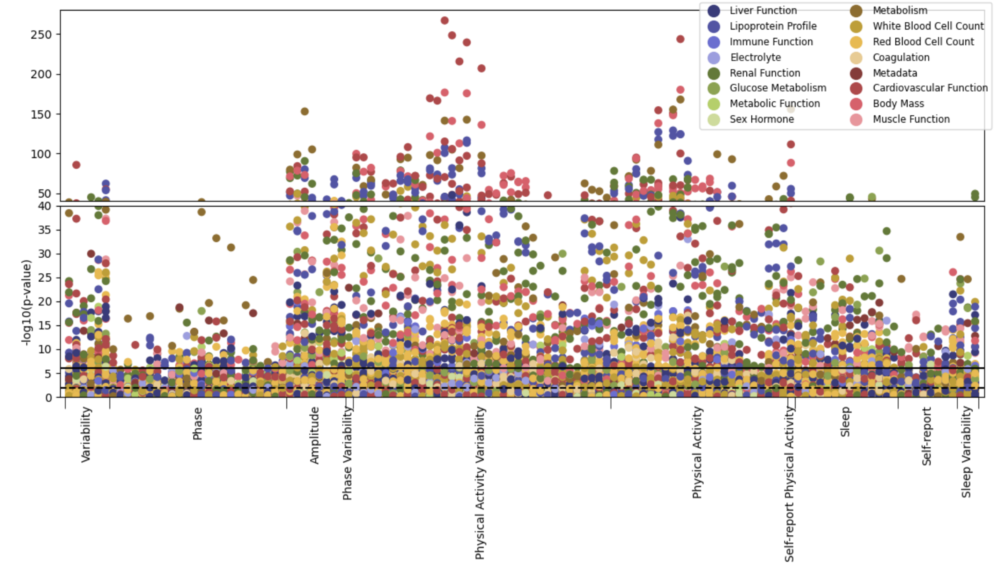

Figure S 2 – Associations between Behavioral Health Measures and Disease Phenotypes / Quantitative Traits

Manhattan-style plot of activity-phenotype association p-values, grouped by activity variable categories in the x-axis and colored by phenotype category in (top) Phecode diagnoses and (bottom) quantitative phenotypes.

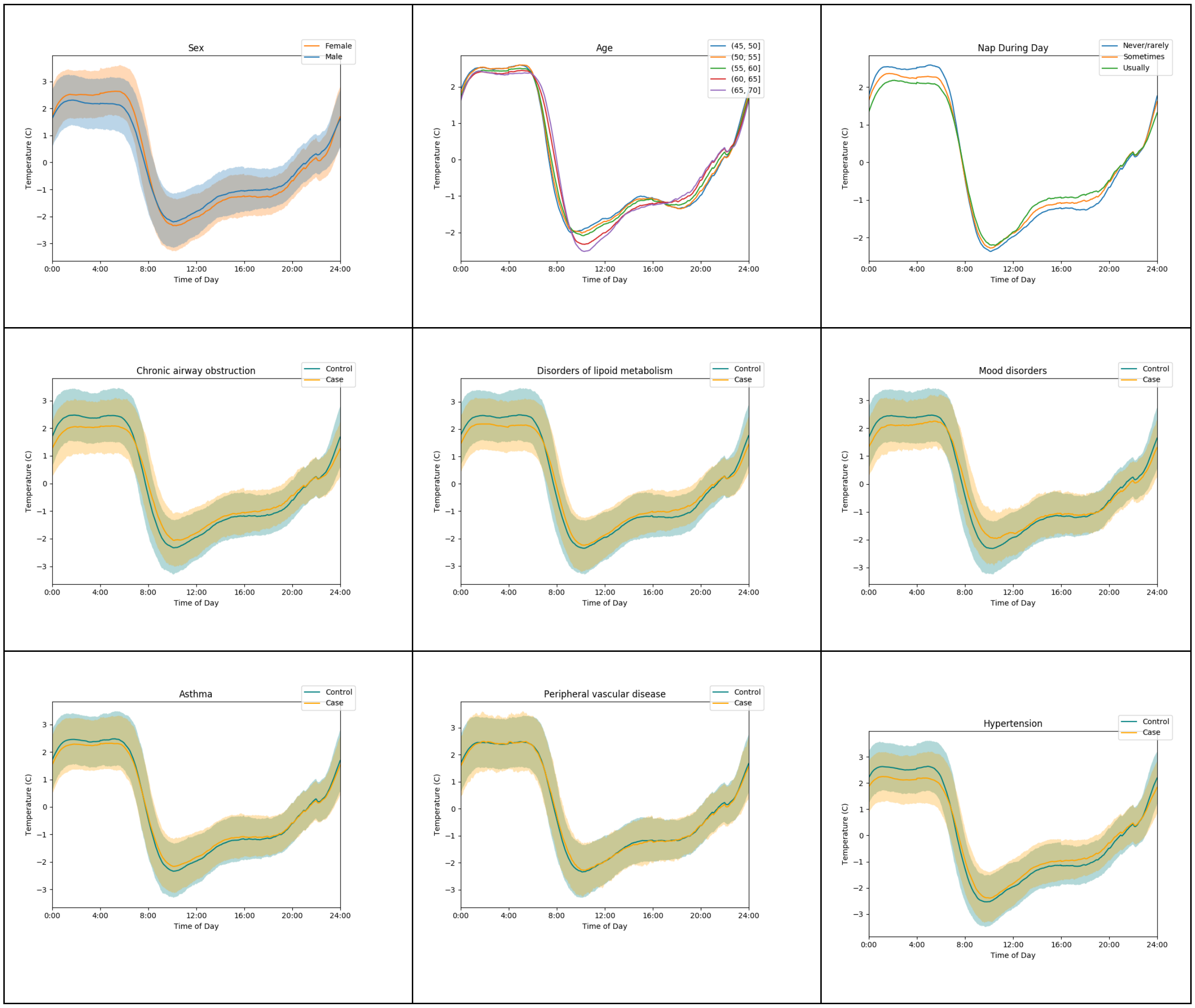

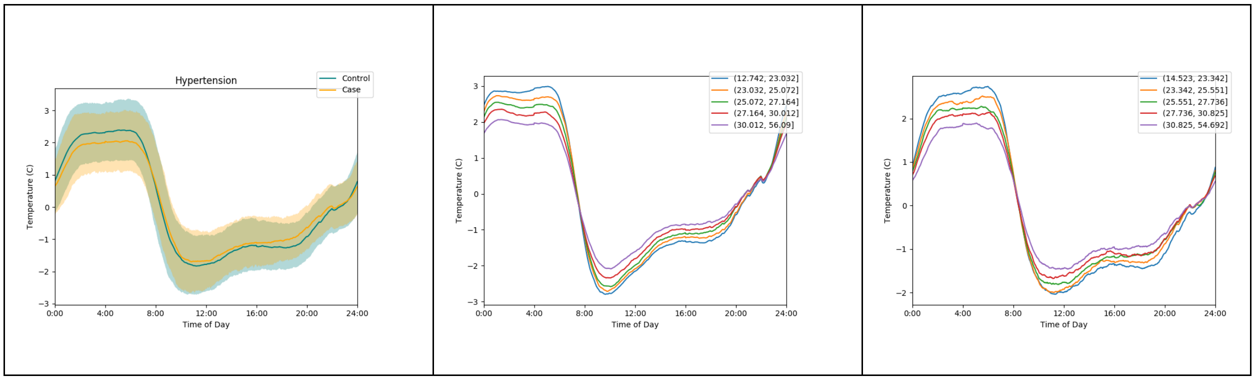


**Figure S 3 – Detailed Wrist temperature traces**

Wrist temperature traces comparing cases and controls for differences by sex, age, self-reported napping during the day as well as by disease condition for cases with chronic airway obstruction, disorders of lipoid metabolism, mood disorders, asthma and peripheral vascular disease. $25^{th}$ to $75^{th}$ percentiles of the population are displayed in shaded regions (controls in blue, cases in yellow and overlap in grayish green). Temperature is plotted relative to an individual's overall mean. To explore associations of 'chronotype of person' with 'circadianness of disease', we plotted the peripheral temperature trace for patients with hypertension and controls of self-reported morning chronotype. As expected, patients with hypertension compared to controls of self-reported evening chronotype display a right-shifted temperature-over-time curve but the shape of the curves are similar for cases and controls, suggesting that an interaction between chronotype and disease phenotype does not lead to aberrant temperature cycles. Increase in body weight (in quintiles of BMI) associates with a loss of amplitude in skin temperature which tracks with self-reported morning and evening chronotype but without indication of an interaction.

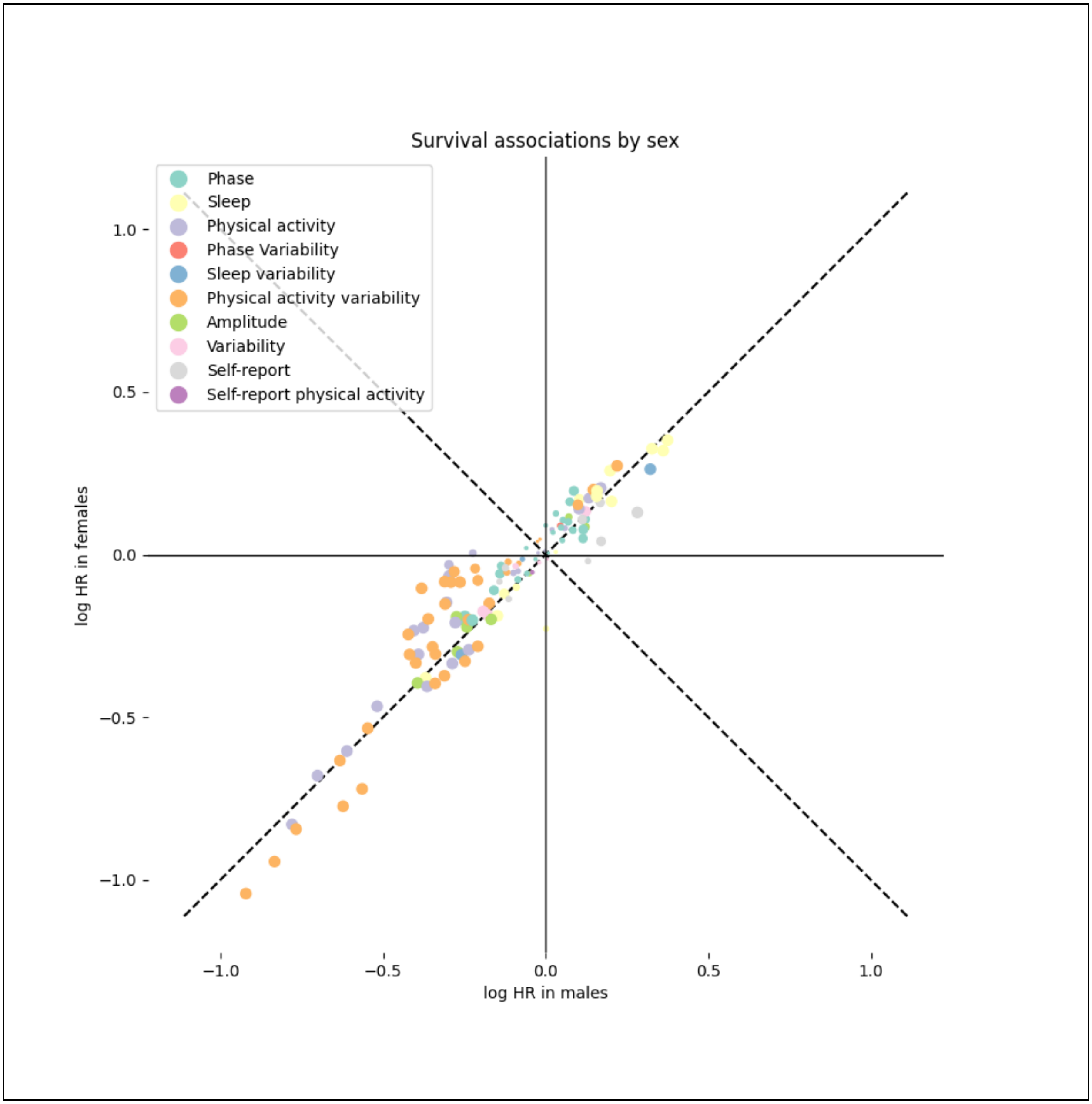


**Figure S 4 – Associations between Survival and Behavioral Health Variables**

Standardized effect sizes of survival in males plotted versus females.

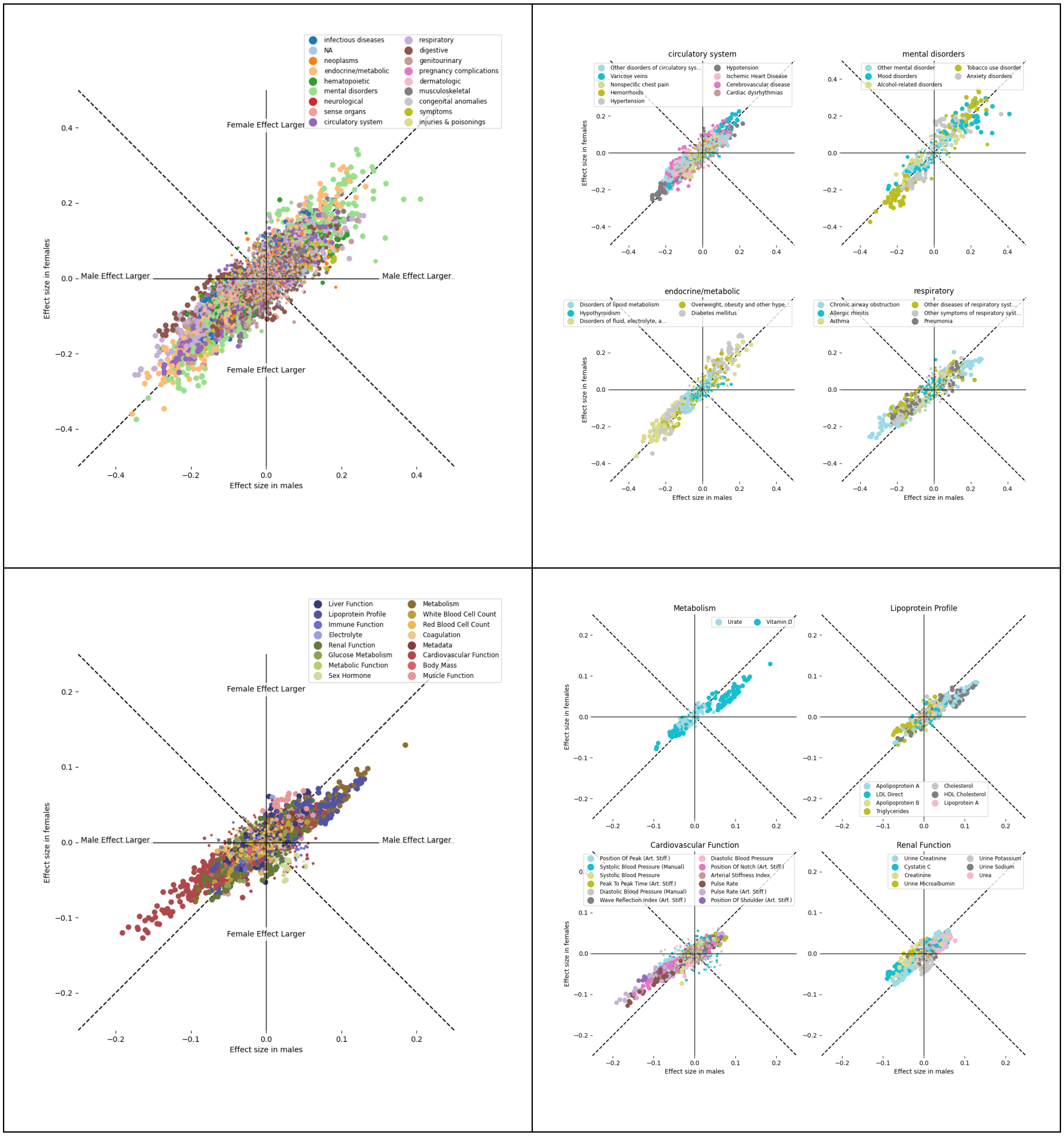
infectious diseases
NA
neoplasms
endocrine/metabolic
hematopoietic
mental disorders
neurological
sense organs
circulatory system
respiratory
digestive
genitourinary
pregnancy complications
dermatologic
musculoskeletal
congenital anomalies
symptoms
injuries & poisonings
Female Effect Larger
Male Effect Larger
Effect size in females
Effect size in males
circulatory system
mental disorders
endocrine/metabolic
respiratory
Liver Function
Lipoprotein Profile
Immune Function
Electrolyte
Renal Function
Glucose Metabolism
Metabolic Function
Sex Hormone
Metabolism
White Blood Cell Count
Red Blood Cell Count
Coagulation
Metadata
Cardiovascular Function
Body Mass
Muscle Function
Metabolism
Lipoprotein Profile
Cardiovascular Function
Renal Function

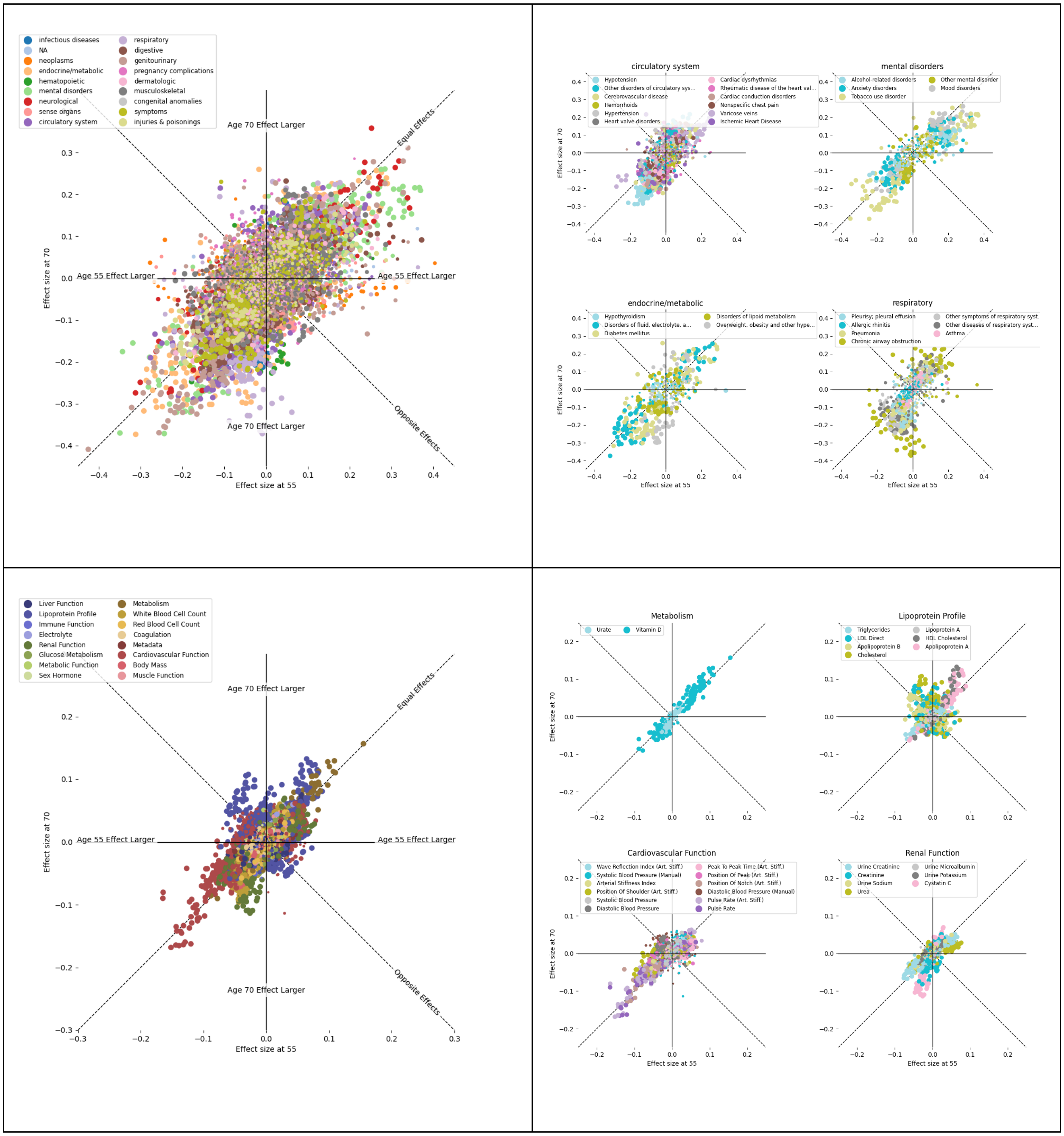


**Figure S 5 – Detailed Sex- and Age-Specific Effects**

Comparison of associations between medical diagnoses and behavioral health measures by sex and age. (a) Comparison of the effect sizes for behavioral health measure (a) and quantitative traits (c) (difference in behavioral health measure or quantitative trait between cases and controls, normalized by the standard deviation of the behavioral health measure or quantitative trait) in males versus females. Effect sizes in males versus females in select categories of phenotypes (b, behavioral health measure; d, quantitative traits). (e) Ten top sex-differential associations with acceleration_RA scores. Standardized effect sizes modelled linearly (difference in cases compared to controls in standard deviations of the actigraphy health variable) in the younger (age 55) and older (age 70) populations, by phenotype category, and within select phenotype categories of the phenotypes with at least 500 cases. Standardized effect sizes (standard deviations of phenotype per standard deviation of variable) in males versus females of all phenotypes (f), of select phenotype categories (g). Standardized effects sizes in younger (age 55) and older (age 70) populations, (h) by phenotype category, and (i) within select phenotype categories. (j) Ten top sex-differential associations with acceleration_RA scores. Lines denote 95% confidence intervals.

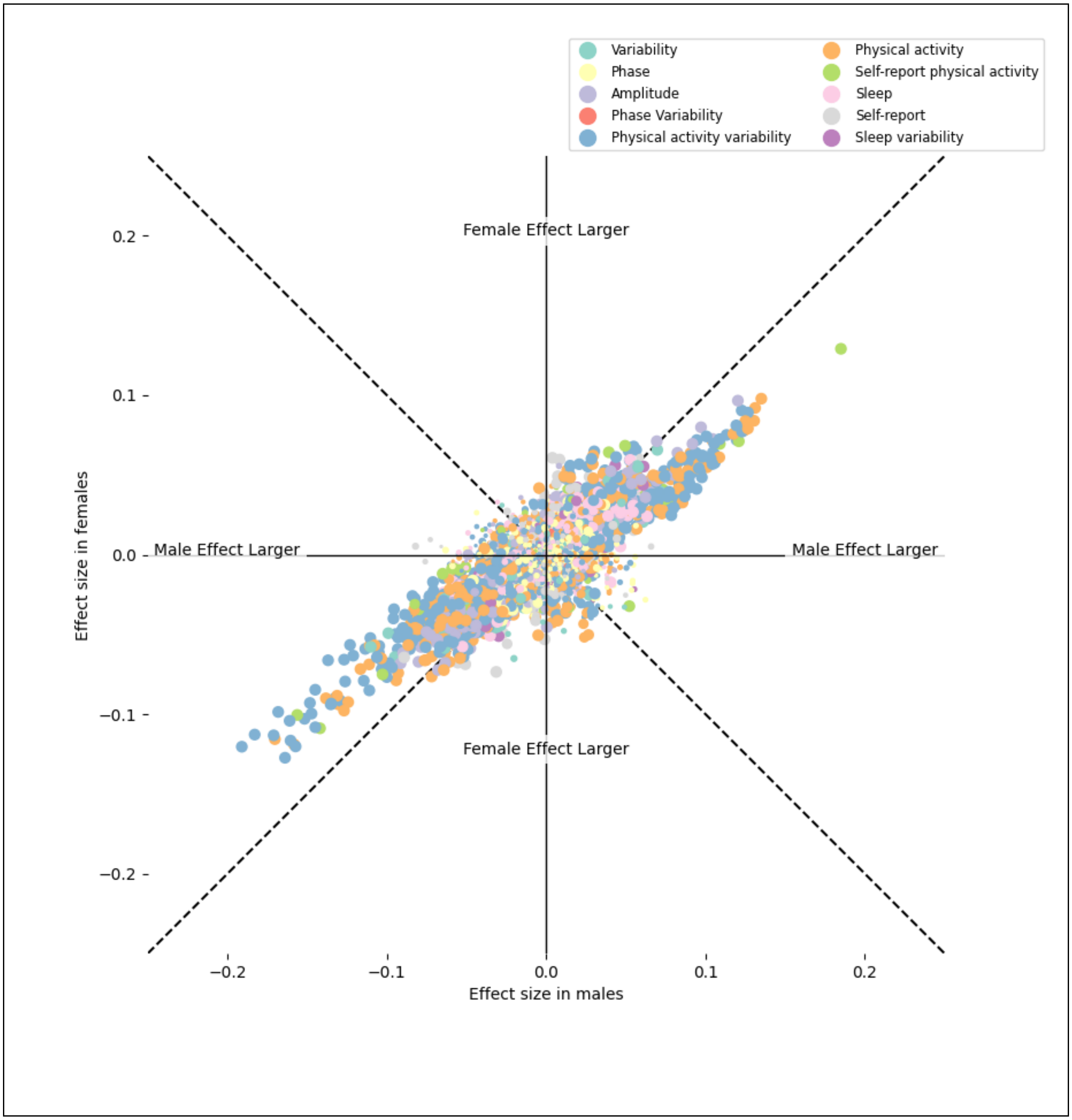


**Figure S 6 – Sexual Dimorphism in Quantitative Phenotypes is Driven by Physical Activity Measures**

Standardized effect sizes (standard deviations of phenotype per standard deviation of behavioral health measure) in males versus females of all phenotypes, colored by actigraphy health variable category. The most sexually dimorphic associations are within physical activity and physical activity variability measures and match effects seen with the self-reported physical activity measure.

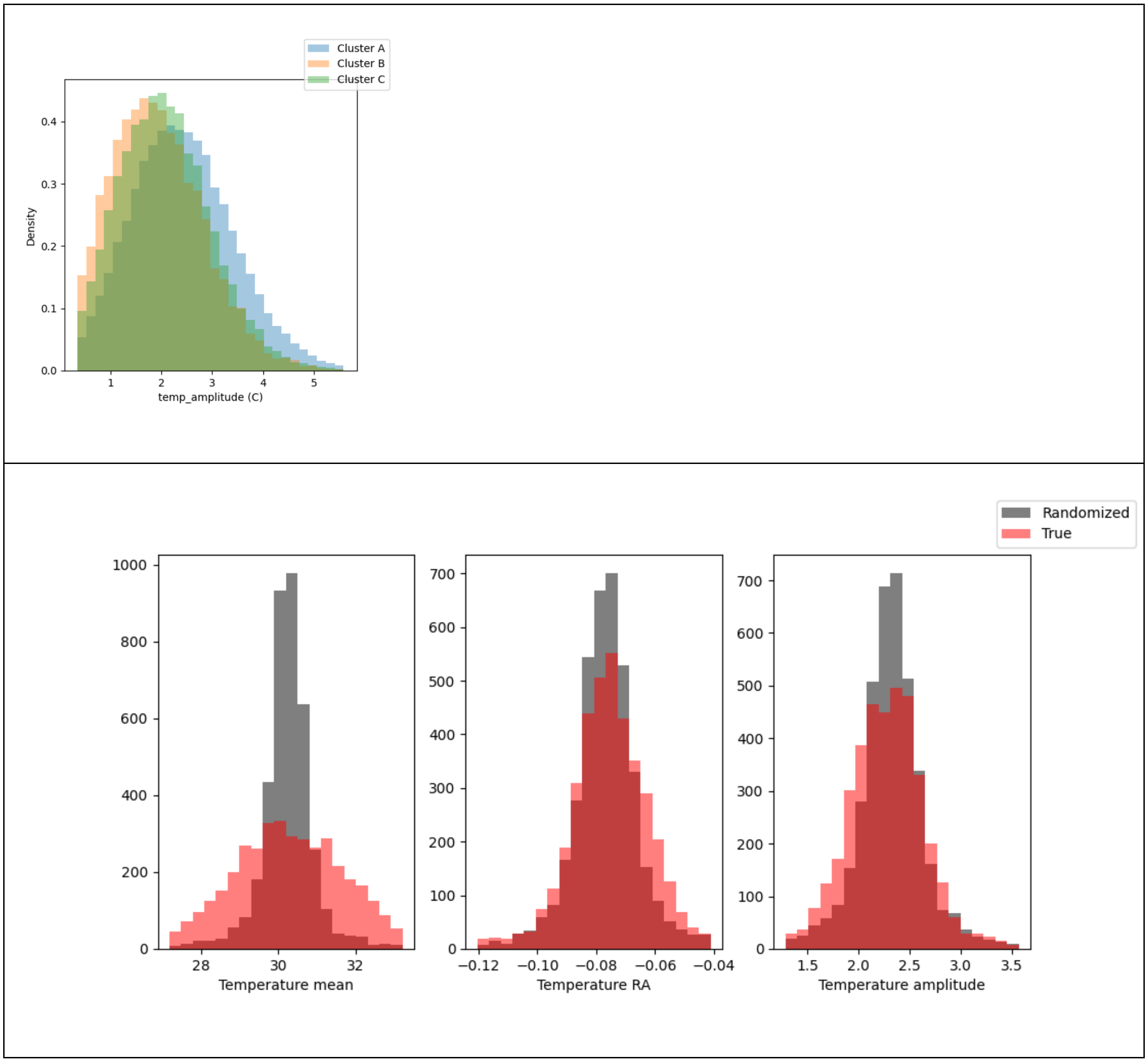


Figure S 7 – Calibration of temperature data

(Top) The distributions of the temperature cosinor amplitude (temp_amplitude) in each of three clusters of actigraphy devices, defined by ID ranges. (Cluster A: device IDs up to 7500, Cluster B: 7501 through 12500, Cluster C: 12501 and above). (Bottom row) The distribution of average (left) temperature mean, (center) temperature RA, and (right) temperature cosinor amplitude scores among all measurements from each device, in red, compared to the distribution after a random permutation of device IDs (in grey) to show expected variance. Wider distributions than by random permutation indicate device-specific biases.

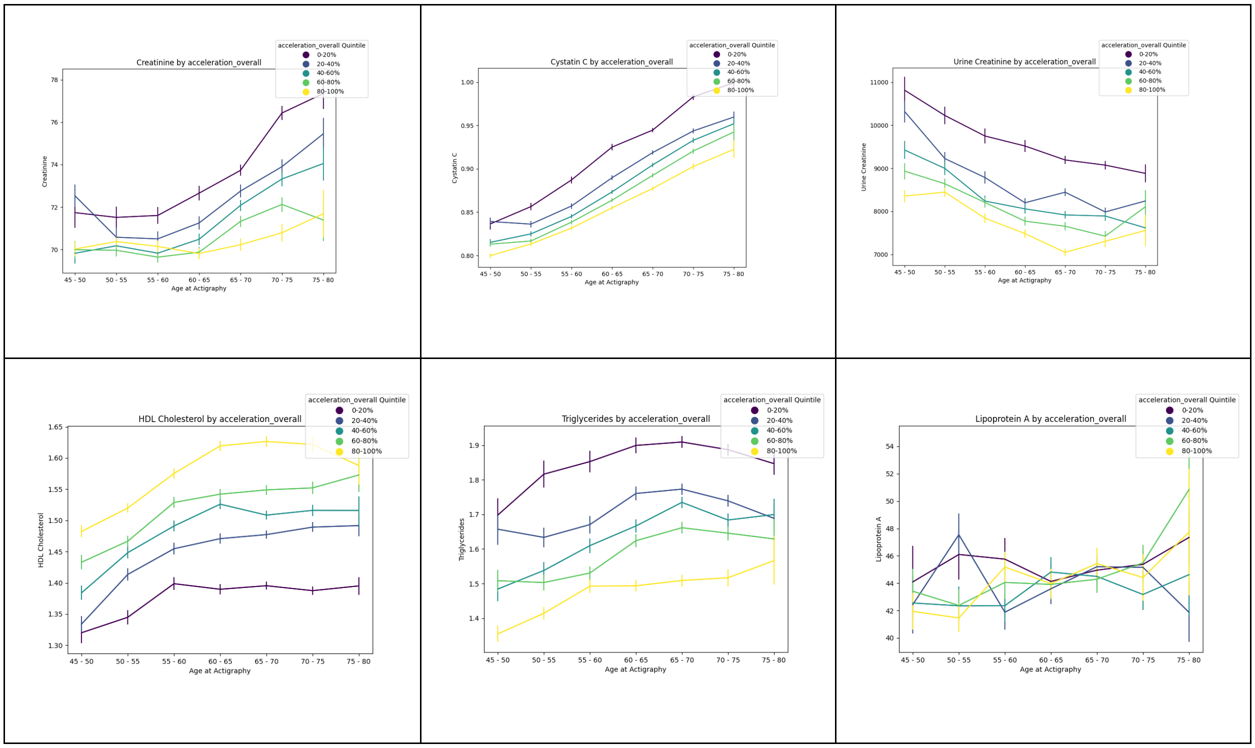


Figure S 8 – Interactions between age & physical activity for selected quantitative traits

Mean values of plasma creatinine, cystatin C and urine creatinine (top) and HDL Cholesterol, triglycerides and lipoprotein A (bottom) by age at actigraphy broken up by overall acceleration levels quintiles. All line plots have 95% confidence intervals shown.

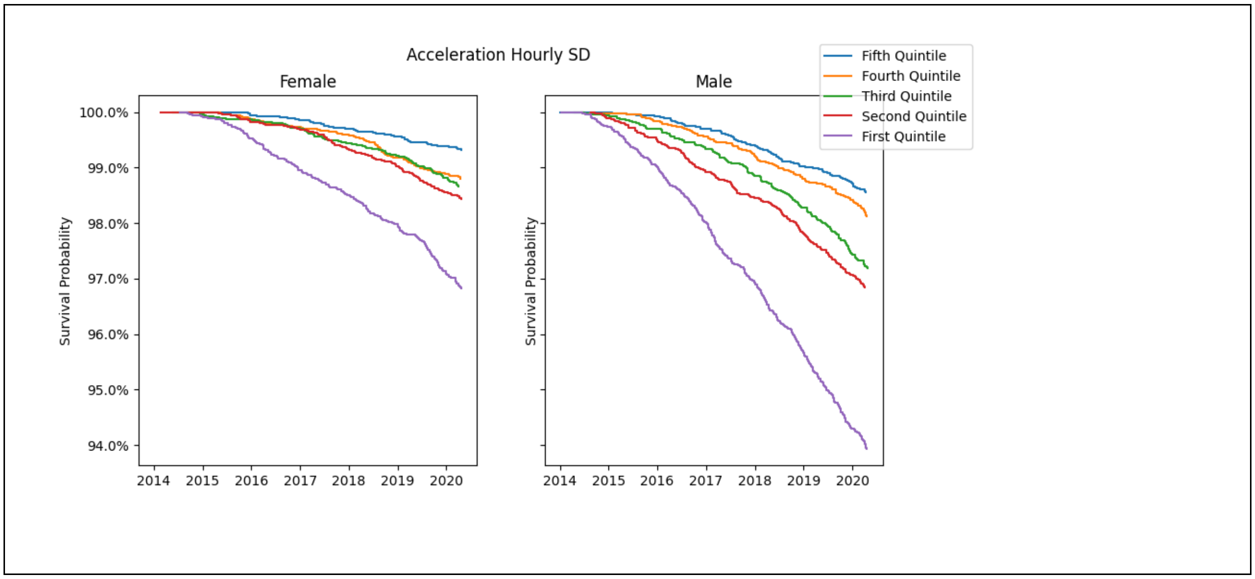

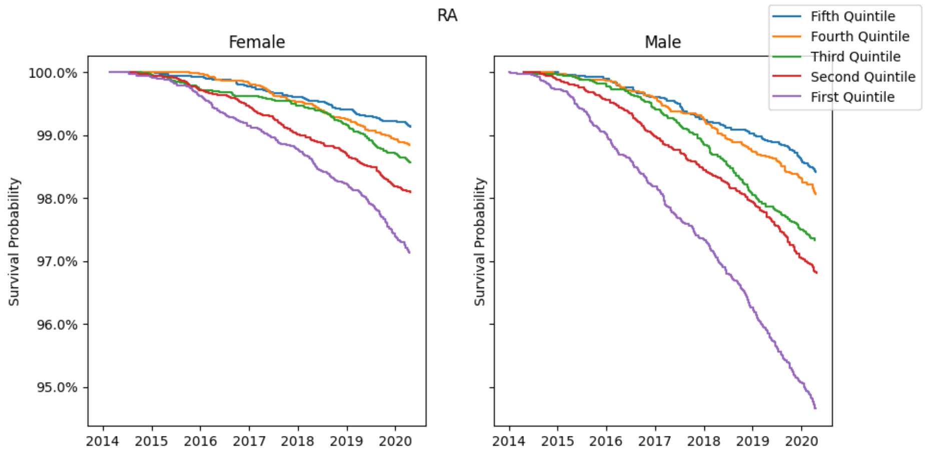
RA
Female
Male
Fifth Quintile
Fourth Quintile
Third Quintile
Second Quintile
First Quintile
Survival Probability
100.0%
99.0%
98.0%
97.0%
96.0%
95.0%
2014
2015
2016
2017
2018
2019
2020

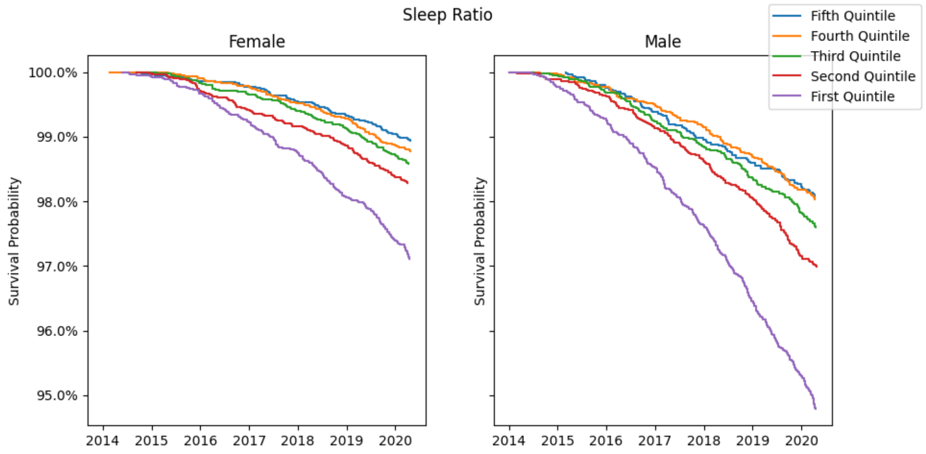
Sleep Ratio
Female
Male
Fifth Quintile
Fourth Quintile
Third Quintile
Second Quintile
First Quintile
Survival Probability
100.0%
99.0%
98.0%
97.0%
96.0%
95.0%
2014
2015
2016
2017
2018
2019
2020

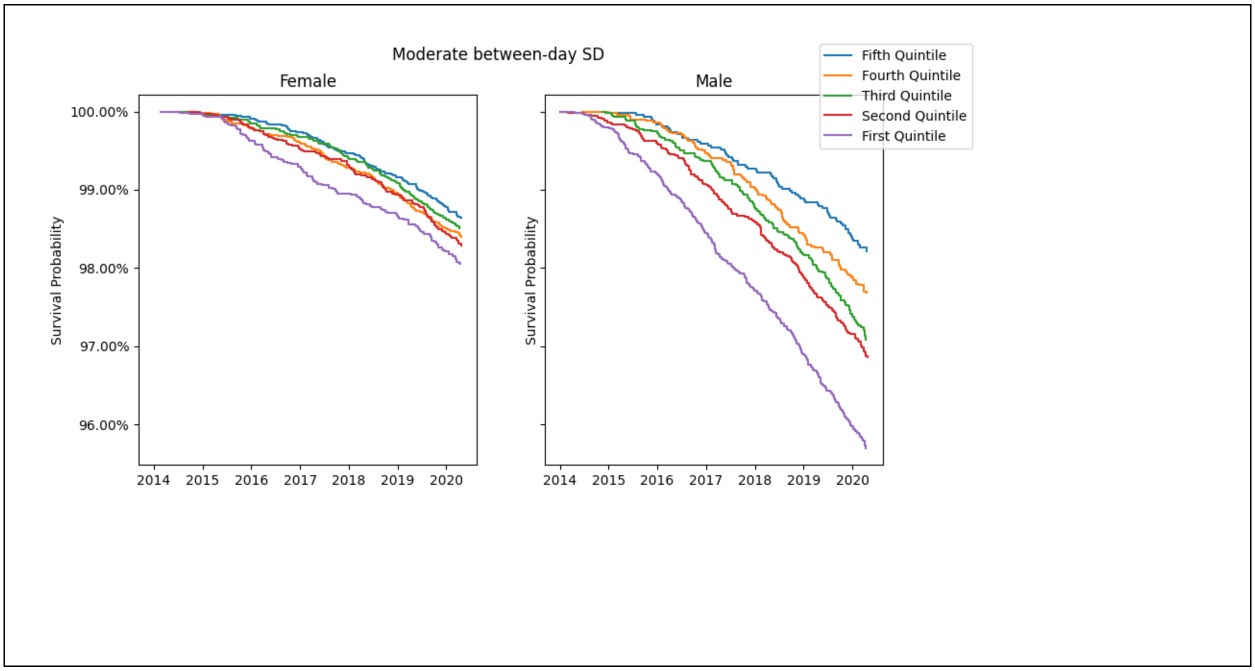


**Figure S 9 – Sex-Dependent Quintile Distribution of Physical Activity, Sleep and Diurnal Rhythmicity**

Survival curves separated by sex for four activity variables (acceleration_hourly_SD, acceleration_RA, main_sleep_ratio_mean, and moderate_between_day_SD).

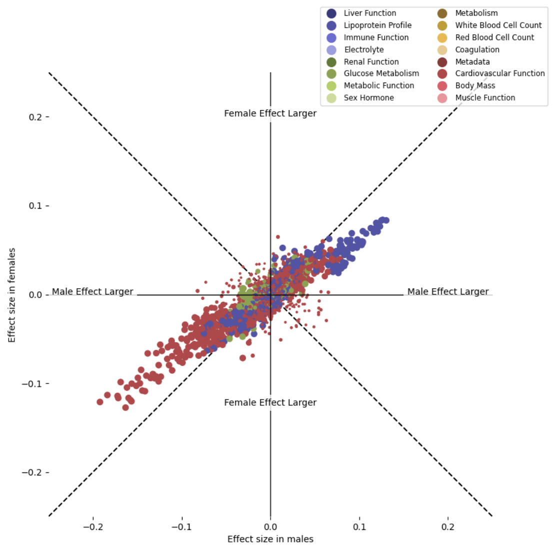


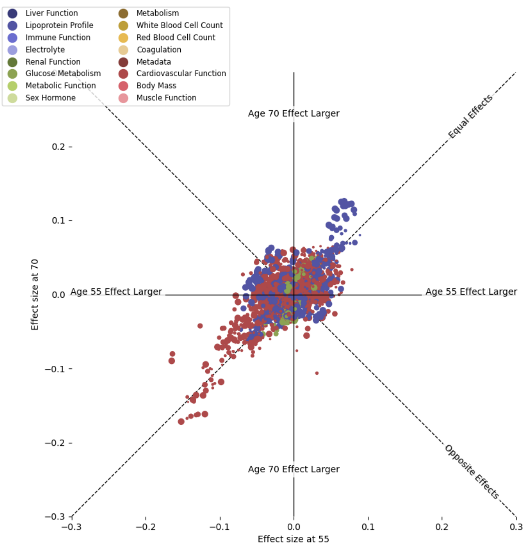


Without medication

With medication

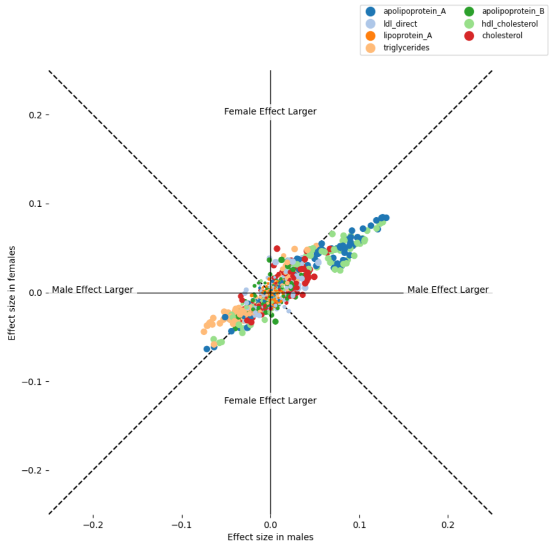


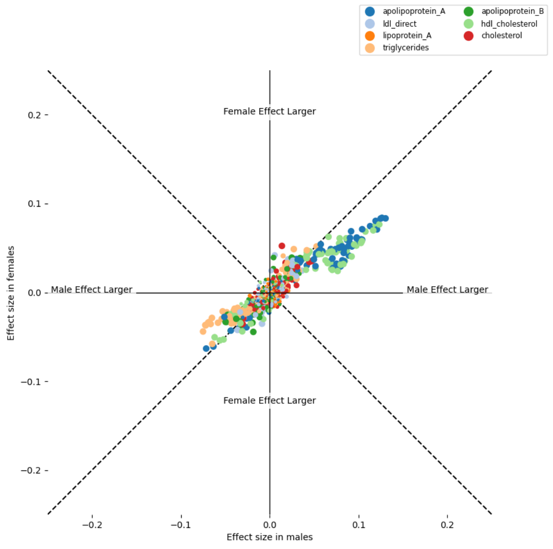


Without medication

With medication

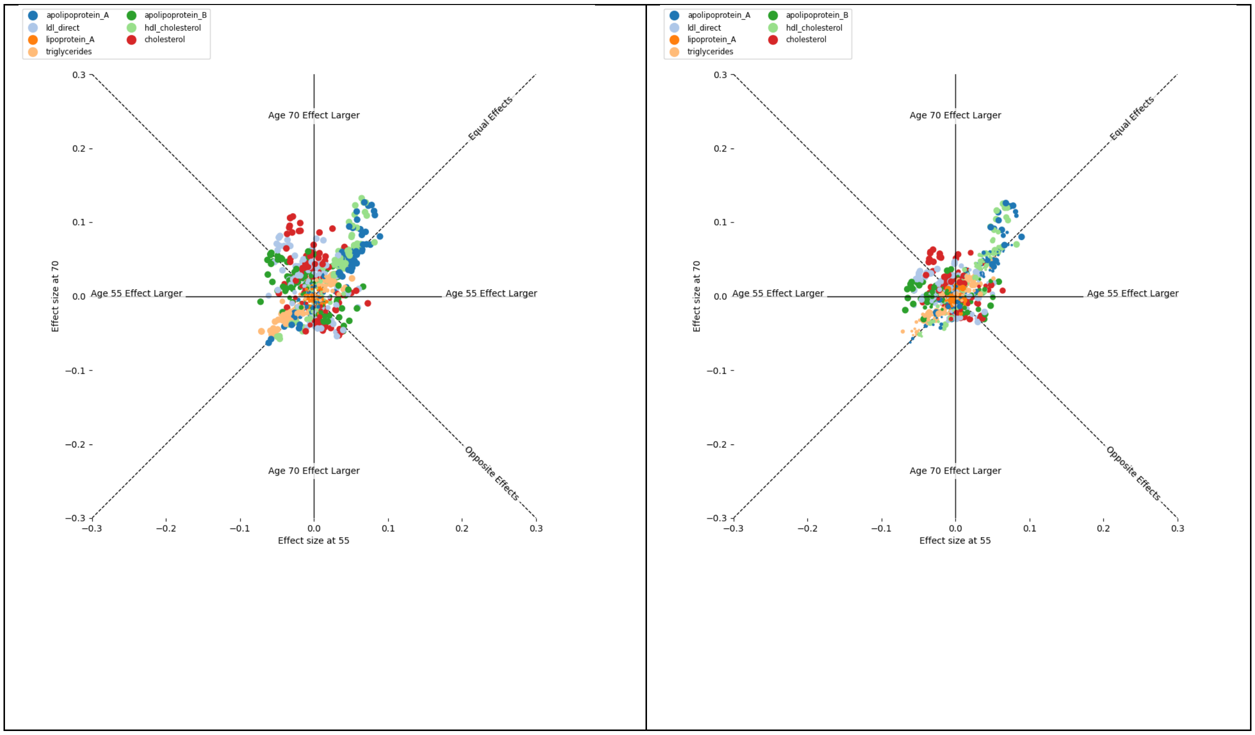


Figure S 10 – Medication-controlled Quantitative Traits

Sex-specific (top left) and age-specific (top right) actigraphy associations in lipoprotein profile, glucose metabolism, and cardiovascular function variables after controlling for medication status. Effects on the lipoprotein profile without medication control (center and bottom left) and with medication control (center and bottom right).

| Sex-specific trajectories of LDL-C over age categorized by quantitatively and qualitatively assessed physical activity | Sex-specific trajectories of LDL-C over age categorized by quantitatively and qualitatively assessed physical activity limited to individuals not reporting intake of statins | Sex-specific trajectories of LDL-C over age categorized by quantitatively and qualitatively assessed physical activity limited to individuals reporting intake of statins |
|---|---|---|
| 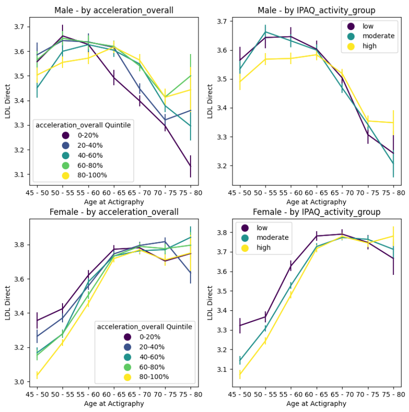  | 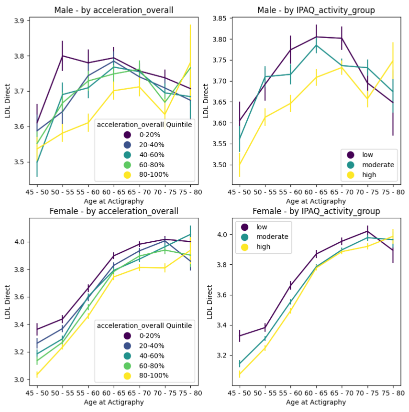  | 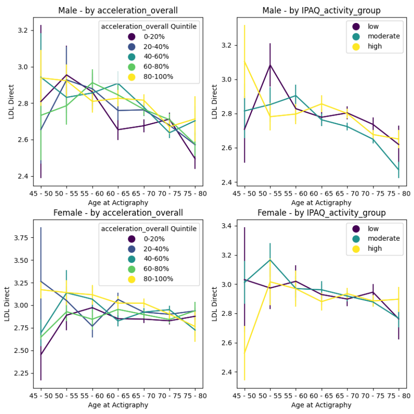  |

Figure S 11 - Age-specific LDL-C levels modified by sex and prescription medicine intake

**Supplemental Tables**

Table S 1 - Demographics of people with and without actigraphy in the UK Biobank.

| | With Actigraphy | Without Actigraphy |
|---|---|---|
| **Male** | 44% | 46% |
| **Female** | 56% | 54% |
| **White** | 97% | 93% |
| **Nonwhite** | 3% | 7% |
| **Birth Year (SD)** | 1952.0 (7.8) | 1951.4 (8.2) |
| **BMI (SD)** | 26.72 (4.5) | 27.62 (4.9) |

Table S 2 – Hazard Ratios for select diagnoses in exploratory cohort

Same as Table 1 but computed in the smaller, independent exploratory cohort of 25,000 individuals.

| Activity Variable | Hazard Ratio From 1 SD increase (95% CI) | Hazard Ratio From 2 SD increase (95% CI) |
|---|---|---|

| | | |
|---|---|---|
| **Diabetes** | 1.29 (1.2-1.39) | 1.67 (1.44-1.93) |
| **Hypertension** | 1.10 (1.06-1.14) | 1.21 (1.12-1.3) |
| **Renal Failure** | 1.19 (1.08-1.3) | 1.41 (1.17-1.69) |
| **Chronic Airway Obstruction** | 1.18 (1.07-1.3) | 1.39 (1.15-1.68) |
| **Pneumonia** | 1.18 (1.08-1.28) | 1.38 (1.16-1.65) |
| **Anxiety Disorders** | 1.12 (1.02-1.23) | 1.26 (1.05-1.52) |
| **Disorders of Lipoid Metabolism** | 1.11 (1.05-1.17) | 1.23 (1.11-1.38) |

Table S 3 – Main Results

Contains variable definitions and statistics, survival associations, and phecodes.

Table S 4 – Phewas Results

Contains the PheWAS-SensR results on diagnosis PheCODEs, as well as sex and age interaction results.

Table S 5 – Phewas Quantitative Results

Contains the PheWAS-SensR results run on quantitative measures (blood work, urine assays, physical measures), as well as sex and age interaction results.

Table S 6 – Three Component Results

Contains the associations of PheCODE and quantitative measures in models containing three activity variables for diurnal rhythms, physical activity, and sleep quality.

Table S 7 – Proportional Hazards Results

Contains results of proportional hazard models for predictive power of circadian rhythm (measured by temp_RA), also including age and sex-specific effects.

**Supplementary Files**

Supplementary File 1:

Zip file containing all results of the smaller exploratory cohort 1. Contains figures and tables as presented in this paper but generated in the exploratory cohort for comparison in an independent random sample.

### References


1. Kaskie, R.E., B. Graziano, and F. Ferrarelli, *Schizophrenia and sleep disorders: links, risks, and management challenges.* Nat Sci Sleep, 2017. **9**: p. 227-239.

2. Liguori, C., et al., *Sleep problems affect quality of life in Parkinson's disease along disease progression.* Sleep Med, 2021. **81**: p. 307-311.

3. Madrid-Navarro, C.J., et al., *Validation of a Device for the Ambulatory Monitoring of Sleep Patterns: A Pilot Study on Parkinson's Disease.* Front Neurol, 2019. **10**: p. 356.

4. Biswas, A., et al., *Sedentary time and its association with risk for disease incidence, mortality, and hospitalization in adults: a systematic review and meta-analysis.* Ann Intern Med, 2015. **162**(2): p. 123-32.

5. Kurina, L.M., et al., *Sleep duration and all-cause mortality: a critical review of measurement and associations.* Ann Epidemiol, 2013. **23**(6): p. 361-70.

6. Garfield, V., et al., *The relationship between sleep quality and all-cause, CVD and cancer mortality: the Southall and Brent REvisited study (SABRE).* Sleep Med, 2019. **60**: p. 230-235.

7. Wallace, M.L., et al., *Which Sleep Health Characteristics Predict All-Cause Mortality in Older Men? An Application of Flexible Multivariable Approaches.* Sleep, 2018. **41**(1).

8. Jenkins, J.T. and P.J. O'Dwyer, *Inguinal hernias.* BMJ, 2008. **336**(7638): p. 269-72.

9. Kingsnorth, A. and K. LeBlanc, *Hernias: inguinal and incisional.* Lancet, 2003. **362**(9395): p. 1561-71.

10. Simons, M.P., et al., *European Hernia Society guidelines on the treatment of inguinal hernia in adult patients.* Hernia, 2009. **13**(4): p. 343-403.

11. Tolver, M.A., et al., *Female gender is a risk factor for pain, discomfort, and fatigue after laparoscopic groin hernia repair.* Hernia, 2013. **17**(3): p. 321-7.

12. Hombali, A., et al., *Prevalence and correlates of sleep disorder symptoms in psychiatric disorders.* Psychiatry Res, 2019. **279**: p. 116-122.

13. Mallampalli, M.P. and C.L. Carter, *Exploring sex and gender differences in sleep health: a Society for Women's Health Research Report.* J Womens Health (Larchmt), 2014. **23**(7): p. 553-62.

14. Boccabella, A. and J. Malouf, *How Do Sleep-Related Health Problems Affect Functional Status According to Sex?* J Clin Sleep Med, 2017. **13**(5): p. 685-692.

15. https://www.gundersenhealth.org/health-wellness/move/physical-activity/minutes-in-motion/pedometer-conversion-chart/. **Accessed August 10, 2021**.

16. Lin, C.L., et al., *Association between sleep disorders and hypertension in Taiwan: a nationwide population-based retrospective cohort study.* J Hum Hypertens, 2017. **31**(3): p. 220-224.

17. Odden, M.C., et al., *Age and cystatin C in healthy adults: a collaborative study.* Nephrol Dial Transplant, 2010. **25**(2): p. 463-9.

18. Baxmann, A.C., et al., *Influence of muscle mass and physical activity on serum and urinary creatinine and serum cystatin C.* Clin J Am Soc Nephrol, 2008. **3**(2): p. 348-54.

19. Sato, T., et al., *Association between physical activity and change in renal function in patients after acute myocardial infarction.* PLoS One, 2019. **14**(2): p. e0212100.

20. Barr, D.B., et al., *Urinary creatinine concentrations in the U.S. population: implications for urinary biologic monitoring measurements.* Environ Health Perspect, 2005. **113**(2): p. 192-200.

21. Wilson, D.P., et al., *Use of Lipoprotein(a) in clinical practice: A biomarker whose time has come. A scientific statement from the National Lipid Association.* J Clin Lipidol, 2019. **13**(3): p. 374-392.

22. Bahls, M., et al., *Statins are related to impaired exercise capacity in males but not females.* PLoS One, 2017. **12**(6): p. e0179534.

23. Parker, B.A., et al., *Effect of statins on skeletal muscle function.* Circulation, 2013. **127**(1): p. 96-103.

24. Agarwala, A., et al., *The Use of Sex-Specific Factors in the Assessment of Women's Cardiovascular Risk.* Circulation, 2020. **141**(7): p. 592-599.

25. Doherty, A., et al., *Large Scale Population Assessment of Physical Activity Using Wrist Worn Accelerometers: The UK Biobank Study.* PLoS One, 2017. **12**(2): p. e0169649.

26. Willetts, M., et al., *Statistical machine learning of sleep and physical activity phenotypes from sensor data in 96,220 UK Biobank participants.* Sci Rep, 2018. **8**(1): p. 7961.

27. Sanford, S.D., et al., *Gender differences in sleep, fatigue, and daytime activity in a pediatric oncology sample receiving dexamethasone.* J Pediatr Psychol, 2008. **33**(3): p. 298-306.

28. Short, M.A., et al., *The discrepancy between actigraphic and sleep diary measures of sleep in adolescents.* Sleep Med, 2012. **13**(4): p. 378-84.

29. Marino, M., et al., *Measuring sleep: accuracy, sensitivity, and specificity of wrist actigraphy compared to polysomnography.* Sleep, 2013. **36**(11): p. 1747-55.

30. Witting, W., et al., *Alterations in the circadian rest-activity rhythm in aging and Alzheimer's disease.* Biol Psychiatry, 1990. **27**(6): p. 563-72.

31. Brown, A.C., et al., *Actigraphy: a means of assessing circadian patterns in human activity.* Chronobiol Int, 1990. **7**(2): p. 125-33.

32. Lyall, L.M., et al., *Association of disrupted circadian rhythmicity with mood disorders, subjective wellbeing, and cognitive function: a cross-sectional study of 91 105 participants from the UK Biobank.* Lancet Psychiatry, 2018. **5**(6): p. 507-514.

33. Kennard, H.R., et al., *The associations between thermal variety and health: Implications for space heating energy use.* PLoS One, 2020. **15**(7): p. e0236116.
